# Beyond ERBB2 amplification: RNA and proteomic HER2 assessment for pan-tumor treatment stratification

**DOI:** 10.64898/2026.10.11.26365242

**Authors:** Kristian Egebjerg, Kristoffer Staal Rohrberg, Frederik Otzen Bagger, Anne-Vibeke Lænkholm, Maj-Lis Møller Talman, Annelaura Bach Nielsen, Juanjuan Wang, Eduard Chelebian, Maximilian T. Strauss, Andreas Mund, Xiang Zheng, Rajendra Singh Garbyal, Mette Kjølhede Nedergaard, Benjamin H. Conlon, Luca Robinson, Tina Kringelbach, Martin Højgaard, Ane Yde Schmidt, Matilde Fjeldsø Larsen, Daniela Alosi, Annette Raskov Kodahl, Rikke Løvendal Eefsen, Morten Ladekarl, Lise Søndergaard, Lars Henrik Jensen, Ulrik Niels Lassen, Morten Mau-Sørensen

**Author notes:** **Correspondence:** Kristian Egebjerg.

## Abstract

HER2-directed therapies are expanding across solid tumors, but ERBB2 amplification incompletely captures HER2 expression and pan-tumor IHC lacks standardization. We developed ERBB2 mRNA, bulk HER2 protein and RNA-signature screening benchmarked against IHC/FISH. Screening identified HER2-enriched populations beyond amplification in a pan-tumor discovery cohort of 1,331 tumor biopsies and validated RNA-based screening in an external cohort of 3,898 tumor biopsies. All six objective responses among 51 treated patients in the discovery cohort occurred in tumors positive by ERBB2 mRNA and the combined RNA-signature. In the external dataset (n=102), ERBB2 mRNA positivity was associated with longer time on treatment (hazard ratio 0.42, 95% confidence interval 0.27–0.64) and overall survival (0.41, 0.25–0.65), with concordant associations for the combined RNA signature. Spatial proteomics comprising 2,721 tiles across 88 samples captured HER2 heterogeneity and identified HER2-enriched profiles below bulk thresholds. These findings support prospective evaluation of RNA and proteomic screening for treatment selection.

## Introduction

HER2-targeted therapy has transformed the treatment of HER2-positive breast and gastroesophageal adenocarcinomas^1,2^. In these indications, HER2 testing is embedded in routine clinical care through standardized immunohistochemistry (IHC) scoring, with reflex in situ hybridization (ISH) for equivocal IHC2+ cases^3,4^. This diagnostic framework has been refined over more than two decades through indication-specific scoring criteria, assay standardization, pre-analytical quality control and clinical validation, making HER2 IHC/ISH one of the most established companion diagnostics in solid tumors.

The therapeutic relevance of HER2 has now expanded beyond its conventional disease settings. HER2-directed antibody-drug conjugates have demonstrated clinically meaningful activity across multiple HER2-expressing solid tumors, shifting HER2 from an indication-specific biomarker toward a pan-tumor treatment target^5^. Accordingly, HER2 IHC3+ status has become a treatment-selection criterion for selected patients with unresectable or metastatic solid tumors ^6^. However, the diagnostic frameworks established in breast and gastroesophageal cancers may not transfer directly across tumor types because pan-tumor HER2 assessment lacks universally standardized IHC scoring criteria^7^.

*ERBB2* amplification provides a highly specific genomic marker of HER2-driven disease but incompletely captures HER2 expression at the protein level. Although amplification frequently results in high HER2 expression, real-world and pan-tumor trial data show that many HER2 IHC-positive tumors lack detectable amplification by orthogonal genomic assays ^8,9^. In DESTINY-PanTumor02, *ERBB2* amplification was detected in only 35% of HER2 IHC2+ or IHC3+ tumors by tissue ISH and in 18% by circulating tumor DNA ^9^. *ERBB2* amplification is therefore informative when present but cannot serve as a complete surrogate for HER2 expression.

Conventional HER2 IHC also presents challenges in the pan-tumor setting. At the time of the tumor-agnostic approval of trastuzumab deruxtecan, no FDA-approved companion diagnostic was available for identifying HER2 IHC3+ solid tumors across indications ^7^. In DESTINY-PanTumor02, agreement between local and central HER2 scoring was 59% for IHC3+ tumors and 55% for IHC2+ tumors ^9^. Cross-algorithm analyses have also demonstrated poor concordance among established breast, gastric and endometrial scoring systems, together with limited agreement among pathologists experienced in HER2 assessment ^10^. In a large real-world analysis of 65,075 solid tumors assessed by HER2 IHC, only 3.1% were classified as IHC3+ overall, although prevalence varied considerably across tumor types, highlighting the relatively low yield of broad pan-tumor IHC screening^11^.

These limitations motivate complementary HER2 assessment within existing molecular-profiling workflows. Comprehensive molecular profiling captures ERBB2 alterations at the DNA level, ERBB2 transcriptional output by RNA-seq and HER2 protein abundance by mass spectrometry (MS) ^8,11–13^. Beyond these bulk approaches, spatial proteomic methods can preserve information on intratumoral distribution ^14,15^. We therefore integrated centrally reviewed HER2 IHC/FISH with ERBB2 copy number, RNA sequencing, mass spectrometry-based proteomics and deep visual proteomics. Using breast and gastroesophageal cancers as clinical references, we benchmarked molecular HER2 measurements, applied them to internal pan-tumor screening and treated cohorts, validated RNA-based approaches externally, and evaluated spatial HER2 profiling as a distribution-aware extension of bulk assessment.

## Results

### 1. *ERBB2* copy number incompletely captures clinical HER2 biology

To establish a clinically anchored framework for pan-tumor HER2 screening, we analyzed breast and gastroesophageal cancers, where HER2 assessment by IHC/FISH is an established treatment-selection standard. The reference cohort included 181 patients with breast cancer and 48 patients with gastroesophageal adenocarcinoma (Figure 1; Supplementary Table 1). Central HER2 assessment identified 32 HER2-positive breast cancers and 10 HER2-positive gastroesophageal cancers; IHC scores were available for 223 tumors, FISH results for 215, and both for 209 (Supplementary Fig. 1). Matched *ERBB2* copy-number data, RNA-seq, MS-based HER2 protein quantification and quantitative image-based staining features were available for overlapping subsets.

**Fig. 1.**
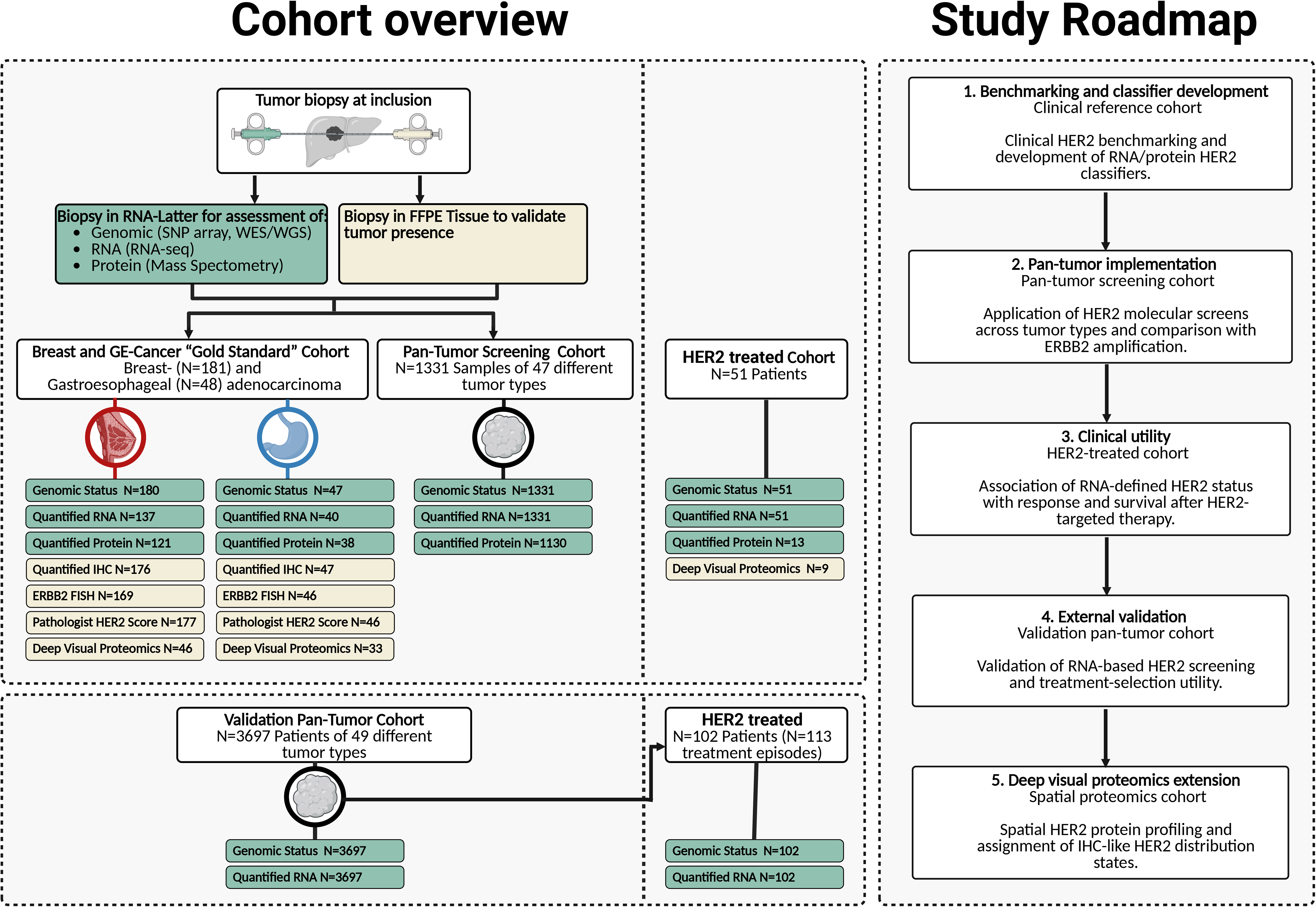
Cohort overview and study roadmap. Overview of the study cohorts and analytical workflow. Left, schematic of the cohort structure and available molecular, pathological and spatial proteomic data. Tumor biopsies collected at inclusion were processed for genomic profiling, RNA-sequencing and mass spectrometry-based proteomics, with parallel FFPE tissue used for histological confirmation and HER2 assessment. The breast and gastroesophageal reference cohort was used for clinical HER2 benchmarking and classifier development. The pan-tumor screening cohort was used to implement molecular HER2 screening across tumor types, and HER2-treated cohorts were used to evaluate associations between molecular HER2 status and clinical outcome. External validation cohorts were analyzed independently using the same prespecified molecular screening framework. Numbers indicate available samples or patients for each data modality. Right, study roadmap summarizing the sequential analytical strategy: clinical benchmarking and classifier development in breast and gastroesophageal cancers; pan-tumor implementation of molecular HER2 screening; evaluation of clinical utility in HER2-treated patients; external validation; and spatial HER2 profiling using deep visual proteomics. FFPE, formalin-fixed paraffin-embedded; FISH, fluorescence in situ hybridization; GE, gastroesophageal; IHC, immunohistochemistry; RNA-seq, RNA-sequencing; WES, whole-exome sequencing; WGS, whole-genome sequencing.

Among 219 tumors with available ERBB2 copy-number calls and clinical IHC/FISH status, ERBB2 amplification showed high specificity but limited sensitivity for clinical HER2 positivity, with a sensitivity of 54.8%, specificity of 99.4%, positive predictive value of 95.8% and overall accuracy of 90.9% (Figure 2A; Supplementary Fig. 3 and Supplementary Table 3). Nineteen of 42 clinically HER2-positive tumors were not classified as ERBB2-amplified. Direct comparison of IHC and FISH identified 13 discordant cases among 209 tumors with both assessments (6.2%). Notably, 9 of 30 IHC3+ tumors (30.0%) were FISH-negative (Supplementary Figs. 1, 2, 4 and 5; Supplementary Tables 2 and 4).

**Fig. 2.**
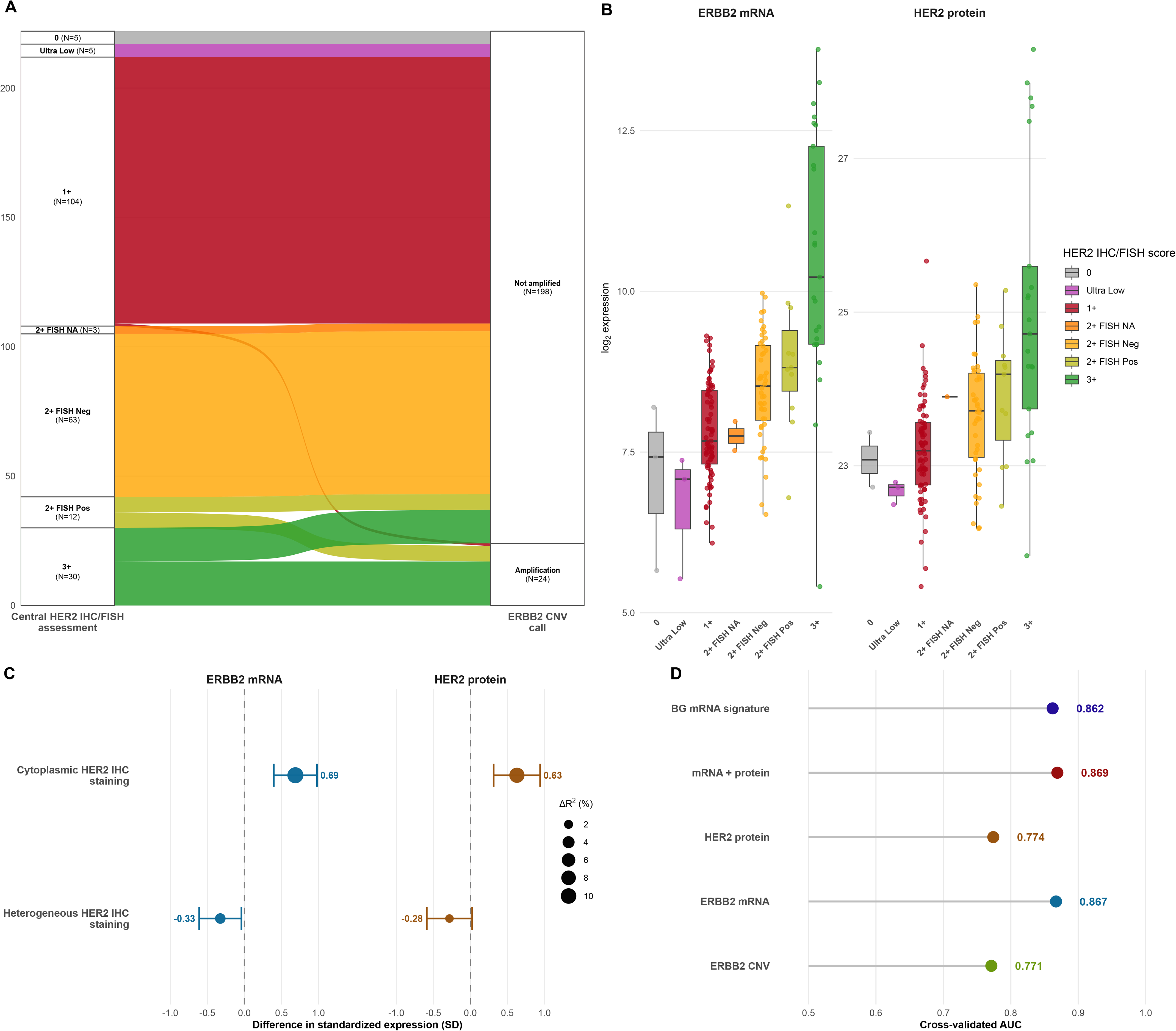
Clinical benchmarking of genomic, transcriptomic and proteomic HER2 assessment in breast and gastroesophageal cancers. **A)** Alluvial plot showing the relationship between centrally reviewed HER2 IHC/ISH categories and binary ERBB2 copy-number amplification status in the breast and gastroesophageal reference cohort. Flow widths represent the number of tumors in each category. **B)** Boxplots showing ERBB2 RNA expression and HER2 protein abundance across centrally reviewed HER2 IHC/ISH categories. Expression values are shown on the log2-transformed scale. **C)** Covariate analysis of standardized ERBB2 RNA expression and HER2 protein abundance. Points show average marginal effects for cytoplasmic staining and heterogeneous staining after adjustment for IHC/ISH category and interaction terms; point size denotes the proportion of explained variance attributable to each covariate block. **D)** Cross-validated receiver operating characteristic analysis for molecular classifiers of IHC/ISH-defined HER2-positive status, including ERBB2 copy-number amplification, ERBB2 RNA expression, HER2 protein abundance, combined RNA/protein profiling and the breast/gastroesophageal RNA-seq HER2 signature. Receiver operating characteristic analysis of molecular classifiers of IHC/ISH-defined HER2-positive status. Expression-based classifiers were evaluated using cross-validation; binary ERBB2 amplification was included as a fixed comparator. Boxplots show the median and interquartile range, with whiskers extending to 1.5 times the interquartile. IHC, immunohistochemistry; ISH, in situ hybridization; RNA-seq, RNA-sequencing; ROC, receiver operating characteristic. The alluvial includes 222

Quantitative RNA and protein profiling captured HER2 expression states across clinical categories. Both *ERBB2* mRNA and HER2 protein abundance increased progressively across ordered IHC/FISH groups, with significant ordinal trends (Figure 2B). IHC/FISH-positive tumors had higher *ERBB2* RNA and HER2 protein levels than non-positive tumors, and paired *ERBB2* RNA and HER2 protein abundance were correlated, consistent with partial RNA–protein coupling (Pearson r = 0.69, P < 0.001; Supplementary Fig. 6 and 7). However, molecular measurements overlapped across adjacent IHC/FISH categories, indicating that bulk RNA and protein readouts captured continuous HER2 expression gradients rather than simply reproducing categorical clinical scoring. Variation in staining patterns was also associated with differences in molecular abundance: cytoplasmic staining was associated with higher standardized *ERBB2* mRNA expression and HER2 protein abundance (+0.69 and +0.63 SD, respectively), whereas heterogeneous staining was associated with significantly lower *ERBB2* mRNA expression (−0.33 SD) and a trend toward lower HER2 protein abundance (−0.28 SD), after adjustment for IHC/FISH category and cancer type (Figure 2C; Supplementary Table 5; Supplementary Fig. 8).

Finally, we used the breast and gastroesophageal reference cohort to derive molecular classifiers of IHC/FISH-defined HER2 positivity: models based on ERBB2 mRNA or HER2 protein alone, an mRNA + protein model combining both measurements, and a multigene breast/gastroesophageal RNA signature (BG RNA signature) (Supplementary fig. 9). In out-of-fold evaluation, the ERBB2 mRNA model, mRNA + protein model and BG RNA signature achieved AUCs of 0.867, 0.869 and 0.862, respectively, compared with 0.774 for the HER2 protein model. At a probability threshold of 0.50, sensitivities were 53.1%, 50.0%, 47.2% and 31.2%, respectively, with specificities ranging from 95.8% to 99.3% across the models (Figure 2D; Supplementary Table 6).

### 2. Pan-tumor molecular screening identifies HER2-enriched tumors beyond *ERBB2* amplification

We next applied the breast/gastroesophageal-derived HER2 screening framework to 1,331 pan-tumor samples spanning 47 tumor types, all with genomic *ERBB2* annotation and RNA-seq data; 1,130 also had MS-based HER2 protein measurements (Figure 1; Supplementary Tables 1 and 7).

*ERBB2* mRNA and HER2 protein abundance were moderately correlated (Pearson r = 0.62; R² = 0.39) (Figure 3A,B). *ERBB2*-amplified tumors had markedly higher *ERBB2* mRNA and HER2 protein abundance than tumors without *ERBB2* alterations (P < 0.001). Tumors harboring clinically interpreted *ERBB2* mutation events also showed higher ERBB2 mRNA expression (P < 0.001), whereas HER2 protein abundance was not significantly increased (P = 0.075) (Figure 3B). ERBB2 amplification was associated with both transcriptional and protein-level HER2 enrichment, whereas ERBB2 mutation was associated predominantly with increased ERBB2 transcript abundance. Using the positivity thresholds, RNA- and protein-based screening identified HER2-enriched populations that exceeded the prevalence defined by ERBB2 amplification alone. In the complete matched molecular subset (n = 1,130), ERBB2 copy-number amplification identified 3.2% of tumors, compared with 5.0%, 4.8%, 5.1% and 6.3% identified by ERBB2 mRNA, HER2 protein, the mRNA + protein model and the BG RNA signature, respectively (Figure 3C; Supplementary Fig. 10). Molecular HER2 positivity was enriched in tumor types with established HER2-positive subgroups, including breast and gastroesophageal cancers, but was also detected in other tumor types, including urothelial, prostate, colorectal and non-small cell lung cancers (Supplementary Table 8). Threshold sensitivity analyses changed the absolute prevalence but preserved the overall pattern across tumor types and screening modalities (Supplementary Fig. 11).

**Fig. 3.**
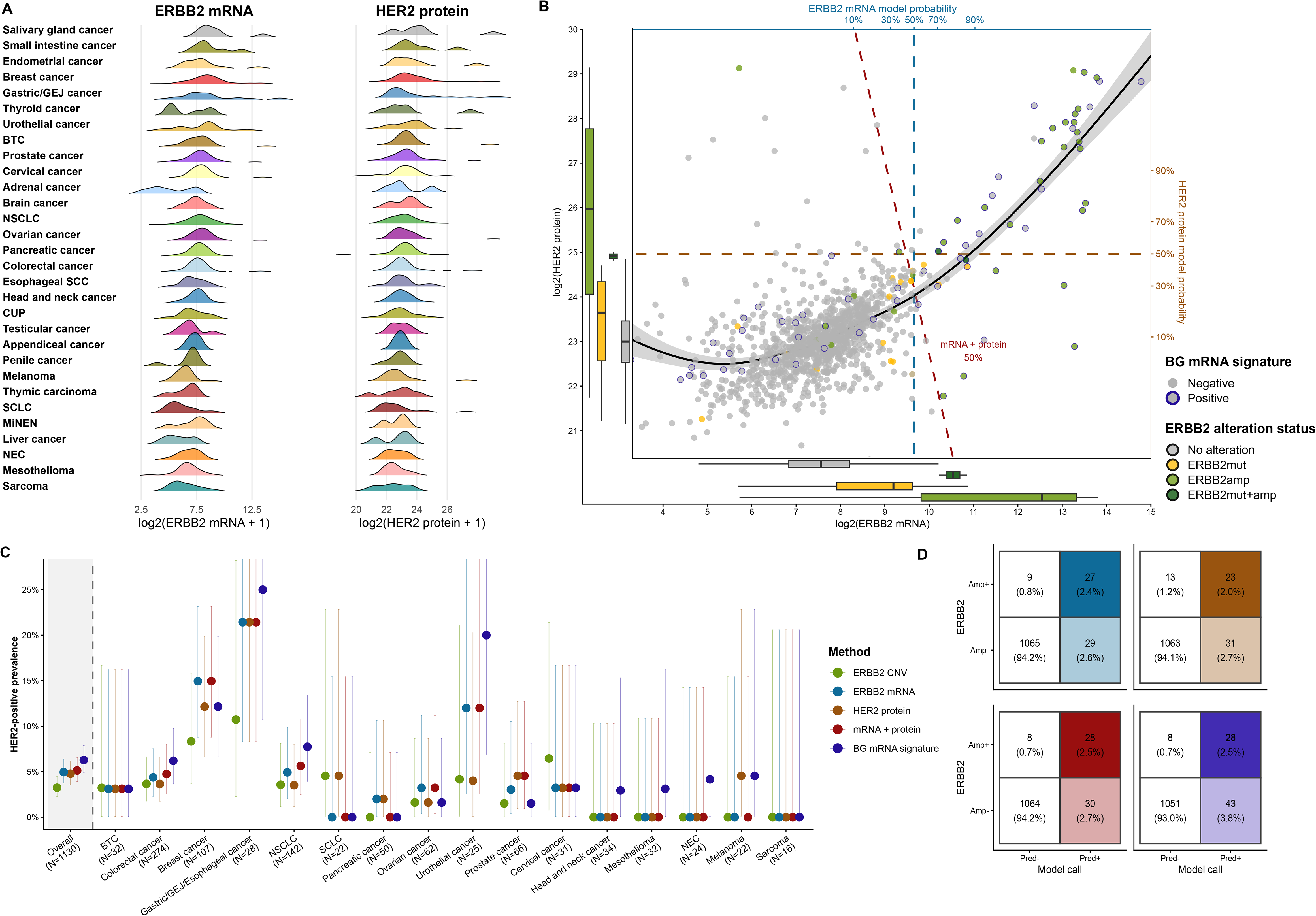
Pan-tumor implementation and overlap of molecular HER2 screening strategies. **A)** Distribution of ERBB2 mRNA expression and HER2 protein abundance across tumor types in the pan-tumor cohort. **B)** Relationship between ERBB2 mRNA expression and HER2 protein abundance in samples with paired measurements. The scatterplot shows log2-transformed ERBB2 mRNA and HER2 protein abundance; marginal boxplots summarize each analyte by ERBB2 alteration status. Dashed reference lines indicate the ERBB2 mRNA and HER2 protein positivity thresholds, and the dashed contour denotes the mRNA/protein model probability threshold of 0.50. Point outline indicates breast/gastroesophageal RNA signature status, and point fill indicates ERBB2 alteration status. **C)** Tumor-type-specific prevalence of HER2 positivity according to ERBB2 copy-number amplification, ERBB2 mRNA, HER2 protein abundance, the mRNA/protein model and the breast/gastroesophageal RNA signature. Points show prevalence estimates, and error bars indicate exact binomial 95% confidence intervals.

Overlap analyses confirmed that DNA-, RNA- and protein-based screens captured related but distinct HER2-positive populations. Of 36 ERBB2-amplified tumors, 27 (75.0%) were positive by ERBB2 mRNA, 23 (63.9%) by HER2 protein, 28 (77.8%) by the mRNA + protein model and 28 (77.8%) by the BG RNA signature (Figure 3D; Supplementary Fig. 12). Combining ERBB2 amplification with molecular screening expanded the positive population to 65 tumors for ERBB2 mRNA, 67 for HER2 protein, 66 for the mRNA + protein model and 79 for the BG RNA signature. Concordance was greatest between ERBB2 mRNA and the mRNA + protein model (Jaccard index, 0.87), whereas concordance between HER2 protein and ERBB2 amplification was lower (Jaccard index, 0.34) (Supplementary Fig. 13 and Supplementary Table 9). Notably, 29 of 56 ERBB2 mRNA-positive tumors (51.8%) and 43 of 71 BG RNA signature-positive tumors (60.6%) lacked ERBB2 amplification, demonstrating that expression-based screening identified substantial HER2-enriched populations outside the genomically amplified sub-group.

Because matched MS data were unavailable for most treated patients, we developed a pan-tumor RNA model trained to predict HER2 protein abundance, termed the PAN protein-informed RNA score (Supplementary Fig. 14). This score enabled protein-informed HER2 assessment from RNA-seq data alone and its subsequent evaluation in treated cohorts.

### 3. RNA-defined HER2 biology is associated with response to pan-tumor HER2-targeted therapy

Having shown that RNA- and protein-based screening identified HER2-enriched tumors beyond ERBB2 amplification, we next tested whether RNA-defined HER2 biology was associated with outcomes after HER2-targeted therapy. Because RNA-seq was available for all treated patients whereas matched MS-based HER2 protein quantification was available for only 13 patients, clinical outcome analyses focused on two RNA-based strategies: single-gene ERBB2 mRNA expression and a combined RNA signature requiring positivity for both the BG RNA signature and the PAN protein-informed RNA score.

The internal treated cohort comprised 51 patients with ERBB2-altered solid tumors treated through the CoPPO molecularly guided trial framework or the ProTarget phase II basket trial (Figure 1; Supplementary Table 10). Figure 1; Supplementary Table 10). The median number of prior systemic therapy lines was two. Treatment was dominated by T-DM1 and trastuzumab plus pertuzumab, with additional patients receiving trastuzumab, tyrosine kinase inhibition or trastuzumab deruxtecan. Most patients were treated on the basis of *ERBB2* amplification, while the remainder harbored *ERBB2* mutations. The cohort was enriched for non-small cell lung and colorectal cancers but included multiple additional tumor types.

At the specified thresholds, all six partial responses occurred in patients classified as positive by each RNA strategy (Figure 4A,B). HER2 protein showed a similar descriptive pattern among 13 patients with matched MS data (Supplementary Fig. 15). For *ERBB2* mRNA, partial response was observed in 6 of 28 RNA-positive patients and in 0 of 23 RNA-negative patients (21.4% versus 0.0%; Fisher’s exact P= 0.027) (Figure 4C, Supplementary Table 11). Similarly, for the combined RNA signature, partial response was observed in 6 of 20 signature-positive patients and in 0 of 31 signature-negative patients (30% versus 0.0%; Fisher’s exact P= 0.002). Because no objective responses occurred in either RNA-negative group, logistic regression was not performed for this endpoint owing to complete separation.

**Fig. 4.**
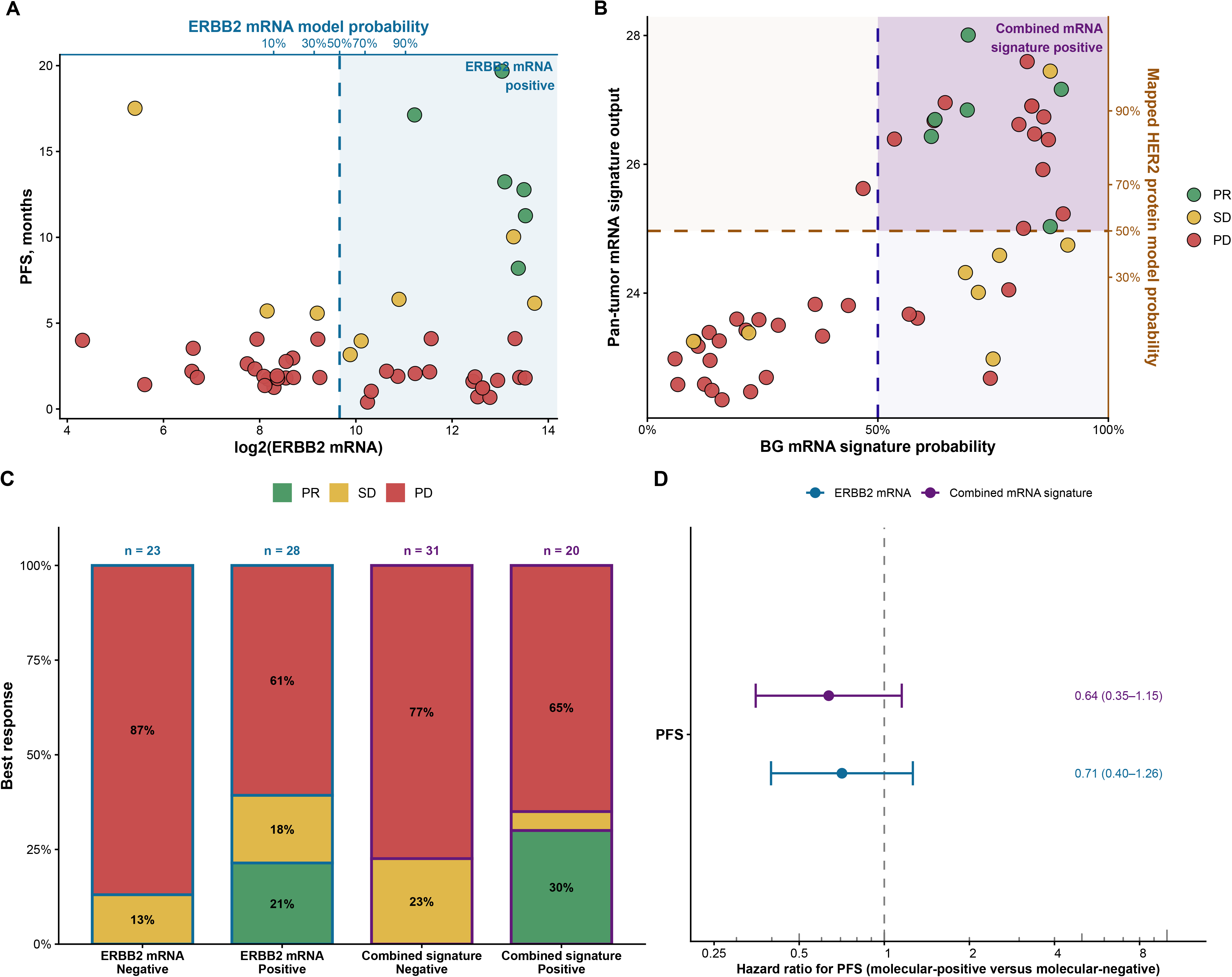
Transcriptional HER2 profiling and benefit from HER2-targeted therapy in the discovery treated cohort. **A)** ERBB2 mRNA expression and progression-free survival (PFS) in patients receiving HER2-targeted therapy. Points represent individual patients and are colored by best response. The dashed vertical line indicates the prespecified ERBB2 mRNA positivity threshold, and the shaded region denotes ERBB2 mRNA-positive disease. The upper x axis shows the corresponding model-derived probability of HER2 positivity. **B)** Transcriptional HER2 signature space defined by the breast/gastroesophageal HER2 RNA signature probability and the pan-tumor protein-informed RNA signature score. Dashed lines indicate the prespecified positivity thresholds for each signature, and the shaded upper-right quadrant denotes combined RNA signature-positive disease. Points represent individual patients and are colored by best response to HER2-targeted therapy. **C)** Best overall response to HER2-targeted therapy according to ERBB2 mRNA status and combined RNA signature status. Stacked bars show the proportion of patients with progressive disease red (PD), stable disease yellow (SD) or objective response green (partial or complete response; PR/CR) in molecular-negative and molecular-positive groups; sample sizes are indicated above each bar.FD) Association of ERBB2 mRNA positivity and combined RNA signature positivity with progression-free survival (PFS). Points indicate hazard ratios for molecular-positive versus molecular-negative disease from univariable Cox proportional-hazards models, and horizontal bars indicate 95% confidence intervals. The dashed vertical line denotes no difference in risk (hazard ratio = 1). **D)** Association of ERBB2 mRNA positivity and combined RNA signature positivity with progression-free survival (PFS). Points indicate hazard ratios for molecular-positive versus molecular-negative disease from univariable Cox proportional-hazards models, and horizontal bars indicate 95% confidence intervals. The dashed vertical line denotes no difference in risk (hazard ratio = 1).

Clinical benefit showed the same direction of effect (Figure 4C). For *ERBB2* mRNA, clinical benefit was observed in 11 of 28 RNA-positive patients in comparison to 3 of 23 RNA-negative patients (39.3% versus 13.0%; Fisher’s exact P= 0.058). For the combined RNA signature, clinical benefit was observed in 7 of 20 signature-positive patients compared with 7 of 31 signature-negative patients (35.0% versus 22.6%; Fisher’s exact P = 0.35).

RNA positivity was directionally associated with longer progression-free survival (PFS) (Figure 4D; Supplementary Fig. 16 and Supplementary Table 11). *ERBB2* mRNA positivity was associated with a PFS hazard ratio of 0.71 (95% CI 0.40–1.26; P= 0.242), while the combined RNA signature showed a directionally favorable association with PFS (HR 0.64, 95% CI 0.35–1.15; P= 0.136). Neither RNA-based strategy was associated with improved overall survival (OS) in this internal cohort (Supplementary Fig. 17 and Supplementary Table 11).

In this genomically selected, heterogeneous cohort, RNA-positive tumors accounted for all objective responses, whereas associations with clinical benefit, PFS and OS were imprecise or not statistically significant. Because the two RNA approaches performed similarly, these findings motivated external evaluation.

### 4. External validation confirms that RNA refines HER2 treatment selection beyond amplification

We next evaluated the RNA-based HER2 screening framework in an independent pan-tumor cohort (NCT01855477) with matched ERBB2 genomic annotation and RNA-seq data, including clinical treatment and outcome data for patients subsequently treated with HER2-targeted therapy^16^.

#### 4.1 RNA-based screening identifies HER2-enriched tumors beyond ERBB2 amplification

The external molecular screening cohort comprised 3,898 samples from 3,697 unique patients, representing 49 tumor types (Figure 1; Supplementary Tables 12 and 13). *ERBB2* mRNA expression was highest in amplified tumors, whereas tumors with *ERBB2* mutations alone showed modest enrichment relative to tumors without an interpreted *ERBB2* alteration (Figure 5A).

**Fig. 5.**
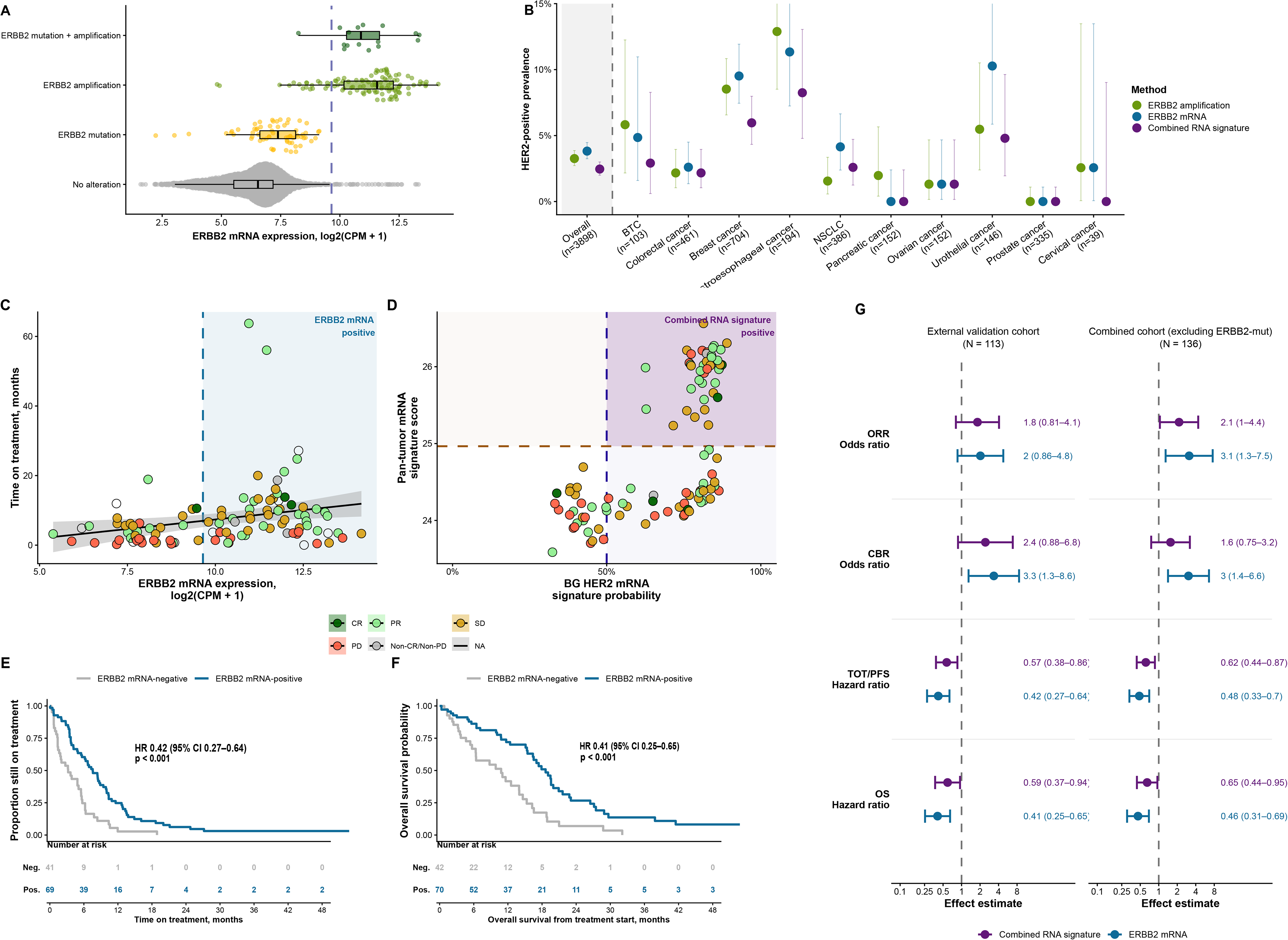
External validation of RNA-based HER2 screening and treatment selection. **A)** ERBB2 mRNA expression according to interpreted ERBB2 driver status in the external validation cohort. Violin plots show the distribution of ERBB2 expression, boxplots indicate the median and interquartile range, and points represent individual tumors. The dashed line denotes the prespecified ERBB2 mRNA positivity threshold. **B)** Prevalence of HER2 positivity by ERBB2 amplification, ERBB2 mRNA and the combined RNA signature. “Overall” represents the entire external screening cohort (3,898 samples); the remaining estimates represent selected cancer types, with sample sizes indicated. Points show prevalence estimates and error bars indicate 95% confidence intervals. **C)** ERBB2 mRNA expression and time on HER2-targeted treatment in the external treated validation cohort, including tumors with ERBB2 mutation drivers. Points represent individual treatment episodes with a documented treatment end and are colored by first radiological response. The dashed vertical line indicates the ERBB2 mRNA positivity threshold, and the shaded region denotes ERBB2 mRNA-positive disease. The black line shows the fitted linear trend with a 95% confidence interval. **D)** Treated validation samples plotted in breast/gastroesophageal HER2 RNA signature and pan-tumor protein-informed RNA signature space. Dashed lines indicate the prespecified positivity thresholds, and the shaded upper-right quadrant denotes combined RNA signature-positive disease. Points represent individual patients and are colored by first radiological response. **E)** Kaplan–Meier analysis of time on treatment according to ERBB2 mRNA status. Curves show the proportion of patients remaining on treatment over time, with the number at risk shown below the plot. The hazard ratio and P value were estimated using a Cox proportional hazards model. **F)** Kaplan–Meier analysis of overall survival from initiation of HER2-targeted therapy according to ERBB2 mRNA status. Curves show overall survival probability over time, with the number at risk shown below the plot. The hazard ratio and P value were estimated using a Cox proportional hazards model. BG, breast/gastroesophageal; CI, confidence interval; OS, overall survival. **G)** Forest plots summarizing associations between RNA-based HER2 molecular screening and clinical outcomes. The left panel shows the entire external validation cohort, including all ERBB2 driver categories (102 response, 110 ToT and 112 OS episodes); the right panel shows the combined internal and external HER2-treated cohort after exclusion of tumors with ERBB2 mutation. Molecular-positive versus molecular-negative cases were classified using ERBB2 mRNA expression (blue) or the combined RNA signature (purple). Odds ratios are shown for objective response rate (ORR; complete or partial response) and clinical benefit rate (CBR), and hazard ratios are shown for time on treatment/progression-free survival (TOT/PFS) and overall survival (OS). Points indicate univariable effect estimates and horizontal bars indicate 95% confidence intervals. The dashed vertical line denotes the null value (odds ratio or hazard ratio = 1)

At the sample level, *ERBB2* amplification was identified in 127 of 3,898 samples (3.3%), compared with 96 (2.5%) classified as positive by the combined RNA signature (Figure 5B). The two approaches identified overlapping but distinct populations: 74 samples were positive by both, while the combined RNA signature identified 22 additional samples without *ERBB2* amplification (Supplementary Figs. 18 and 19).

Tumor-type-specific prevalence patterns were consistent with established HER2 biology, with the highest HER2-positive fractions observed in breast and gastroesophageal cancers, while smaller RNA-positive subsets were also detected in selected non-breast and non-gastroesophageal tumors, including colorectal, biliary tract, non-small cell lung and urothelial cancers.

#### 4.2 RNA positivity enriches for benefit beyond *ERBB2* amplification

We then evaluated whether RNA-based HER2 positivity was associated with clinical benefit among patients treated with HER2-targeted therapy after molecular profiling. The treated validation cohort comprised 102 unique patients contributing 113 HER2-targeted treatment episodes, as individual patients could receive more than one line of HER2-targeted therapy after molecular profiling (Supplementary Table 14). At the treatment-episode level, breast cancer was most frequent (n=67), followed by gastroesophageal cancers (n=21), lung cancer (n=14) and colorectal cancer (n=9). Treatment consisted predominantly of trastuzumab plus pertuzumab (n = 57) or trastuzumab monotherapy (n = 54); ERBB2 amplification was present in 67 of 113 treatment episodes (59.3%).

Both RNA-based molecular screens were associated with favorable clinical outcomes in the validation cohort. ERBB2 mRNA positivity was associated with a higher clinical benefit rate (55/65 [84.6%] vs 23/37 [62.2%]; OR 3.35, 95% CI 1.30–8.62; P = 0.015), longer time on treatment (HR 0.42, 95% CI 0.27–0.64; P < 0.001) and longer overall survival (HR 0.41, 95% CI 0.25–0.65; P < 0.001). Objective response was numerically more frequent among ERBB2 mRNA-positive than mRNA-negative cases (30/65 [46.2%] vs 11/37 [29.7%]), although this difference was not statistically significant (OR 2.03, 95% CI 0.86–4.77; P = 0.142). The combined RNA signature showed a directionally consistent time-to-event pattern, with longer time on treatment (HR 0.57, 95% CI 0.38–0.86; P = 0.007) and overall survival (HR 0.59, 95% CI 0.37–0.94; P = 0.028) among signature-positive cases (Figure 5C–G; Supplementary Table 15 and Supplementary Fig. 20). To compare the outcome associations of RNA-based HER2 assessment and copy number, we evaluated continuous ERBB2 copy number and ERBB2 mRNA expression, standardized per one-standard-deviation increase, in the external HER2-treated validation cohort. Higher ERBB2 mRNA expression was associated with clinical benefit (OR 1.75, 95% CI 1.10–2.87; P = 0.020), longer time on treatment (HR 0.65, 95% CI 0.52–0.81; P < 0.001) and longer overall survival (HR 0.65, 95% CI 0.51–0.84; P < 0.001). ERBB2 copy number was not significantly associated with these endpoints (Supplementary Table 16 and Fig. 21).

Finally, pooled analyses across the internal and validation HER2-treated cohorts showed that the association between RNA-based HER2 positivity and treatment benefit was not confined to breast or gastroesophageal cancers and was observed across HER2-targeted treatment classes, although interaction testing was limited by sample size (Supplementary Fig. 22). In pooled analyses excluding ERBB2-mutated tumors (n = 136), both RNA strategies were associated with higher odds of objective response and more favorable time-to-event outcomes. ERBB2 mRNA positivity was additionally associated with clinical benefit (OR 3.0, 95% CI 1.4–6.6), as well as favorable time on treatment/progression-free survival (HR 0.48, 95% CI 0.33–0.70) and overall survival (HR 0.46, 95% CI 0.31–0.69) (Figure 5G; Supplementary Table 17).

Together, these external analyses suggest that RNA-based HER2 assessment provides information complementary to *ERBB2* amplification, identifying additional RNA-positive tumors during molecular screening and stratifying HER2 expression within amplified disease.

### 5. Spatial proteomics captures HER2 protein heterogeneity beyond bulk molecular profiling

Bulk DNA-, RNA- and protein-based measurements identified HER2-enriched tumors and enriched for benefit from HER2-targeted therapy, but these assays collapse spatially heterogeneous tissue into a single sample-level measurement. This is particularly relevant for HER2, where clinical IHC scoring integrates both staining intensity and the fraction of tumor cells showing membranous expression. We therefore applied tile-level deep visual proteomics (TileDVP), a morphology-guided spatial proteomic workflow, to quantify HER2 protein abundance across morphology-defined tumor regions while preserving intratumoral distribution.

We first benchmarked TileDVP in a breast (n = 46) and gastroesophageal (n = 33) reference cohort, comprising 1,362 and 955 tile-level MS measurements, respectively, and using the same clinical IHC/FISH categories as the reference standard (Supplementary Table 18). Each specimen was partitioned into morphology-defined regions for microdissection and proteomic analysis, generating spatially resolved HER2 measurements rather than a single bulk readout (Figure 6A). Across breast and gastroesophageal cancer samples, TileDVP achieved comparable analytical depth, with medians of 27,321 and 25,646 quantified peptides and 4,552 and 4,514 quantified proteins per tile, respectively (Supplementary Fig. 23).

**Fig 6.**
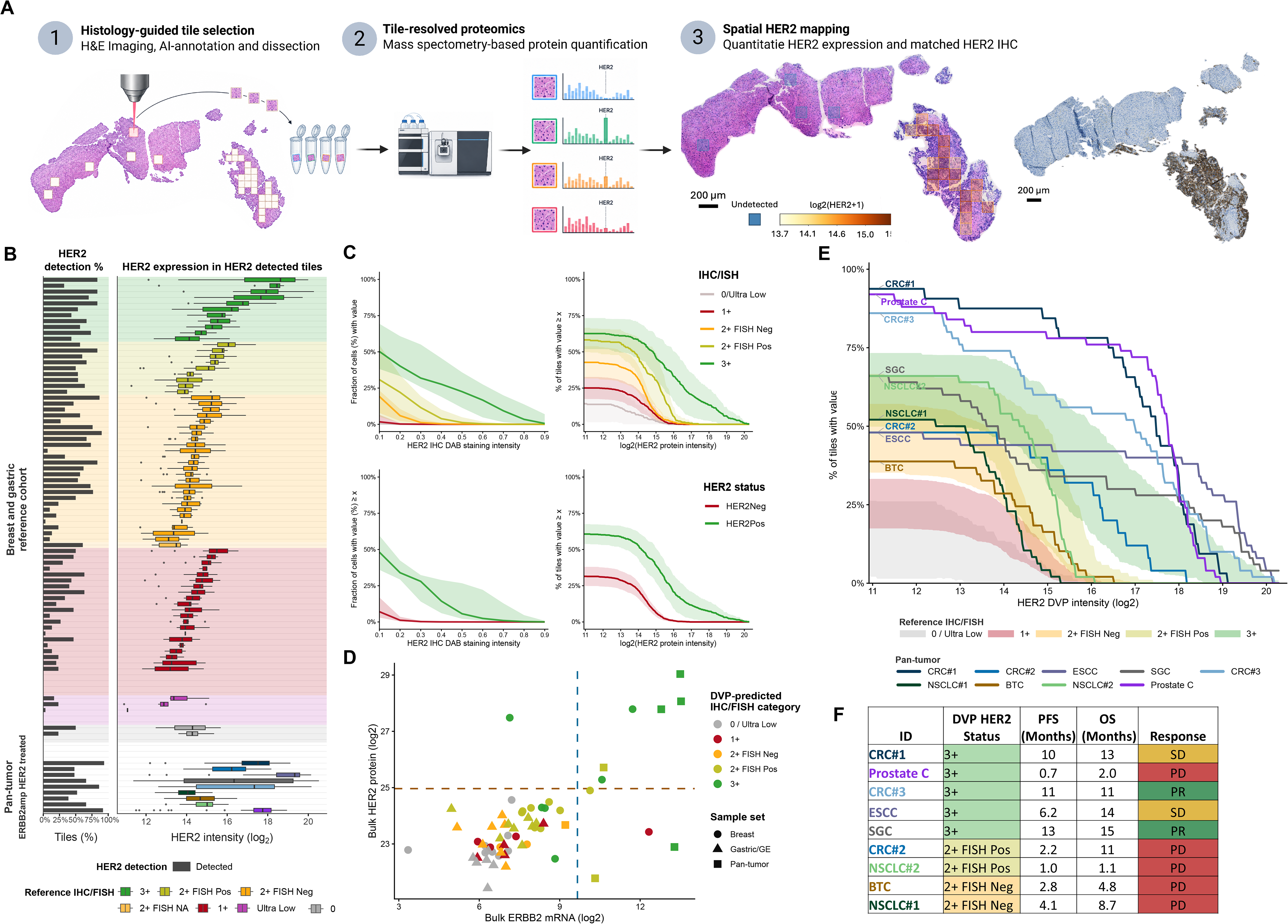
Spatial proteomics captures HER2 heterogeneity and identifies HER2-enriched profiles below bulk positivity thresholds. **A)** Tile-level deep visual proteomics (TileDVP) workflow, comprising histology-guided tile selection and laser microdissection, mass spectrometry-based protein quantification and spatial HER2 mapping alongside matched HER2 immunohistochemistry (IHC). **B)** Slide-level HER2 detection and abundance in the breast and gastroesophageal reference cohort and ERBB2-amplified, HER2-treated pan-tumor cohort. Bars indicate the percentage of tiles with detected HER2. Boxplots summarize HER2 intensity among detected tiles, with medians, interquartile ranges and whiskers extending to the most extreme values within 1.5 interquartile ranges. Reference slides are grouped by clinical IHC/FISH category. **C)** HER2 distribution curves stratified by clinical HER2 status. The upper row shows distributions across IHC/FISH categories, while the lower row compares HER2-positive and HER2-negative tumors. Left, digital IHC showing the median percentage of cells exceeding each DAB staining-intensity threshold, with bootstrap 95% confidence intervals. Right, DVP showing the mean percentage of sampled tiles exceeding each HER2 intensity threshold, with shaded pointwise intervals representing the mean ± 1.96 standard errors across slides. **D)** Bulk ERBB2 mRNA and HER2 protein abundance colored by DVP-assigned IHC/FISH category. Reference slides were classified by leave-one-slide-out analysis; pan-tumor slides were assigned using the full reference cohort. Shapes indicate sample cohort, and dashed lines mark the respective bulk positivity thresholds. **E)** Individual pan-tumor HER2 distribution curves overlaid on the breast/gastroesophageal reference intervals. Curves show the percentage of all sampled tiles exceeding each HER2 intensity threshold, retaining non-detected tiles in the denominator. **F)** DVP-assigned HER2 profiles and clinical outcomes in the nine ERBB2-amplified, HER2-treated pan-tumor cases. DVP categories indicate similarity to clinical IHC/FISH reference profiles and do not constitute measured IHC or FISH results. PFS, progression-free survival; OS, overall survival; PD, progressive disease; SD, stable disease; PR, partial response.

#### 5.1 Spatial proteomics resolves HER2 gradients across clinical IHC/FISH categories

Among 78 reference samples with resolved IHC/FISH status, the median proportion of tiles with detectable HER2 increased across clinical categories, from 10.0% in IHC0/ultra-low tumors to 23.3% in IHC1+, 50.0% in IHC2+/FISH-negative, 56.7% in IHC2+/FISH-positive and 66.7% in IHC3+ tumors (Supplementary Table 19). Sample-level median HER2 abundance among detected tiles likewise increased across these categories, supporting concordance between spatial proteomic HER2 measurements and clinical HER2 classification (Figure 6B; Supplementary Table 19). When represented as distribution curves, TileDVP-derived HER2 profiles recapitulated the expected ordering of clinical IHC/FISH categories while retaining quantitative information on regional HER2 heterogeneity. HER2-positive and HER2-negative tumors also exhibited pronounced separation, qualitatively resembling the distinction observed with digital IHC staining (Figure 6C).

Leave-one-slide-out (LOO) DVP assignments spanned the clinical IHC/FISH categories in the breast and gastroesophageal reference cohort (Supplementary Fig. 24). In breast cancer, agreement between LOO DVP and central pathology was 32.6% for IHC/FISH categories and 76.1% for binary HER2 status, compared with 39.1% and 84.8%, respectively, between routine and central pathology (Supplementary Fig. 25; Supplementary Table 20). DVP identified HER2-positive-like spatial profiles in 18 reference slides below both bulk ERBB2 mRNA and HER2 protein positivity thresholds, including 15 with IHC2+/FISH-positive-like assignments (Fig. 6D). Among clinically HER2-positive tumors classified as negative by bulk ERBB2 mRNA or protein, DVP assigned HER2-positive-like profiles to 7 of 11 and 6 of 11, respectively (Supplementary Fig. 26). Together, these findings suggest that spatial profiling can reveal regional HER2 enrichment not captured by bulk positivity thresholds.

#### 5.2 Spatial HER2 profiles in HER2-treated pan-tumor cases

Finally, we explored TileDVP-derived HER2 profiles in nine ERBB2-amplified pan-tumor patients treated with HER2-targeted therapy, comprising 404 tile-level MS measurements. Tile-level HER2 abundance distributions were compared with the breast/gastroesophageal reference curves, and each tumor was assigned to the nearest IHC0/ultra-low-like, IHC1+-like, IHC2+/FISH-negative-like, IHC2+/FISH-positive-like or IHC3+-like spatial HER2 profile (Figure 6E). Two tumors were assigned IHC2+/FISH-negative-like profiles, two IHC2+/FISH-positive-like profiles and five IHC3+-like profiles (Figure 6F). In this exploratory subset, clinical benefit occurred only among tumors with IHC3+-like spatial HER2 profiles. Given the small and heterogeneous cohort, these findings are descriptive and do not establish predictive treatment-selection utility.

## Discussion

Our findings support a multilayer framework for pan-tumor HER2 assessment. ERBB2 amplification identifies a highly specific genomic subgroup, RNA expression captures transcriptionally active HER2 biology, and proteomic measurements quantify target abundance. RNA- and protein-based screening identified HER2-enriched tumors beyond amplification, while RNA-defined HER2 biology was associated with outcomes following HER2-targeted therapy. Spatial proteomics further provided a distribution-aware protein readout that recapitulated clinical HER2 gradients while retaining regional heterogeneity. This distinction may become increasingly relevant as HER2-targeted antibody-drug conjugates expand across tumor types, where target abundance and spatial distribution may contribute information beyond copy-number status alone.

Clinically, ERBB2 amplification remains highly specific but may miss RNA- or protein-enriched tumors and does not distinguish expression levels within amplified disease. RNA-based assessment therefore provides a scalable refinement within existing molecular-profiling workflows, whereas spatial proteomics remains a complementary translational approach. Importantly, the treated cohorts were selected primarily on the basis of known ERBB2 genomic alterations. The present outcome data therefore do not establish benefit from HER2-targeted therapy in RNA-positive tumors lacking ERBB2 amplification or mutation. Prospective studies are required to determine whether these tumors represent a therapeutically targetable HER2-enriched population, particularly in the context of antibody-drug conjugates, where protein expression, bystander effects and intratumoral heterogeneity may influence activity ^5,17,18^.

ERBB2 mRNA performed strongly across analyses and offers a simple, interpretable RNA readout. In the external treated cohort, higher continuous ERBB2 mRNA expression was associated with clinical benefit, longer time on treatment and overall survival, whereas corresponding associations with ERBB2 copy number were not statistically significant. The combined RNA signature showed overlapping performance without consistently outperforming ERBB2 mRNA, although broader transcriptional programs may provide complementary information. ERBB2 mRNA therefore represents the most immediately scalable RNA-based marker.

Spatial proteomics addresses tissue averaging by retaining regional target abundance. Across 88 samples and 2,721 tile-level measurements, TileDVP profiles increased across clinical IHC/FISH categories and identified selected HER2-positive tumors missed by bulk RNA or proteomics. Clinical benefit occurred only among tumors with IHC3+-like profiles in the small exploratory pan-tumor subset, warranting further evaluation rather than establishing clinical utility. The ability to quantify multiple targets from the same morphology-defined regions may additionally provide a tissue-sparing complement to sequential single-marker assessment, although target-specific localization, analytical cutoffs and clinical actionability require validation.

This study has limitations. First, the internal treated cohort was modest in size and included heterogeneous tumor types, *ERBB2* alteration classes and HER2-targeted regimens. Although external validation supported the clinical relevance of RNA-based HER2 assessment, subgroup and interaction analyses were limited by sample size. Second, the treated cohorts were not randomized biomarker-stratified trials; therefore, these analyses establish associations with outcomes among treated patients rather than a formal biomarker-treatment interaction or predictive treatment effect. Treatment selection was influenced by genomic eligibility, clinical-trial availability and physician decision-making. Third, clinical outcome analyses were primarily anchored in genomically selected treated populations, limiting inference for RNA-positive tumors without canonical *ERBB2* alterations. Fourth, MS-based HER2 protein quantification was available for only a subset of patients, limiting direct evaluation of bulk protein as a treatment-selection biomarker. Fifth, while RNA-based thresholds were defined before application from the reference breast and gastric framework and externally applied, further clinical calibration will be needed before implementation across platforms and institutions. Finally, TileDVP analyses were exploratory, particularly in the pan-tumor treated cohort. TileDVP sampled selected morphology-defined regions rather than exhaustively mapping each tumor and may therefore not capture the full spatial heterogeneity of a specimen. Larger prospective studies with standardized sampling, classification rules and clinical endpoints are required.

In conclusion, *ERBB2* amplification does not fully capture pan-tumor HER2 biology. RNA-based assessment provides a scalable expression-level complement to copy-number profiling, while proteomic and spatial approaches quantify target abundance and regional heterogeneity. Prospective biomarker-driven studies are needed to determine whether RNA-positive or spatially HER2-enriched tumors without canonical *ERBB2* alterations benefit from HER2-targeted therapy and how these complementary molecular layers should inform treatment selection.

## Online Methods

### 1. Study design and cohorts

This study integrated clinical, genomic, transcriptomic, proteomic and spatial proteomic data to evaluate HER2 assessment beyond *ERBB2* copy-number amplification. The analysis was structured around five components: clinical HER2 benchmarking in breast and gastroesophageal cancers, pan-tumor molecular HER2 screening, clinical outcome analysis in HER2-treated pan-tumor patients, external validation in an independent pan-tumor cohort and spatial HER2 profiling using deep visual proteomics.

The Copenhagen Prospective Personalized Oncology (CoPPO) cohort included patients with advanced solid tumors referred for comprehensive molecular profiling after exhaustion of standard treatment options^19,20^. Patients underwent molecular characterization to support potential enrollment in molecularly guided early-phase or last-line clinical trials. The cohort is therefore enriched for patients with advanced, treatment-refractory disease and represents a selected population with respect to fitness, prior treatment exposure and availability of tumor tissue.

Patients with *ERBB2* genomic alterations treated with HER2-targeted therapy in the internal treated cohort were enrolled in molecularly guided clinical trials within the CoPPO framework. Additional HER2 treated patients from the ProTarget trial were also included in the HER2 treated cohort^21^. ProTarget is a national, non-randomized, open-label, phase II basket trial evaluating EMA-approved targeted therapies outside their labeled indications in patients with advanced cancers harboring actionable genomic alterations. Eligibility required measurable disease according to RECIST v1.1, ECOG performance status 0–2 and confirmation of a drug–variant match by the Danish National Molecular Tumor Board. Key exclusions included tumor types already with approved marketing authorization for the matched study drug and known resistance-associated alterations. Treatment followed EMA label dosing and schedules, with protocol-defined modifications as needed. Response assessments were performed every eight weeks during the first 24 weeks and every 12 weeks thereafter.

For external validation, an independent pan-tumor cohort with matched *ERBB2* genomic annotation and RNA-seq data was used. This cohort was analyzed both as a molecular screening cohort and, among patients subsequently treated with HER2-targeted therapy after profiling, as an external treated validation cohort.

### 2. Clinical HER2 reference cohort

#### 2.1 Central HER2 reference assessment

To establish a clinically anchored HER2 reference framework, breast cancer and gastroesophageal adenocarcinoma samples underwent centralized HER2 assessment with matched molecular profiling. The central IHC/FISH classification served as the reference endpoint for benchmarking *ERBB2* copy-number amplification, *ERBB2* mRNA expression, HER2 protein abundance and molecular classifiers.

The clinical reference cohort comprised 229 tumors (181 breast and 48 gastroesophageal). Central HER2 IHC scores were available for 177 breast and 46 gastroesophageal tumors, and FISH results for 169 and 46, respectively; paired IHC/FISH results were available for 209 tumors (165 breast and 44 gastroesophageal). HER2 IHC was performed using the HercepTest mAb pharmDx assay (Dako Omnis, GE001; Agilent Technologies) according to the manufacturer’s instructions. IHC staining and FISH results were interpreted according to the applicable ASCO/CAP criteria for breast and gastroesophageal adenocarcinoma. Breast cancer cases were assessed by two breast pathologists and gastroesophageal cancer cases by two gastrointestinal pathologists, all blinded to molecular profiling results. Semi-quantitative HER2 IHC categories were recorded as 0, ultra-low, 1+, 2+ and 3+. Staining heterogeneity, predominant staining localization and background staining were also recorded. Staining localization was categorized as membranous, cytoplasmic or nuclear where applicable.

For binary analyses, clinically HER2-positive tumors were defined as IHC3+ or IHC2+ with *ERBB2* amplification by FISH. Tumors scored as IHC0, ultra-low, IHC1+ or IHC2+ without *ERBB2* amplification were classified as HER2 non-positive. Cases with unresolved equivocal status were excluded from analyses requiring binary clinical HER2 status.

#### 2.2 Validation against historical HER2 status on the same tissue block

Historical diagnostic HER2 results were retrieved from pathology records where available, restricting comparisons to cases assessed on the same tissue block as the central study review. Historical results included IHC scores and available in situ hybridization amplification results. Historical IHC staining had been performed using either the PATHWAY anti-HER-2/neu (4B5) assay (Ventana) or the original HercepTest assay (Dako; polyclonal rabbit antibody A0485), whereas central study assessment used the HercepTest mAb pharmDx assay (Dako Omnis, GE001; Agilent Technologies). Agreement in binary clinical HER2 status was evaluated between historical and central assessments (Supplementary Fig. 27). The central IHC/FISH classification was retained as the reference for all primary model-training and benchmarking analyses.

### 3. Digital pathology assessment of HER2 staining

Digital pathology analysis was performed using QuPath. HER2 DAB staining intensity was quantified at the cell or tissue level, with separate assessment of membranous and cytoplasmic staining where feasible. Staining intensity was scaled from 0 to 1. A DAB intensity threshold of 0.10 was used to distinguish true HER2 staining from faint background signal.

To characterize HER2 staining burden continuously, precomputed digital pathology summaries were used to calculate the fraction of tissue exceeding a series of HER2 DAB intensity thresholds. These values generated a per-tumor staining-burden profile describing how the fraction of HER2-positive tissue changed as the required staining intensity increased. Tumors were stratified by IHC/FISH-derived reference category, and median staining-burden curves were generated for each category. Uncertainty bands were calculated using bootstrap resampling of the median with 1,000 resamples to generate pointwise 95% confidence intervals.

### 4. Genomic profiling and *ERBB2* driver annotation

Genomic profiling was performed within the Copenhagen Prospective Personalized Oncology (CoPPO) framework using prospectively collected tumor biopsies and matched germline DNA where available. In the standard CoPPO biopsy workflow, tumor material was collected for parallel genomic and transcriptomic profiling, with additional tissue used for histological assessment. The genomic profiling strategy evolved during the study period and included tumor/normal whole-exome sequencing (WES), whole-genome sequencing (WGS), SNP-array-based copy-number profiling and, in selected cases, targeted genomic sequencing, according to the clinical profiling workflow in use at the time of analysis. Sequencing, variant annotation and clinical interpretation followed the contemporaneous CoPPO procedures as previously described ^19,20^.

Somatic variants were identified by comparison with matched germline DNA where available, followed by variant annotation and clinical interpretation. Genome-wide copy-number alterations were assessed using the genomic copy-number data available from the contemporaneous profiling workflow. For quantitative copy-number analyses, ERBB2 amplification was defined as an estimated ERBB2 copy number of ≥5 copies. Where fresh prospectively collected tumor material was unavailable, archived FFPE tumor tissue or plasma-derived circulating tumor DNA could be used for targeted genomic profiling.

For the present study, ERBB2 genomic status was derived from the interpreted clinical genomic annotation and harmonized into four categories: ERBB2 amplification, ERBB2 mutation, concurrent ERBB2 amplification and mutation, or no interpreted ERBB2 driver event. ERBB2 amplification was used as the primary genomic comparator for RNA- and protein-based HER2 screening analyses. ERBB2 mutation status was used to identify mutation-driven cases and to define sensitivity analyses in HER2-treated cohorts.

### 5. RNA-seq preprocessing and *ERBB2* mRNA expression

Bulk RNA-seq was performed using an Illumina-based workflow as previously described ^19,20^. Gene-level expression was quantified as counts per million. A post-normalized RNA-seq matrix was used for RNA-based HER2 modelling. The matrix comprised 19,368 RefSeq-aligned transcripts with detectable expression in the contributing internal and reference datasets. *ERBB2* mRNA expression was analyzed on the log2CPM scale; where raw CPM values were used, *ERBB2* expression was transformed as log2(CPM + 1).

Sample identifiers were harmonized across RNA-seq, clinical annotation, genomic and proteomic datasets. For single-gene analyses, *ERBB2* mRNA expression was evaluated as a continuous variable and as a binary screening variable.

### 6. RNA-seq normalization and robustness analyses

#### 6.1 Rationale

To evaluate whether technical or compositional factors could bias bulk HER2 measurements sensitivity analyses were performed examining reference-gene normalization of *ERBB2* mRNA, biopsy-site adjustment and stromal-admixture adjustment. These analyses were used to test the robustness of the *ERBB2* mRNA–HER2 protein relationship and downstream inferences. Unadjusted measurements were retained as the primary analytic scale because the correction strategies did not materially alter the results and because aggressive correction risked over-adjusting biologically relevant disease-context signal.

#### 6.2 Reference-gene ratio normalization of *ERBB2* mRNA

Inspired by copy-number ratio reporting in in situ hybridization assays, *ERBB2* transcript abundance was normalized to selected reference transcripts to assess whether ratio-normalized mRNA measures improved concordance with clinical HER2 status or quantitative HER2 protein abundance (Supplementary Fig. 28). For each sample, *ERBB2* mRNA was divided by GAPDH, CHKB and MRPS24 expression to generate *ERBB2*/GAPDH, *ERBB2*/CHKB and *ERBB2*/MRPS24 ratios. GAPDH was used as a conventional housekeeping transcript, whereas CHKB and MRPS24 were selected on the basis of low variance across the dataset.

Reference-gene suitability was evaluated by inspecting overall and cancer-type-stratified expression distributions and variance. Performance of ratio-normalized *ERBB2* measures was compared with unnormalized *ERBB2* mRNA by visual concordance with clinical HER2 calls and by agreement with quantitative HER2 protein abundance. Because ratio normalization did not provide consistent improvement, it was not adopted for primary analyses.

#### 6.3 Biopsy-site adjustment

To assess whether biopsy site systematically influenced bulk *ERBB2* mRNA or HER2 protein abundance, separate linear models were fitted for log2 *ERBB2* mRNA and log2 HER2 protein with biopsy site as a categorical predictor (Supplementary Figs. 29 and 30). Adjusted values were constructed using an intercept-plus-residual re-anchoring approach, subtracting the estimated biopsy-site effect while preserving the original measurement scale and anchoring values to a designated reference biopsy category where available. Removal of biopsy-site effects was evaluated by testing for residual association between adjusted values and biopsy site.

#### 6.4 Stromal-content adjustment

Because low tumor cellularity and stromal admixture can dilute bulk HER2 signal, two independent stromal proxies were calculated for each sample: the xCell StromaScore derived from bulk expression deconvolution and the ESTIMATE StromalScore after gene-symbol harmonization (Supplementary Fig. 31). To remove linear dependence on stromal content while preserving the original log2 *ERBB2* scale, *ERBB2* mRNA values were re-anchored to a common stromal reference defined as the cohort-median stromal score. Each sample was adjusted by the expected difference attributable to its stromal score relative to the cohort median. Adjustments were performed separately using xCell and ESTIMATE.

#### 6.5 Combined biopsy-site and stromal adjustment

Joint models including both biopsy site and stromal score were fitted to evaluate whether apparent biopsy-site effects were explained by stromal admixture (Supplementary Fig. 32). Adjusted values were re-anchored to a common reference condition, defined by the reference biopsy category and the cohort-median stromal score. Across biopsy-site-only, stroma-only and combined biopsy-site plus stromal adjustment strategies, the *ERBB2* mRNA–HER2 protein relationship and derived interpretations were qualitatively unchanged compared with unadjusted analyses. Because biopsy site is strongly correlated with cancer type and may also reflect true differences in tumor content and biology, uncorrected values were used for primary analyses and corrected values were reported only as sensitivity analyses.

### 7. Bulk HER2 proteomic quantification

#### 7.1 Peptide preparation

Protein fractions were processed using the protein aggregation capture protocol on a KingFisher™ Flex System (Thermo Scientific™) with MagReSyn® Hydroxyl beads (ReSyn Biosciences). The bead storage solution was replaced with 70% acetonitrile (ACN). KingFisher deep-well plates were prepared with either 1 mL of 95% ACN or 70% ethanol. Each sample received 100 µL of digestion solution containing 60 mM triethylammonium bicarbonate (TEAB), Lys-C and trypsin at an enzyme-to-substrate ratio of 1:500. Samples were brought to a volume of 300 µL with 100 mM Tris buffer (pH 8.0), followed by the addition of ACN to a final concentration of 70% (v/v). Beads were added at a protein-to-bead ratio of 1:2.

Protein aggregation was facilitated by two rounds of mixing, each comprising 1 min at medium speed followed by a 10-min rest. Subsequent washes were performed for 2.5 min with gentle mixing while the beads remained attached to the magnet. Digestion proceeded overnight at 37 °C using 100 cycles of 45-s agitation separated by 6-min pauses. Protease activity was quenched by adding trifluoroacetic acid (TFA) to a final concentration of 1% (v/v).

#### 7.2 LC-MS/MS analysis

Peptides were loaded onto Evotips Pure and analyzed by data-independent acquisition (DIA). For each analysis, 200 ng of peptides were partially eluted from the Evotips with <35% ACN and analyzed using an Evosep One LC system (Evosep Biosystems) coupled online to an Orbitrap Astral mass spectrometer (Thermo Fisher Scientific). Peptides were separated on an 8-cm PepSep column (150 µm inner diameter) packed with 1.5-µm Reprosil-Pur C18 particles (Dr Maisch), using the standard 21-min gradient method corresponding to 60 samples per day and a stainless-steel emitter (30 µm inner diameter). The mobile phases were 0.1% formic acid in LC–MS-grade water (buffer A) and 0.1% formic acid in ACN (buffer B).

Each DIA acquisition cycle comprised an MS1 survey scan acquired in the Orbitrap at a resolution of 240,000, with a normalized automatic gain control (AGC) target of 500% and a maximum injection time of 100 ms. Fragment ion scans were acquired with a maximum injection time of 5 ms using 200 windows of 3 Th spanning m/z 380–980. Higher-energy collisional dissociation (HCD) was performed at a normalized collision energy of 25%.

#### 7.3 Raw file analysis for proteomics data

Raw files were converted to mzML format using MSConvert from ProteoWizard^22^. The mzML files were analyzed using DIA-NN (v2.2) in a library-free workflow against a whole-human-proteome FASTA reference dated 25 March 2025. In silico spectral libraries were generated using the DIA-NN deep-learning predictor with tryptic cleavage after lysine or arginine, allowing up to two missed cleavages. Peptides of 7–52 amino acids, precursor m/z values of 300–1800 and charge states of 2–7 were considered. Methionine oxidation and protein N-terminal acetylation were specified as variable modifications, with up to two variable modifications per peptide. MS1 and MS2 mass tolerances were set to 15 ppm, and identifications were filtered at a q-value of 0.01. Peptidoform scoring, quantification matrix generation, XIC extraction, relaxed protein inference and smart profiling were enabled.

### 8. Molecular classifier development in the breast/gastroesophageal reference cohort

#### 8.1 Logistic regression models of clinical HER2 positivity

Molecular classifiers were developed in the breast/gastroesophageal reference cohort using clinical IHC/FISH-defined HER2 status as the binary endpoint. Logistic regression models were trained using: (i) log₂-transformed ERBB2 mRNA expression, (ii) log₂-transformed HER2 protein abundance and (iii) both measurements, termed the mRNA + protein model. Models were trained using five-fold cross-validation with class-probability output. Performance was summarized using cross-validated area under the receiver operating characteristic curve and operating characteristics at a probability threshold of 0.50, including sensitivity, specificity, positive predictive value, negative predictive value and accuracy. For downstream single-analyte screening, the continuous ERBB2 mRNA and HER2 protein thresholds were defined as the values corresponding to a predicted probability of 0.50 in the respective univariate logistic regression models.

#### 8.2 Breast/gastroesophageal RNA classifier of clinical HER2 positivity

A breast/gastroesophageal RNA signature (BG RNA signature) was developed to predict clinically defined HER2 positivity from bulk RNA expression. The training cohort comprised breast cancer and gastroesophageal adenocarcinoma samples with matched RNA-sequencing data and central clinical HER2 annotation. Clinical HER2 status was derived from the IHC/FISH reference classification and encoded as a binary endpoint. Samples without an assigned clinical HER2 label were excluded from model training.

To reduce dimensionality before model fitting, differential expression analysis was performed between clinically HER2-positive and HER2-negative tumors using limma. Linear models were fitted to the log2CPM expression matrix with clinical HER2 status as the explanatory variable, followed by empirical Bayes moderation with trend correction. Transcripts were ranked according to the absolute estimated log fold-change between HER2-positive and HER2-negative tumors, and the top 10,000 transcripts were retained as candidate predictors.

A random forest classifier was trained using the selected transcript set as input features and clinical HER2 status as the binary outcome. Model training was performed using the ranger implementation through caret with repeated five-fold cross-validation repeated three times. Class probabilities were estimated, and hyperparameters were tuned within cross-validation using area under the receiver operating characteristic curve as the optimization metric. The final model was refit on the full labeled breast/gastroesophageal training cohort using the optimal hyperparameters. For downstream analyses, the BG RNA signature output was the predicted probability of clinical HER2 positivity, with a threshold of ≥0.50 defining positivity.

#### 8.3 Classifier benchmarking against clinical HER2 status

For benchmarking in the breast/gastroesophageal reference cohort, clinical IHC/FISH-defined HER2 status was used as the reference endpoint. The evaluated approaches comprised ERBB2 copy-number amplification, ERBB2 mRNA, HER2 protein, the mRNA + protein model and the BG RNA signature. For probability-based models, samples were classified as positive at a predicted probability ≥0.50. ERBB2 copy-number amplification was analyzed as a binary predictor.

Classifier performance was assessed using cross-validated predicted probabilities where available. Discrimination was summarized by the area under the receiver operating characteristic curve. At the specified classification thresholds, operating characteristics included sensitivity, specificity, positive predictive value, negative predictive value, accuracy and the fraction of clinically HER2-positive tumors not captured by each molecular method.

### 9. Pan-tumor RNA model of HER2 protein abundance

Because clinically validated IHC/FISH criteria are not established across all solid tumor types, a second RNA-based model was developed in the pan-tumor internal cohort using quantitative HER2 protein abundance as the endpoint. The modelling cohort included samples with matched bulk RNA-seq and mass spectrometry-based HER2 protein quantification. The endpoint was continuous log2-transformed HER2 protein abundance.

RNA-seq profiles were restricted to samples with available HER2 protein measurements and linked to the corresponding clinical and proteomic annotation by sample identifier. To select informative transcript features, each transcript was tested for association with continuous HER2 protein abundance using limma. Linear models were fitted with log2 HER2 protein abundance as a continuous explanatory variable, and empirical Bayes moderation was applied. Transcripts were ranked by the moderated test statistic, and the top 10,000 transcripts were retained for model training.

A random forest regression model was trained to predict continuous HER2 protein abundance from the selected RNA features using ranger through caret. Model development used five-fold cross-validation repeated three times. Hyperparameters were tuned within cross-validation, and root mean squared error was used as the primary optimization metric. The final fitted model generated a continuous PAN protein-informed RNA score for each sample using RNA expression alone. Variable importance was estimated using permutation-based importance.

### 10. RNA-based HER2 screening signatures

For downstream screening and clinical outcome analyses, RNA-derived HER2 status was evaluated using two approaches. First, ERBB2 mRNA expression was dichotomized using the threshold of ≥ 9.647615 derived from the breast/gastroesophageal reference cohort. Second, a combined RNA signature was defined by joint positivity for the BG RNA signature and the PAN protein-informed RNA score.

Combined RNA signature positivity required a BG RNA signature probability ≥0.50 and a PAN protein-informed RNA score ≥24.96546. Samples with both model outputs available that did not meet both thresholds were classified as negative. Both component outputs were derived from RNA expression, so application of the combined RNA signature did not require measured protein abundance.

This dual-positive rule was used consistently in the internal cohort, external validation cohort and HER2-treated clinical outcome analyses.

#### 10.1 Sensitivity analysis of model development

To assess the robustness of our findings to potential information leakage during model development, we performed a sensitivity analysis in which RNA signature development was repeated with feature selection incorporated within cross-validation folds, rather than performed before resampling. Internal discriminatory performance was reassessed using out-of-fold predictions. Models trained on the full development cohorts were then applied to the same internal and external validation samples, using identical positivity thresholds (Supplementary Table 21). We compared HER2-positive prevalence, patient-level classification agreement, and associations with treatment outcomes, including objective response, clinical benefit, time on treatment, progression-free survival, and overall survival. Clinical endpoint definitions and censoring rules were held constant between analyses to isolate the effect of the alternative model-development procedure.

### 11. Pan-tumor molecular HER2 screening

The breast/gastroesophageal-derived HER2 screening framework was applied to the pan-tumor internal cohort. Each tumor was assigned binary HER2 screening calls for *ERBB2* copy-number amplification, *ERBB2* mRNA positivity, HER2 protein positivity, the combined RNA/protein logistic regression model and the breast/gastroesophageal mRNA signature. The combined RNA signature was evaluated as described above.

Tumor types were harmonized before analysis. Gastroesophageal and esophageal categories were grouped for tumor-type-specific prevalence estimates where appropriate. HER2-positive prevalence was calculated overall and by tumor type as the proportion of evaluable samples classified as positive by each method. Exact binomial 95% confidence intervals were calculated for prevalence estimates.

To assess the incremental yield of expression-based screening beyond *ERBB2* copy-number amplification, overlap with *ERBB2* amplification was defined as the intersection between a molecular method and *ERBB2* amplification, whereas combined yield was defined as positivity by either *ERBB2* amplification or the molecular method. Pairwise method concordance was quantified using the Jaccard index, calculated as the size of the intersection divided by the size of the union of the two positive sets.

Threshold sensitivity analyses were performed by varying positivity thresholds around the specified cut-points to assess whether overall prevalence patterns and tumor-type distributions were robust to alternative threshold choices.

### 12. Covariate-adjusted analysis of RNA and protein readouts

To evaluate whether bulk *ERBB2* mRNA and HER2 protein measurements were influenced by histopathological and cohort-level features beyond ordinal IHC/FISH category, expression values were analyzed in covariate-adjusted linear models. *ERBB2* mRNA and HER2 protein abundance were standardized to z scores and modelled separately. The full model included IHC/FISH category, cancer type, cytoplasmic staining and heterogeneous staining, together with interactions between IHC/FISH category and each covariate.

Signed average marginal effects were estimated for cancer type, cytoplasmic staining and heterogeneous staining. The relative contribution of each covariate block was quantified as the change in model R² between the full model and a reduced model omitting the covariate and its interaction with IHC/FISH category. Nested models were compared using analysis of variance.

### 13. HER2-treated cohort and clinical endpoints

The HER2-treated internal cohort included patients with *ERBB2*-altered solid tumors who received HER2-targeted therapy in molecularly guided clinical trials and had available RNA-seq data. Patients were treated with HER2-directed regimens including antibody-drug conjugates, dual-antibody therapy, trastuzumab-based therapy, tyrosine kinase inhibition or trastuzumab deruxtecan. Clinical characteristics, *ERBB2* alteration class, treatment regimen, prior systemic treatment exposure and tumor type were abstracted from trial records.

Clinical response was assessed according to RECIST v1.1 where measurable disease and response data were available. OR was defined as complete response or partial response versus stable disease, progressive disease or non-complete response/non-progressive disease. Clinical benefit was defined as complete response, partial response, stable disease or non-complete response/non-progressive disease versus progressive disease.

Progression-free survival was defined as time from initiation of HER2-targeted therapy to radiographic or clinical progression or death, whichever occurred first. OS was defined as time from initiation of HER2-targeted therapy to death from any cause. Patients without events were censored at last follow-up. Time on treatment was calculated from HER2-targeted treatment start date to treatment discontinuation or last known treatment date, according to available treatment records.

Because matched bulk HER2 protein quantification was available for only a subset of treated patients, clinical outcome analyses focused on two RNA-based methods: single-gene *ERBB2* mRNA positivity and the combined RNA signature. Analyses were performed using the specified positivity thresholds.

### 14. External validation cohort

#### 14.1 Cohort source and data access

The external validation cohort was derived from data made available by Hartwig Medical Foundation through the Hartwig Medical Database under Data Access Request DR-505 ^16^. The approved project was designed to validate RNA-seq-based HER2 signatures using Hartwig genomic, transcriptomic and clinical annotations. The requested data comprised pan-cancer metastatic samples with RNA-seq, matched somatic genomic analyses including copy-number data, core tumor and patient annotations, previous treatment data and post-biopsy treatment response data where available. Analyses were performed using derived molecular and clinical variables, and no original or raw licensed data are redistributed with this publication.

Hartwig RNA-sequencing expression data were imported as counts per million (CPM) and transformed as log2(CPM + 1), matching the expression scale used for model training. Gene identifiers were mapped to gene names, duplicate entries were averaged on the log-transformed scale, and the expression matrix was aligned to the predictor gene lists required by the trained models. Missing expression values were imputed using the median for the corresponding gene across Hartwig samples, whereas model-required genes absent from the matrix were assigned a value of zero.

#### 14.2 External validation screening analyses

The external validation cohort included patients with available *ERBB2* genomic annotation and RNA-seq data. The full cohort was used for molecular screening validation. *ERBB2* mRNA positivity and combined RNA signature positivity were assigned using the same thresholds as in the internal cohort.

Interpreted *ERBB2* genomic driver annotations were harmonized into four categories: no alteration, *ERBB2* mutation, *ERBB2* amplification and concurrent *ERBB2* mutation plus amplification. Tumors with *ERBB2* amplification or concurrent *ERBB2* mutation plus amplification were considered *ERBB2* amplification-positive for genomic screening comparisons.

For tumor-type-specific screening analyses, primary tumor labels were harmonized into predefined cancer-type groups, including biliary tract cancer, colorectal cancer, breast cancer, gastric/gastroesophageal junction/esophageal cancer, non-small-cell lung cancer, pancreatic cancer, ovarian cancer, urothelial cancer, prostate cancer and cervical cancer. Tumor-type-specific prevalence estimates were restricted to cancer types with at least five evaluable samples. Exact binomial 95% confidence intervals were calculated for prevalence estimates.

Overlap between *ERBB2* amplification and RNA-based screening was quantified in two ways. “Overlap” was defined as tumors positive by both *ERBB2* amplification and the RNA-based method. “Combined yield” was defined as tumors positive by either *ERBB2* amplification or the RNA-based method. Pairwise concordance between methods was summarized using the Jaccard index, calculated as the number of tumors positive by both methods divided by the number positive by either method.

#### 14.3 External validation treated cohort

A treated validation subset was defined as patients who received HER2-targeted therapy after molecular profiling and had available RNA-seq data. The treated validation cohort comprised 113 HER2-targeted treatment episodes from 102 unique patients. Time on treatment and overall survival were evaluable for 110 and 112 episodes, respectively. Response information was available for 104 episodes; two episodes recorded as mixed responses were excluded from the objective-response and clinical-benefit analyses, leaving 102 evaluable episodes for both endpoints.

### 15. HER2-treated clinical utility analyses

The main clinical utility analyses evaluated associations between RNA-based HER2 screening status and clinical outcomes separately in the internal and external validation cohorts. Both ERBB2-amplified and ERBB2-mutated tumors were included in these cohort-specific analyses.

The internal cohort was analyzed at the patient level. In the external validation cohort, patients could contribute more than one eligible HER2-targeted treatment episode; each treatment episode with available molecular and clinical outcome data was analyzed separately. RNA-based HER2 status was assigned using the ERBB2 mRNA threshold and the combined RNA signature definition described above.

Clinical response categories were harmonized across cohorts. Objective response was defined as complete or partial response, and clinical benefit as complete response, partial response, stable disease or non-complete response/non-progressive disease. Associations between RNA-based HER2 status and objective response or clinical benefit were evaluated using Fisher’s exact tests and logistic regression where both outcome classes and both molecular-status groups were represented. Odds ratios are reported for molecular-positive versus molecular-negative tumors. Logistic regression was not performed when complete separation was present.

For time-to-event analyses, progression-free survival was evaluated in the internal cohort. In the external validation cohort, time on treatment was analyzed using treatment discontinuation as the event. Time on treatment was calculated from treatment initiation to treatment discontinuation (event), with episodes lacking a recorded treatment end date censored at the response assessment date. Overall survival was evaluated in both cohorts, with death from any cause as the event and patients without a recorded death censored at last available follow-up. Cox proportional hazards models were used to estimate hazard ratios for molecular-positive versus molecular-negative tumors.

Pooled analyses combined the internal and external treated cohorts and were restricted to observations without an interpreted ERBB2 mutation driver. Tumors with ERBB2 mutation alone or concurrent ERBB2 mutation and amplification were excluded from these pooled analyses to reduce potential confounding by mutation-directed HER2 sensitivity. In these pooled analyses, the cohort-specific primary time-to-event endpoint was progression-free survival for the internal cohort and time on treatment for the external validation cohort.

### 16. Continuous *ERBB2* copy-number versus *ERBB2* mRNA analysis

To compare the outcome associations of RNA-based HER2 assessment and copy number, continuous *ERBB2* copy number and continuous *ERBB2* mRNA expression were compared in the external HER2-treated validation cohort. *ERBB2* minimum copy number and *ERBB2* mRNA expression were analyzed as continuous predictors and standardized to z scores before modelling, such that effect estimates corresponded to a one-standard-deviation increase.

For objective response and clinical benefit, logistic regression models were fitted separately for *ERBB2* copy number and *ERBB2* mRNA. Model performance was summarized using odds ratios with 95% confidence intervals. For time on treatment and overall survival, Cox proportional hazards models were fitted using the respective endpoint-specific follow-up times and censoring definitions described in Section 15. Cox model performance was summarized using hazard ratios with 95% confidence intervals.

### 17. Pooled treated-cohort subgroup analyses

For exploratory subgroup analyses, the internal HER2-treated cohort and the external validation treated cohort were harmonized into a shared patient/treatment-level dataset. Harmonized variables included cohort, patient identifier, tumor type, treatment class, *ERBB2* driver class, *ERBB2* mRNA expression, breast/gastroesophageal RNA classifier score, pan-tumor protein-informed RNA score, RNA-based HER2 screening status, best response.

Subgroup analyses evaluated whether the association between RNA-based HER2 status and the cohort-specific primary time-to-event endpoint was consistent across clinically relevant strata. This endpoint was PFS in the internal cohort and time on treatment in the external validation cohort. These cohort-specific time-to-event endpoints were combined only for exploratory pooled analyses and were not considered equivalent clinical endpoints. Tumor type was grouped as breast cancer, esophageal/gastroesophageal junction cancer and all other tumors. HER2-targeted treatment was grouped as trastuzumab plus pertuzumab, trastuzumab, T-DM1, trastuzumab deruxtecan, HER2 tyrosine kinase inhibitor, other HER2-targeted therapy or unknown. *ERBB2* gene status was grouped as amplified, mutated or none/other, with amplification prioritized when concurrent amplification and mutation were present.

Within each subgroup, Cox proportional hazards models were fitted for the cohort-specific primary time-to-event endpoint, comparing molecular-positive with molecular-negative tumors. Subgroup-specific hazard ratios, 95% confidence intervals, P values and median treatment durations by molecular group were reported where both molecular groups were represented. Interaction tests were performed by comparing Cox models with and without predictor-by-subgroup interaction terms using likelihood-ratio tests.

### 18. Deep Visual Proteomics

**Tile-level deep visual proteomics (TileDVP)** was performed using the previously described TileDVP workflow^14,15^. Tissue sections were digitized, and morphology-defined tissue regions were computationally selected and partitioned into spatially defined image tiles for laser microdissection. The corresponding tissue regions were subjected to high-throughput mass spectrometry (MS)-based proteomic profiling, generating paired spatial and proteomic measurements for each tile. The number of tiles analyzed per specimen varied according to tissue availability and the workflow used in the respective cohort.

Breast and gastroesophageal cancer samples were used as the TileDVP reference cohort, with centrally reviewed HER2 IHC/FISH categories serving as the clinical benchmark. Slides were eligible for analysis if they contained at least 20 tile-level proteomic measurements. After applying this criterion, the breast/gastroesophageal reference cohort comprised 79 samples: 46 breast and 33 gastroesophageal cancer samples (Supplementary Table 18). One gastroesophageal sample had unresolved IHC2+/FISH status and was retained in the descriptive cohort summary but excluded from analyses requiring resolved clinical HER2 classification, leaving 78 evaluable reference samples.

HER2 protein abundance was quantified at tile level and used as the primary spatial proteomic readout. Analytical depth was summarized by the number of quantified peptides and proteins per tile. HER2 detection was assessed at tile level and summarized as the proportion of tiles with detectable HER2 protein per sample and per IHC/FISH reference category. Among HER2-detectable tiles, HER2 abundance was analyzed as a continuous log2-transformed measure, and tile-level detection and abundance distributions were compared across clinical IHC/FISH categories.

TileDVP was additionally performed in a subset of ERBB2-amplified pan-tumor cases treated with HER2-targeted therapy. Tile-level HER2 abundance distributions from these cases were subsequently compared with the breast/gastroesophageal TileDVP reference framework as described below.

### 19. TileDVP-derived HER2 reference curves and classification

Tile-level HER2 protein abundance was analyzed on the log2-transformed scale. Clinical IHC/FISH categories were harmonized before reference-curve construction: IHC0 and ultra-low tumors were grouped as 0/ultra-low, and slides with unresolved IHC2+ FISH status were excluded from classification analyses. Clinical HER2-positive status was defined as IHC2+/FISH-positive or IHC3+; IHC0/ultra-low, IHC1+ and IHC2+/FISH-negative were considered HER2-negative. Analyses were performed at the slide level. Slides were eligible if they contained at least 20 tile-level measurements.

For each slide, a HER2 distribution curve was calculated as the percentage of all sampled tiles with detectable HER2 abundance greater than or equal to each value on a shared log2 abundance grid. Tiles without detectable HER2 remained in the denominator and did not exceed any plotted abundance threshold; a slide without detected HER2 therefore had a zero-valued curve. For classification, a 300-point grid spanning the detected HER2 values in the training reference slides was used. Within each IHC/FISH category, slide-level curves were averaged at every grid value, giving each slide equal weight. The descriptive reference curves shown in the figures were calculated from the full eligible reference cohort. Their pointwise shaded intervals represent the mean ± 1.96 standard errors across slides, bounded between 0% and 100%.

For comparison with the spatial proteomic curves, digital HER2 IHC distribution curves were generated from the fraction of cells in each reference specimen with DAB staining intensity greater than or equal to each evaluated intensity threshold. Within each clinical IHC/FISH group, the plotted IHC curve represents the median specimen-level fraction at each threshold, with bootstrap 95% confidence intervals for that median. Binary HER2-negative and HER2-positive reference curves were also displayed descriptively for digital IHC and DVP.

Each DVP slide curve was compared with the mean curves of the five clinical IHC/FISH reference categories. Distance was defined as the root mean squared error (RMSE) across the abundance grid, and the slide was assigned to the category with the lowest RMSE. The difference between the second-lowest and lowest RMSE was recorded as a classification margin. Binary DVP HER2 status was assigned separately by comparing the slide curve with HER2-negative and HER2-positive mean reference curves; it was not obtained by collapsing the five-category prediction.

Internal classification was assessed using leave-one-slide-out validation. For each held-out reference slide, the abundance grid and category-specific mean curves were reconstructed from the remaining slides before assigning the held-out curve. One slide, rather than every slide from the same patient, was excluded in each fold. Five-category and binary predictions were evaluated separately using their respective reference curves. Accuracy and confusion matrices were calculated against the clinical reference classification.

For the comparison with historical pathology, breast slides with available routine and central GE001 IHC/FISH results were matched to their leave-one-slide-out DVP predictions. The three assessments were compared on the same eligible slide set for five-category IHC/FISH classification and binary HER2 status. Pairwise agreement was calculated as the proportion of slides with matching classifications, with exact binomial 95% confidence intervals. Slides remained the unit of analysis; these intervals did not account for correlation between slides from the same patient or between leave-one-out folds.

To assess relationships with bulk assays, leave-one-slide-out DVP IHC/FISH assignments from the breast and gastroesophageal reference cohort were paired with available bulk ERBB2 mRNA and HER2 protein measurements. Measurements were classified using the previously defined bulk positivity thresholds. Figure 6D includes slides with both bulk measurements; assay-specific pathology–bulk discordance analyses include slides with an available measurement for the respective assay. A pathology–bulk discordance was defined by disagreement between binary clinical HER2 status and the bulk assay classification. DVP-assigned five-category profiles were examined descriptively among these discordant slides.

For exploratory pan-tumor translation, curves from eligible ERBB2-amplified, HER2-treated pan-tumor slides were compared with category-specific mean curves generated from the full breast/gastroesophageal reference cohort. Each slide was assigned to the nearest IHC/FISH-like category by RMSE using the full-reference abundance grid. These labels indicate similarity to spatial proteomic reference profiles and do not constitute clinical IHC or FISH results. DVP profiles were compared descriptively with response, PFS and overall survival.

### 20. Statistical analysis

Continuous variables were summarized using medians and interquartile ranges unless otherwise specified. Categorical variables were summarized using counts and percentages. Comparisons between two groups were performed using Wilcoxon rank-sum tests for continuous variables and Fisher’s exact tests or χ² tests for categorical variables, as appropriate. Ordinal trends across HER2 IHC/FISH categories were assessed using trend tests appropriate for ordered groups. Correlations between *ERBB2* mRNA and HER2 protein abundance were assessed using Pearson and Spearman correlation coefficients.

Associations between HER2 screening status and objective response or clinical benefit were evaluated using Fisher’s exact tests and logistic regression where model assumptions were met. Logistic regression was not performed in analyses with complete separation. Progression-free survival and OS were analyzed using Kaplan–Meier methods, log-rank tests and Cox proportional hazards models. Hazard ratios are reported with 95% confidence intervals. Continuous predictor analyses used standardized predictors scaled per one standard deviation increase.

All statistical tests were two-sided unless otherwise specified. P values below 0.05 were considered statistically significant. Where multiple testing was performed in exploratory analyses, false discovery rate adjustment was applied using the Benjamini–Hochberg procedure.

### 21. Ethics, approvals and trial registration

The Copenhagen Prospective Personalized Oncology (CoPPO) study was conducted in accordance with the Declaration of Helsinki and registered at ClinicalTrials.gov (NCT02290522). The study was approved by the Danish Ethical Committee/Regional Ethics Committee (file no. 1300530). All patients provided written informed consent before study inclusion and molecular profiling.

Patients treated in the ProTarget trial were enrolled under a Danish nationwide, non-randomized, multi-drug, open-label phase II basket protocol registered at ClinicalTrials.gov (NCT04341181). The protocol was approved by the Danish Ethics Committee (H-19089780) and the Danish Medicines Agency. All patients provided written informed consent before treatment initiation.

External validation analyses using Hartwig Medical Foundation data were performed under Hartwig Medical Database Data Access Request DR-505 and the associated data access and license agreements. Analyses were conducted on derived molecular and clinical variables, and no original licensed patient-level data are redistributed with this publication.

## Data availability

Raw and patient-level genomic, transcriptomic, bulk proteomic and clinical data from the CoPPO and ProTarget cohorts are not publicly redistributed because they contain sensitive patient-level molecular and clinical information and are governed by institutional approvals, informed consent and applicable data-protection requirements. Access requests may be considered through the relevant institutional data-access procedures, subject to the necessary approvals.

The list of RefSeq-aligned transcripts, shareable derived deep visual proteomics (DVP) data and accompanying DVP metadata will be publicly available at time of publication and during review through Figshare (DOI: 10.6084/m9.figshare.34300074). The deposition will describe the included datasets; raw mass-spectrometry files are not included.

External validation data from Hartwig Medical Foundation were accessed under Data Access Request DR-505 and the associated data-access and license agreements. These licensed data cannot be redistributed by the authors. Researchers may apply for access directly through Hartwig Medical Foundation according to its data-access procedures.

Summary results are reported in the manuscript and Supplementary Information. Apart from the publicly deposited DVP data, individual-level source data underlying the figures are subject to the access restrictions described above.

## Code availability

The RNA-seq–based HER2 signatures, including the final signature gene lists, trained model objects and R scripts for model scoring, will be publicly available through Figshare at publication and during review (10.6084/m9.figshare.34300074), enabling independent application of the signatures. Scripts for recreating Fig6B and 6E are also available.

## Acknowledgements

We thank the investigators of the ProTarget Study Group for their contributions to patient recruitment, clinical data collection, biospecimen acquisition and study infrastructure. This publication and the underlying study were made possible in part by data made available through the Hartwig Medical Database by Hartwig Medical Foundation and the Center of Personalized Cancer Treatment (CPCT), both based in the Netherlands.

## Author contributions

K.E., K.S.R., F.O.B., U.N.L. and M.M.-S. conceived and designed the study. K.E. performed the primary statistical analyses, integrated the clinical, genomic, transcriptomic and proteomic datasets, generated the figures and wrote the original manuscript draft.

K.E., F.O.B., M.F.L and L.R. performed RNA data processing and bioinformatic analyses, including development, evaluation and interpretation of the RNA-based HER2 models. K.E. and B.H.C. reassessed ERBB2 amplification status and contributed to interpretation of the genomic analyses. A.B.N. and J.W. generated, processed and analyzed the bulk proteomic data and contributed to interpretation of the bulk proteomic findings. X.Z. performed digital HER2 immunohistochemistry analysis, including cell-wise quantification of HER2 staining intensity.

A.-V.L., M.-L.M.T., R.S.G. and M.K.N. performed pathological assessment of HER2 immunohistochemistry and contributed to interpretation of the pathology data. E.C., M.T.S. and A.M. performed deep visual proteomics experiments and contributed to spatial proteomic data generation, analysis and interpretation. K.S.R. and M.H. contributed to patient recruitment and inclusion in the CoPPO and ProTarget studies. T.K. contributed to ProTarget trial execution and data management. A.Y.S., D.A., U.N.L. and M.M.-S. contributed to clinical study implementation, cohort annotation and interpretation of the clinical data. The ProTarget Study Group contributed to patient recruitment, clinical data collection, biospecimen acquisition and study infrastructure.

All authors contributed to interpretation of the findings, reviewed and edited the manuscript, approved the final version and accept responsibility for the integrity of the work.

## Use of artificial intelligence and figure creation tools

During preparation of this manuscript, the authors used ChatGPT (OpenAI) to assist with language editing and improvement of manuscript clarity, as well as with the development, refinement and troubleshooting of analysis code. ChatGPT was also used to assist in the graphical design and generation of Figure 6A. All AI-assisted outputs, including generated or modified code, were reviewed, verified and, where necessary, revised by the authors. The authors take full responsibility for the content of the manuscript and the accuracy of the analyses.

BioRender was used for the creation of graphical elements in Figures 1 and 6A. Figure 1 was created in BioRender: Egebjerg, K. (2026) https://BioRender.com/5knahs6. Figure 6A was created in BioRender: Egebjerg, K. (2026), https://BioRender.com/88798vd.

## References

1. Bang, Y. J. et al. Trastuzumab in combination with chemotherapy versus chemotherapy alone for treatment of HER2-positive advanced gastric or gastro-oesophageal junction cancer (ToGA): A phase 3, open-label, randomised controlled trial. The Lancet 376, 687–697 (2010).

2. Slamon, D. J. et al. Use of Chemotherapy plus a Monoclonal Antibody against HER2 for Metastatic Breast Cancer That Overexpresses HER2. New England Journal of Medicine 344, 783–792 (2001).

3. Wolff, A. C. et al. Human Epidermal Growth Factor Receptor 2 Testing in Breast Cancer: ASCO-College of American Pathologists Guideline Update. Journal of Clinical Oncology 41, 3867–3872 (2023).

4. Bartley, A. N. et al. HER2 Testing and Clinical Decision Making in Gastroesophageal Adenocarcinoma: Guideline From the College of American Pathologists, American Society for Clinical Pathology, and the American Society of Clinical Oncology. J. Clin. Oncol. 35, 446–464 (2017).

5. Meric-Bernstam, F. et al. Efficacy and Safety of Trastuzumab Deruxtecan in Patients With HER2-Expressing Solid Tumors: Primary Results From the DESTINY-PanTumor02 Phase II Trial. Journal of Clinical Oncology 42, 47–58 (2023).

6. FDA grants accelerated approval to fam-trastuzumab deruxtecan-nxki for unresectable or metastatic HER2-positive solid tumors | FDA. https://www.fda.gov/drugs/resources-information-approved-drugs/fda-grants-accelerated-approval-fam-trastuzumab-deruxtecan-nxki-unresectable-or-metastatic-her2.

7. Jorgensen, J. T., Egebjerg, K., Lænkholm, A. V. & Mau-Sørensen, M. Human Epidermal Growth Factor Receptor 2 Pan Tumor Indication for Trastuzumab Deruxtecan. Journal of Clinical Oncology 10.1200/JCO.24.00884 (2024) doi:10.1200/JCO.24.00884.

8. Shayeb, A. M., Kurzrock, R., Adashek, J. J. & Kato, S. Comprehensive Analysis of Human Epidermal Growth Factor Receptor 2 Through DNA, mRNA, and Protein in Diverse Malignancies. JCO Precis. Oncol. 10.1200/PO.22.00604/SUPPL_FILE/DS.PO.22.00604.PDF (2023) doi:10.1200/PO.22.00604/SUPPL_FILE/DS.PO.22.00604.PDF.

9. Makker, V. et al. 148P DESTINY-PanTumor02 study of trastuzumab deruxtecan (T-DXd) in patients (pts) with HER2-expressing solid tumors: Exploratory biomarker analyses of HER2 expression and gene amplification in tissue and plasma. Annals of Oncology 34, S239–S240 (2023).

10. Yang, W. et al. 177P HER2 testing in multiple solid tumors: Concordance between 3 scoring algorithms. Annals of Oncology 35, S286–S287 (2024).

11. Bryant, D. et al. A Real-World Experience in Pan-Tumor Testing for HER2 IHC in More Than 65000 Solid Tumors. JAMA Oncol. 11, 919–921 (2025).

12. Ross, J. et al. A phase 2 basket trial of ado-trastuzumab emtansine for patients with HER2 amplified cancers. Journal of Clinical Oncology 43, 3020–3020 (2025).

13. Li, Z. et al. A pan-cancer analysis of HER2 index revealed transcriptional pattern for precise selection of HER2-targeted therapy. EBioMedicine 62, (2020).

14. Mund, A. et al. Deep Visual Proteomics defines single-cell identity and heterogeneity. Nature Biotechnology 2022 40:8 40, 1231–1240 (2022).

15. Mathian, É. et al. TileDVP: Decoding the Tissue Proteome from H&E Images. Lecture Notes in Computer Science **16126 LNCS**, 21–31 (2026).

16. Priestley, P. et al. Pan-cancer whole-genome analyses of metastatic solid tumours. Nature 575, 210–216 (2019).

17. Ogitani, Y., Hagihara, K., Oitate, M., Naito, H. & Agatsuma, T. Bystander killing effect of DS-8201a, a novel anti-human epidermal growth factor receptor 2 antibody–drug conjugate, in tumors with human epidermal growth factor receptor 2 heterogeneity. Cancer Sci. 107, 1039–1046 (2016).

18. Modi, S. et al. Trastuzumab Deruxtecan in Previously Treated HER2-Low Advanced Breast Cancer. New England Journal of Medicine 387, 9–20 (2022).

19. Tuxen, I. V. et al. Copenhagen Prospective Personalized Oncology (CoPPO)-Clinical Utility of Using Molecular Profiling to Select Patients to Phase I Trials. Clin. Cancer Res. 25, 1239–1247 (2019).

20. Belcaid, L. et al. Copenhagen Prospective Personalized Oncology (CoPPO)—impact of comprehensive genomic profiling in more than 2000 patients in a phase I setting. Annals of Oncology 36, 1078–1087 (2025).

21. Kringelbach, T. et al. ProTarget: a Danish Nationwide Clinical Trial on Targeted Cancer Treatment based on genomic profiling – a national, phase 2, prospective, multi-drug, non-randomized, open-label basket trial. BMC Cancer 23, 1–12 (2023).

22. Chambers, M. C., et al. A cross-platform toolkit for mass spectrometry and proteomics. Nature Biotechnology vol. 30 918–920 Preprint at 10.1038/nbt.2377 (2012).

