## Supplementary Information for "Beyond ERBB2 amplification: RNA and proteomic HER2 assessment for pan-tumor treatment stratification"

Supplementary Table 1. Baseline characteristics of the clinical reference and internal pan-tumor screening cohorts. 4

Supplementary Figure 1. HER2 IHC and FISH distributions in the clinical reference cohort. 5

Supplementary Table 2. FISH amplification status among HER2 IHC3+ tumors. 5

Supplementary Figure 2. ERBB2 and chromosome 17 centromere copy counts by FISH. 6

Supplementary Figure 3. Concordance of ERBB2 CNV calls with FISH and clinical IHC/FISH classification 7

Supplementary Table 3. Diagnostic performance of ERBB2 CNV against clinical IHC/FISH status. 7

Supplementary Figure 4. HER2 IHC distributions across ASCO/CAP-defined FISH groups in breast and gastroesophageal cancers. 8

Supplementary Figure 5. Molecular profiles of FISH/IHC-discordant tumors. 9

Supplementary Table 4. Characteristics of selected FISH/IHC-discordant tumors. 9

Supplementary Figure 6. Correlations among genomic, RNA, protein and image-derived HER2 measurements. 10

Supplementary Figure 7. ERBB2 mRNA and HER2 protein abundance across clinical HER2 categories. 11

Supplementary Table 5. Heterogeneous and cytoplasmic HER2 staining by IHC category. 12

Supplementary Figure 8. Digital quantification of pathologist-assessed cytoplasmic HER2 staining. 12

Supplementary Table 6. Diagnostic performance of molecular HER2 assessment methods. 13

Supplementary Figure 9. Leading transcripts in the BG RNA signature. 14

**Supplementary Table 1. Baseline characteristics of the clinical reference and internal pan-tumor screening cohorts.**

| Category | | Breast Cancer Patients N=181 | GE Cancer Patients (N=48) | Pan Tumor Cohort (N=1331) |
|---|---|---|---|---|
| **Patient Age** | Median [Q1-Q3] | 54 [47-63] | 62 [53-70] | 62 [53-69] |
| **Gender** | Male | 1% | 71% | 49% |
| | Female | 99% | 29% | 51% |
| **Number of Prior Chemotherapeutic treatments** | Median [Q1-Q3] | 4 [2-6] | 2 [2-3] | 2 [2-3] |
| **Biopsy Location** | Metastasis/Recurrence | 97% | 98% | NA |
| | Primary | 3% | 2% | NA |
| **Biopsy Site** | Liver | 53% | 51% | 40% |
| | Lymph Node | 17% | 6% | 10% |
| | Lung/Pleura | 2% | 4% | 11% |
| | Cutaneous / Subcutaneous | 9% | 4% | 4% |
| | Breast | 14% | 0 | 1% |
| | Peritoneal Metastasis | 1% | 2% | 4% |
| | Other | 4% | 33% | 14% |
| | N/A | 0 | 0 | 16% |
| **ER Status:** | ER-Positive | 49% | | |
| | ER-Negative | 51% | | |
| **GE Cancer primary tumor location** | Esophageal | | 12.5% | |
| | Gastric-esophageal junction | | 75% | |
| | Gastric | | 12.5% | |

Clinical characteristics and molecular-data availability in the breast, gastroesophageal and internal pan-tumor cohorts. Continuous variables are summarized as median (interquartile range); categorical variables are reported as indicated.

### Supplementary Figure 1. HER2 IHC and FISH distributions in the clinical reference cohort.

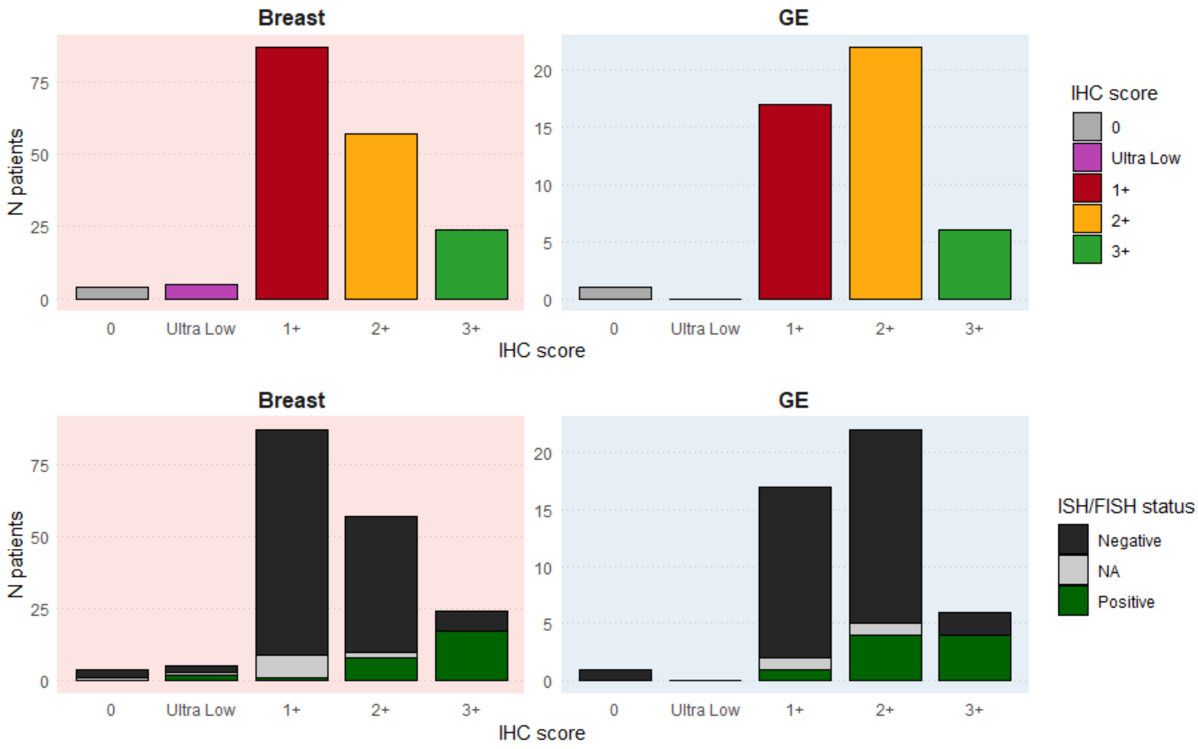

Upper panels show sample counts by IHC category in breast and gastroesophageal cancers. Lower panels show the corresponding distributions of positive, negative and unavailable FISH results.

### Supplementary Table 2. FISH amplification status among HER2 IHC3+ tumors.

| | IHC3+ Total | IHC3+ FISH pos | IHC3+ FISH neg | % not FISH+ |
|---|---|---|---|---|
| Breast | 24 | 17 | 7 | 29% |
| GE | 6 | 4 | 2 | 33% |

Numbers of IHC3+ breast and gastroesophageal tumors with positive or negative FISH results and the proportion without FISH-detected amplification.

**Supplementary Figure 2. ERBB2 and chromosome 17 centromere copy counts by FISH.**

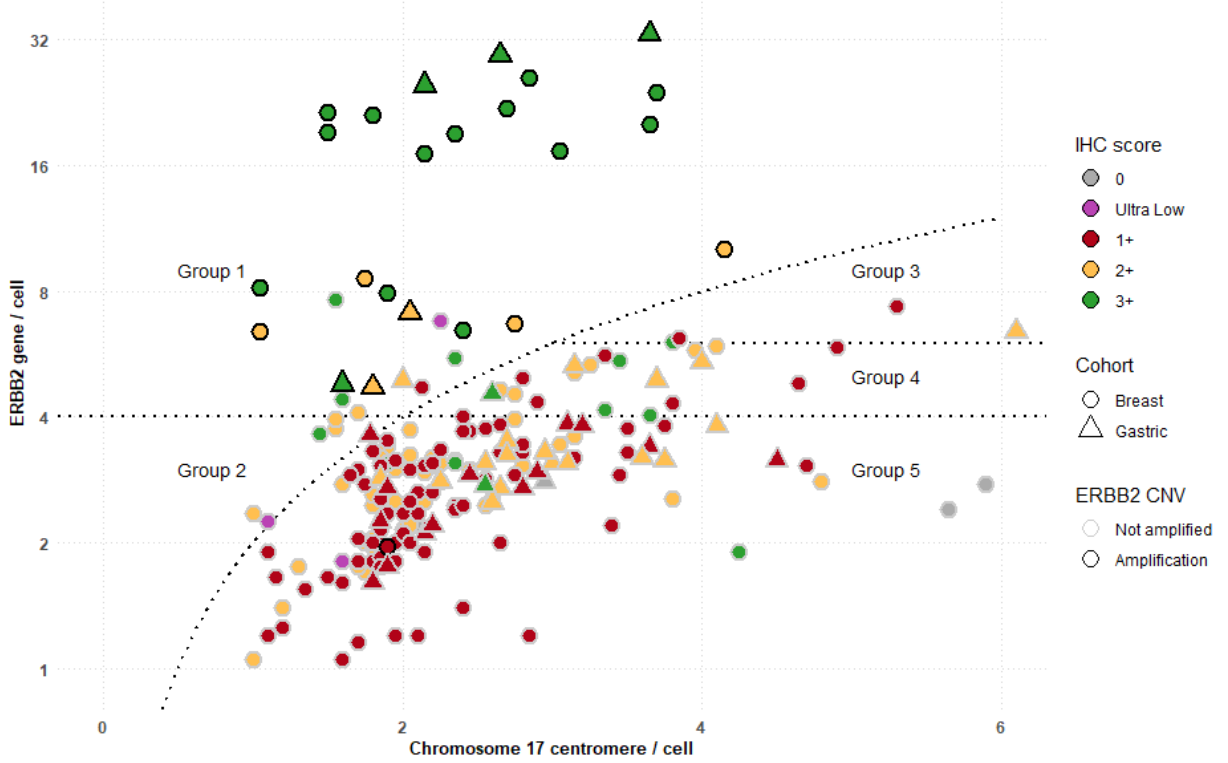

Each point represents a sample. Colors indicate IHC category, shapes distinguish breast and gastroesophageal tumors, and point opacity indicates genomic ERBB2 amplification status. Dotted lines delineate the displayed FISH groups.

### Supplementary Figure 3. Concordance of ERBB2 CNV calls with FISH and clinical IHC/FISH classification

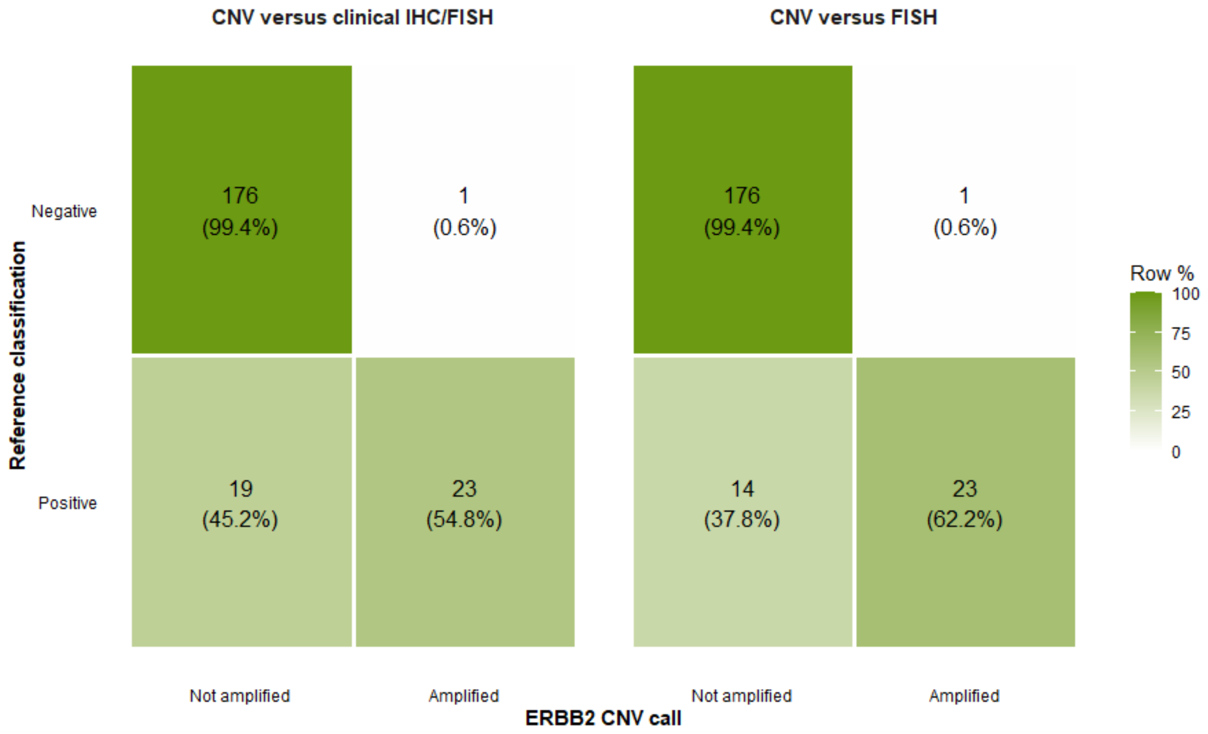

Confusion matrices compare genomic ERBB2 amplification calls with clinical IHC/FISH status and FISH status separately. Cells show sample counts and percentages within each reference-classification row.

### Supplementary Table 3. Diagnostic performance of ERBB2 CNV against clinical IHC/FISH status.

| Predictor | N | TP | FP | FN | TN | AUC | Sensitivity | Specificity | PPV | NPV | Accuracy |
|---|---|---|---|---|---|---|---|---|---|---|---|
| ERBB2 CNV | 219 | 23 | 1 | 19 | 176 | 0.771 | 0.548 | 0.994 | 0.958 | 0.903 | 0.909 |

Performance was evaluated in 219 samples with available CNV calls and resolved clinical HER2 status. IHC2+ samples without available FISH results were excluded. AUC was calculated from the binary amplification call.

**Supplementary Figure 4. HER2 IHC distributions across ASCO/CAP-defined FISH groups in breast and gastroesophageal cancers.**

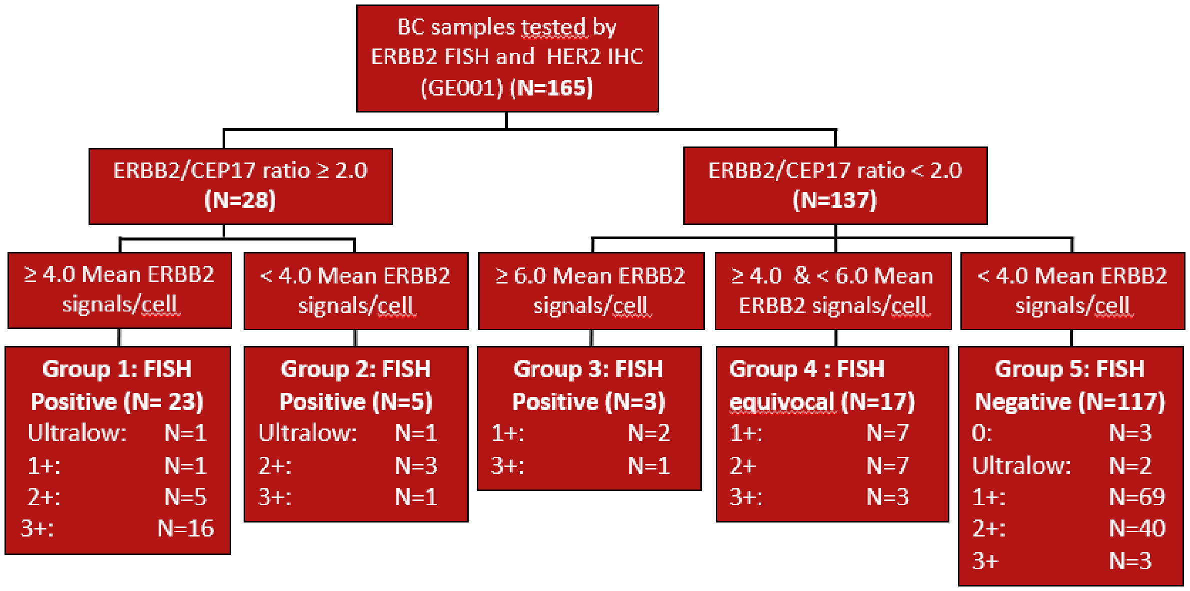

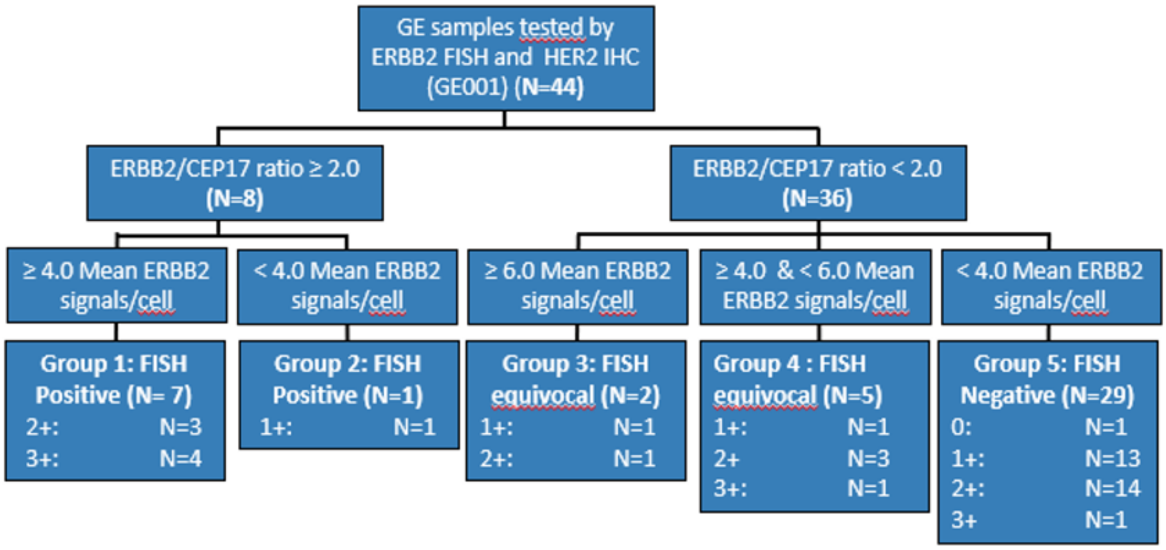

Flow diagrams classify breast (upper panel) and gastroesophageal (lower panel) tumors according to the five FISH groups defined by the ASCO/CAP breast cancer guidelines, based on the ERBB2/CEP17 ratio and mean ERBB2 signals per cell. The same group definitions were applied descriptively to gastroesophageal tumors to facilitate cross-tumor comparison. Each group reports the number of samples and corresponding HER2 IHC-score distribution.

### Supplementary Figure 5. Molecular profiles of FISH/IHC-discordant tumors.

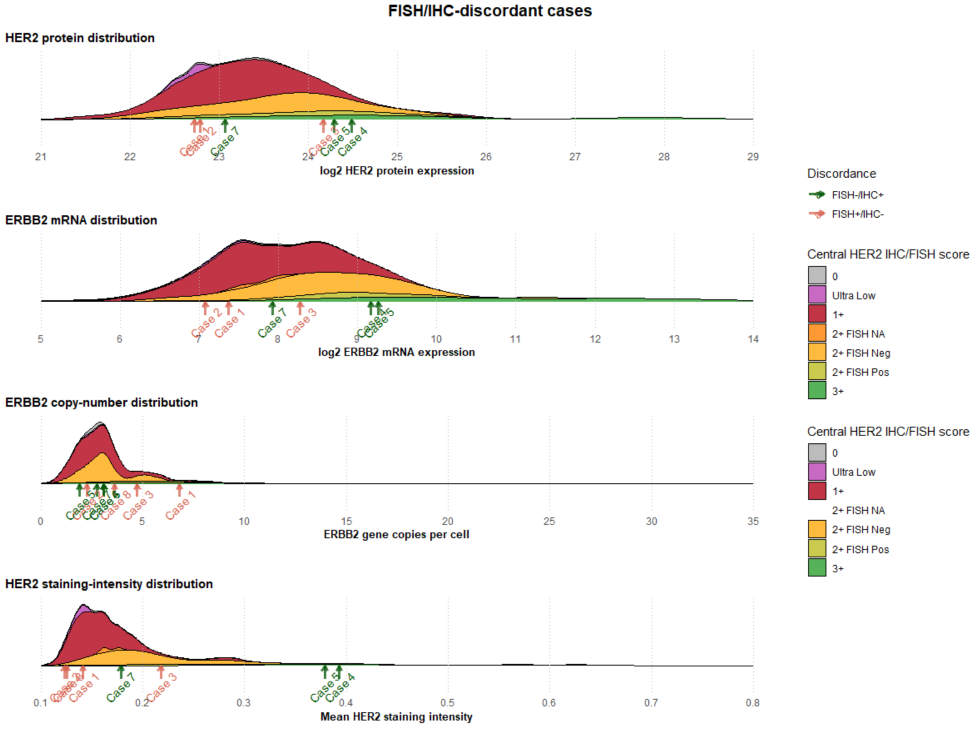

Distributions of HER2 protein, ERBB2 mRNA, ERBB2 gene copy counts and quantitative HER2 staining intensity are shown by clinical IHC/FISH category. Annotated arrows identify discordant cases and the direction of discordance.

### Supplementary Table 4. Characteristics of selected FISH/IHC-discordant tumors.

| | Cancer Type | Biopsy Location | FISH Group | IHC | Heterogeneous Staining | Discrepancy |
|---|---|---|---|---|---|---|
| *Case 1* | Breast | Lymph Node | Group 1 | Ultra Low | yes | FISH+ & IHC- |
| *Case 2* | Breast | Lymph Node | Group 1 | 1+ | yes | FISH+ & IHC- |
| *Case 3* | Breast | Skin | Group 2 | Ultra Low | yes | FISH+ & IHC- |
| *Case 4* | GE | Adrenal Gland | Group 2 | 1+ | no | FISH+ & IHC- |
| *Case 5* | Breast | Liver | Group 5 | 3+ | yes | IHC+ & FISH- |
| *Case 6* | Breast | Liver | Group 5 | 3+ | yes | IHC+ & FISH- |
| *Case 7* | Breast | Lymph Node | Group 5 | 3+ | no | IHC+ & FISH- |
| *Case 8* | GE | Liver | Group 5 | 3+ | yes | IHC+ & FISH- |

Case-level tumor type, biopsy site, FISH group, IHC score and staining heterogeneity for the discordant samples highlighted in Supplementary Figure 5.

**Supplementary Figure 6. Correlations among genomic, RNA, protein and image-derived HER2 measurements.**

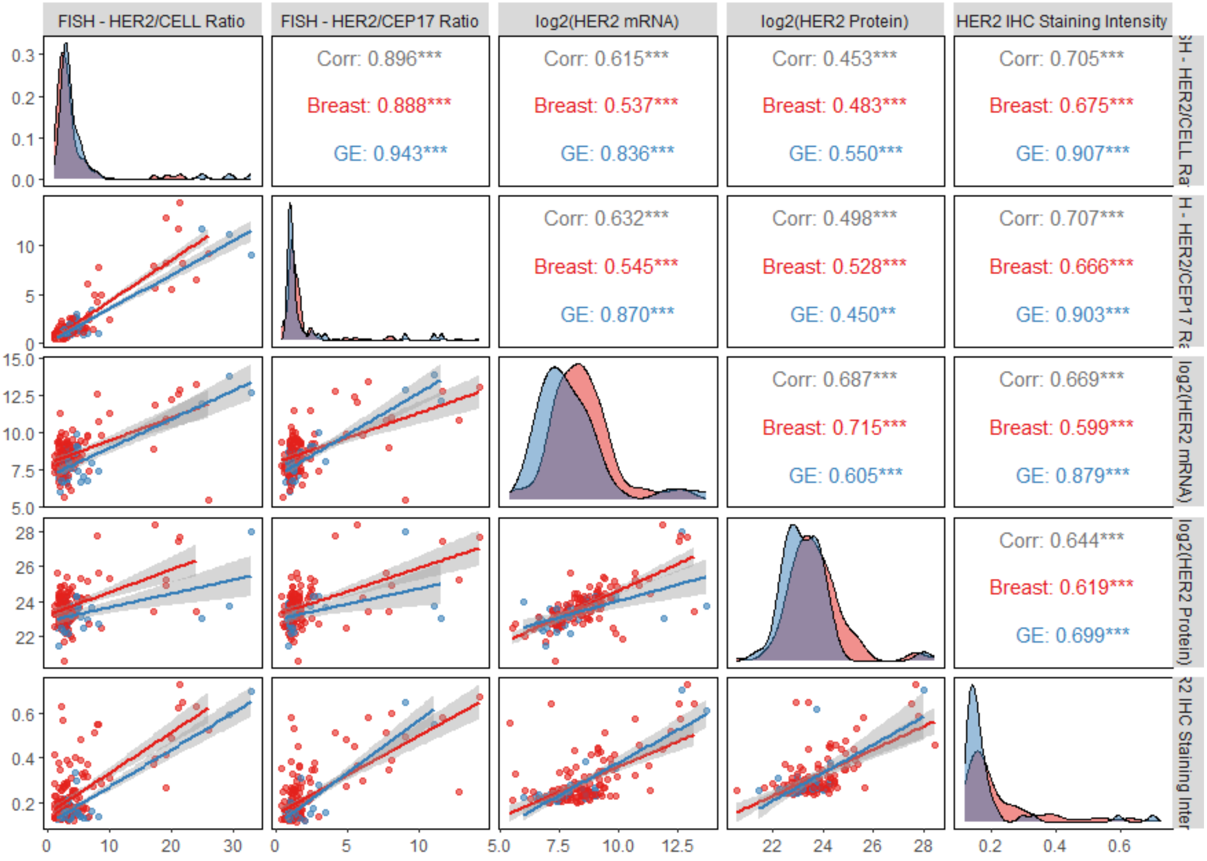

Pairwise comparisons of FISH measurements, ERBB2 mRNA, HER2 protein and membranous staining intensity. Diagonal panels show distributions; lower panels show scatterplots with regression fits and 95% confidence bands; upper panels report Pearson correlations overall and by tumor type.

**Supplementary Figure 7. ERBB2 mRNA and HER2 protein abundance across clinical HER2 categories.**

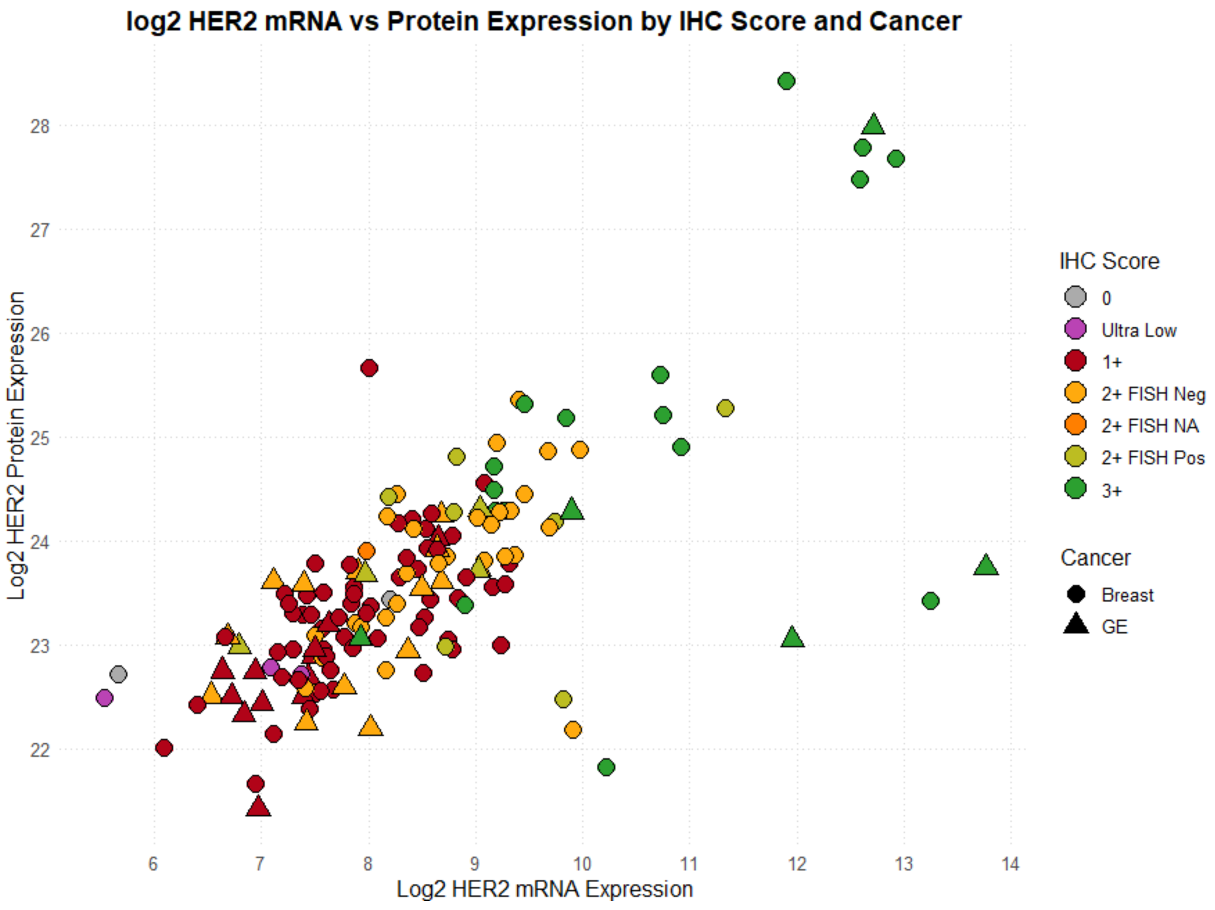

Each point represents a sample with available RNA, protein and IHC/FISH annotation. Colors indicate clinical HER2 category and shapes distinguish breast from gastroesophageal cancers.

**Supplementary Table 5. Heterogeneous and cytoplasmic HER2 staining by IHC category.**

| IHC Score | Heterogeneous Staining | |
|---|---|---|
| | No (N) | Yes (N) |
| 0 | 2 | 2 |
| Ultra Low | 1 | 4 |
| 1+ | 51 | 53 |
| 2+ | 43 | 36 |
| 3+ | 22 | 8 |

| IHC Score | Cytoplasmic Staining | |
|---|---|---|
| | No (N) | Yes (N) |
| 0 | 3 | 1 |
| Ultra Low | 3 | 2 |
| 1+ | 47 | 57 |
| 2+ | 25 | 54 |
| 3+ | 5 | 25 |

Numbers of samples with and without pathologist-assessed heterogeneous or cytoplasmic staining within each IHC category.

**Supplementary Figure 8. Digital quantification of pathologist-assessed cytoplasmic HER2 staining.**

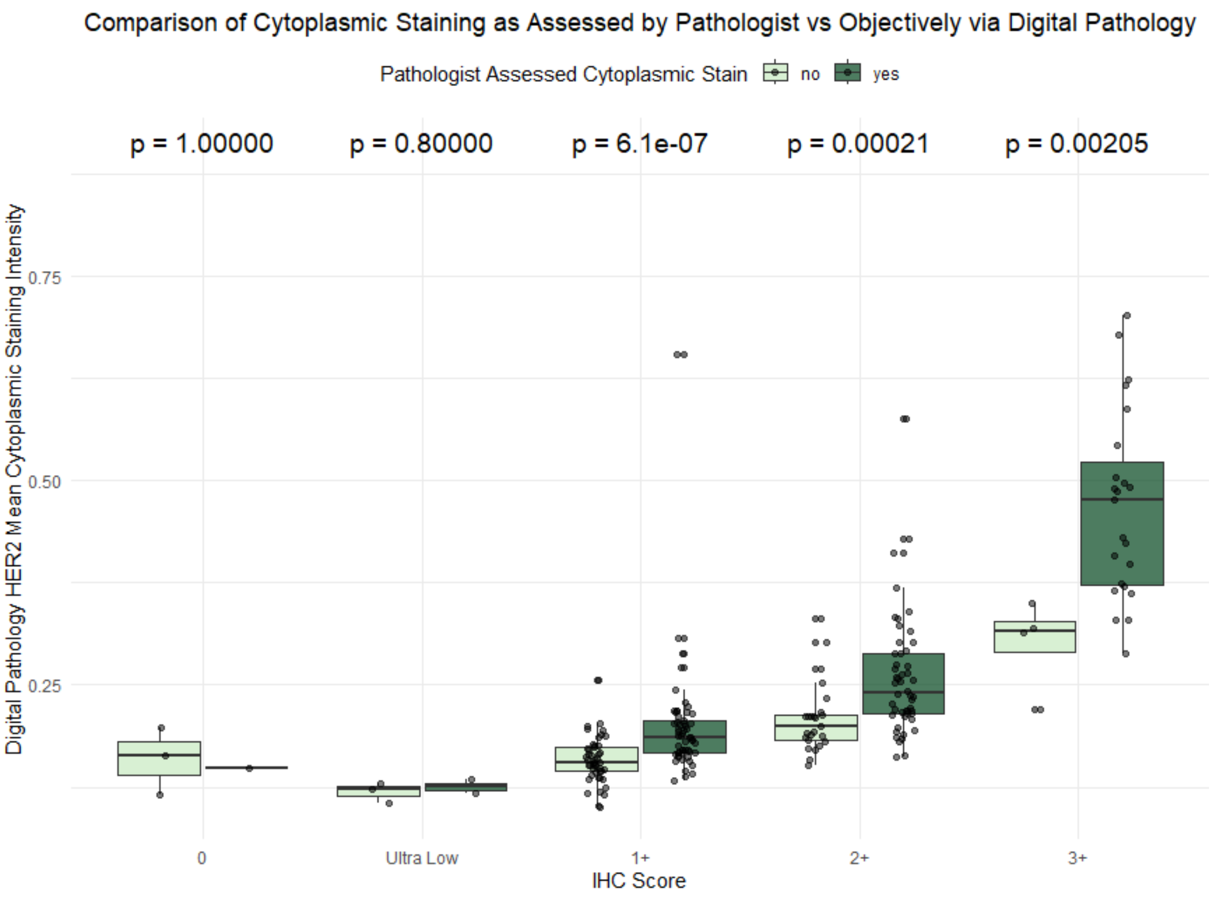

Sample-level mean cytoplasmic HER2 staining intensity measured using QuPath, grouped by IHC category and the presence or absence of pathologist-assessed cytoplasmic staining.

#### Supplementary Table 6. Diagnostic performance of molecular HER2 assessment methods.

| Model | Predictor | N | TP | FP | FN | TN | AUC | Sensitivity | Specificity | PPV | NPV | Accuracy |
|---|---|---|---|---|---|---|---|---|---|---|---|---|
| Fixed binary call | ERBB2 CNV | 219 | 23 | 1 | 19 | 176 | 0.771 | 0.548 | 0.994 | 0.958 | 0.903 | 0.909 |
| Logistic regression | ERBB2 mRNA | 152 | 17 | 4 | 15 | 116 | 0.867 | 0.531 | 0.967 | 0.810 | 0.885 | 0.875 |
| Logistic regression | HER2 protein | 152 | 10 | 2 | 22 | 118 | 0.774 | 0.312 | 0.983 | 0.833 | 0.843 | 0.842 |
| Logistic regression | mRNA + protein | 152 | 16 | 5 | 16 | 115 | 0.869 | 0.500 | 0.958 | 0.762 | 0.878 | 0.862 |
| Random forest | BG RNA signature | 171 | 17 | 1 | 19 | 134 | 0.862 | 0.472 | 0.993 | 0.944 | 0.876 | 0.883 |

Learned models were evaluated using pooled out-of-fold probabilities at a positivity threshold of 0.50. ERBB2 CNV was evaluated as a fixed binary comparator. Method-specific denominators and performance against clinical IHC/FISH status are reported.

### Supplementary Figure 9. Leading transcripts in the BG RNA signature.

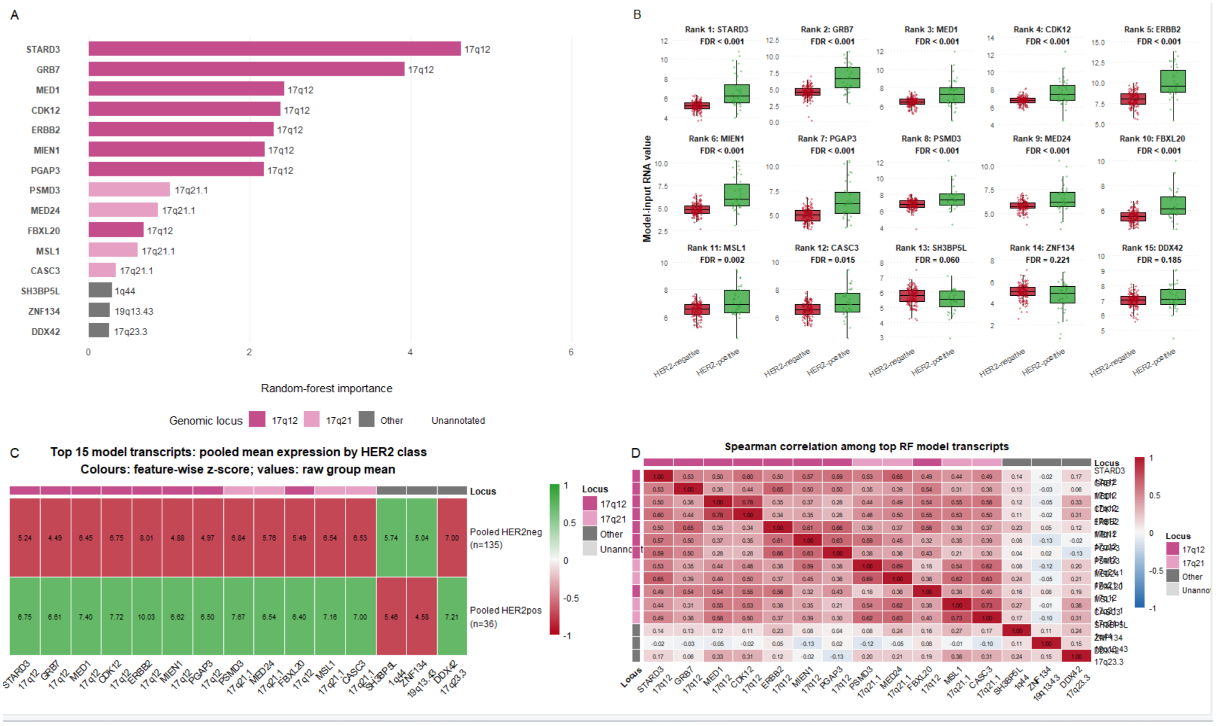

Random-forest importance of the 15 leading transcripts. b, Transcript expression by clinical HER2 status. c, Class-level mean expression, with colors representing feature-wise z-scores. d, Pairwise Spearman correlations among the selected transcripts. Genomic loci are annotated.

**Supplementary Table 7. Tumor-type distribution in the internal pan-tumor screening cohort.**

| ***Cancer type*** | **N** |
|---|---|
| *Colorectal cancer* | 327 |
| *NSCLC* | 165 |
| *Breast cancer* | 115 |
| *Prostate cancer* | 69 |
| *Ovarian cancer* | 63 |
| *Pancreatic cancer* | 57 |
| *BTC* | 41 |
| *Head and neck cancer* | 36 |
| *Mesothelioma* | 36 |
| *CUP* | 35 |
| *Cervical cancer* | 31 |
| *NEC* | 31 |
| *SCLC* | 31 |
| *Gastric/GEJ cancer* | 26 |
| *Urothelial cancer* | 26 |
| *Melanoma* | 25 |
| *Sarcoma* | 23 |
| *Esophageal SCC* | 17 |
| *Adrenal cancer* | 16 |
| *Endometrial cancer* | 16 |
| *N/A* | 12 |
| *Liver cancer* | 10 |
| *Salivary gland cancer* | 10 |
| *Thymic carcinoma* | 10 |
| *Brain cancer* | 9 |
| *Esophageal cancer* | 8 |
| *MiNEN* | 8 |
| *Small intestine cancer* | 8 |
| *Penile cancer* | 7 |
| *Testicular cancer* | 7 |
| *Thymoma* | 7 |
| *Thyroid cancer* | 7 |
| *Appendiceal cancer* | ≤ 5 |
| *Lymphoma non-Hodgkin* | ≤ 5 |
| *NET* | ≤ 5 |
| *Germ cell tumor* | ≤ 5 |
| *Renal cell carcinoma* | ≤ 5 |
| *AML* | ≤ 5 |
| *Skin cancer - non-melanoma* | ≤ 5 |
| *Vaginal cancer* | ≤ 5 |
| *Anal cancer* | ≤ 5 |
| *NUT carcinoma* | ≤ 5 |
| *Peritoneal cancer* | ≤ 5 |
| *Fallopian tube cancer* | ≤ 5 |
| *MANEC* | ≤ 5 |
| *Other* | ≤ 5 |
| *Pseudomyxoma peritonei* | ≤ 5 |

Numbers of samples assigned to each tumor type in the internal screening cohort.

### Supplementary Figure 10. Ranked molecular HER2 probabilities in the internal screening cohort.

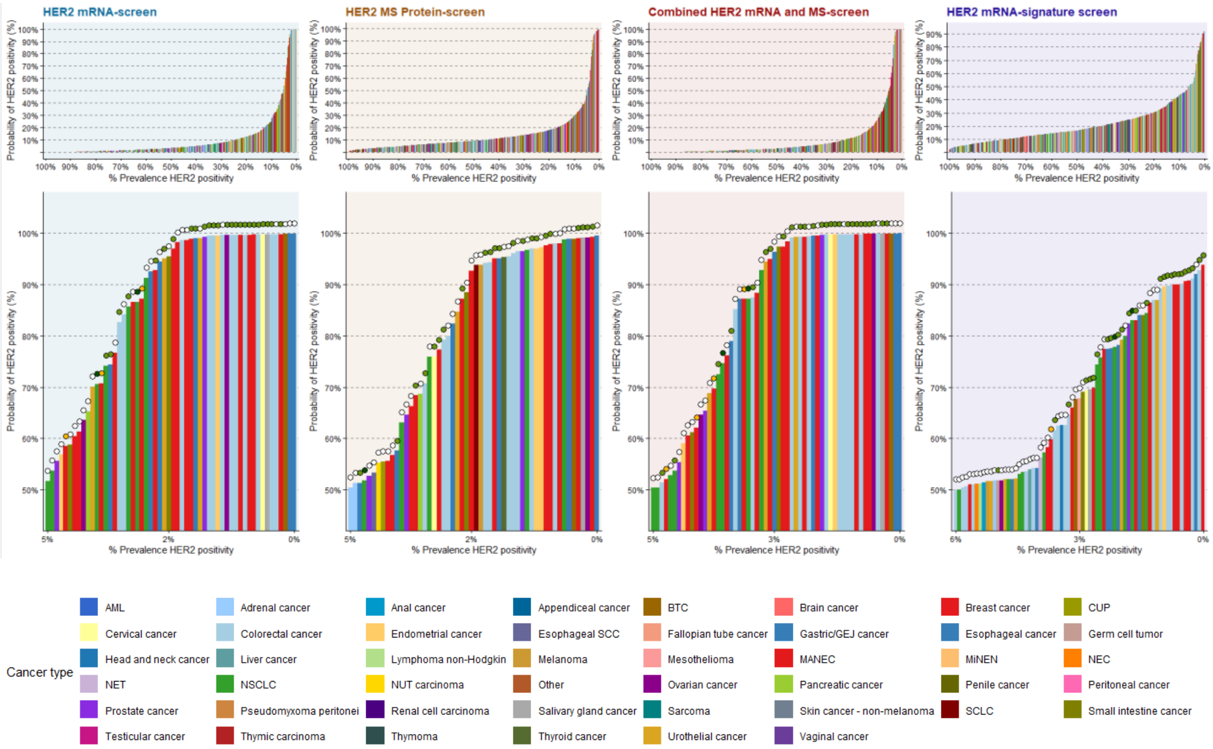

Samples are ranked by predicted HER2 probability for ERBB2 mRNA, HER2 protein, the mRNA + protein model and the BG RNA signature. Bar colors indicate tumor type; overlaid symbols indicate ERBB2 genomic status. Lower panels enlarge the high-probability region.

### Supplementary Table 8. Molecular HER2 positivity by tumor type and screening method.

| Cancer Type | Method | N | N positive | Prevalence (%) | 95% CI (%) |
|---|---|---|---|---|---|
| **Overall** | ERBB2 mRNA | 1130 | 56 | 5.0 | 3.8–6.4 |
| **Overall** | HER2 protein | 1130 | 54 | 4.8 | 3.6–6.2 |
| **BTC** | ERBB2 mRNA | 32 | 1 | 3.1 | 0.1–16.2 |
| **BTC** | HER2 protein | 32 | 1 | 3.1 | 0.1–16.2 |
| **Colorectal cancer** | ERBB2 mRNA | 274 | 12 | 4.4 | 2.3–7.5 |
| **Colorectal cancer** | HER2 protein | 274 | 10 | 3.6 | 1.8–6.6 |
| **Breast cancer** | ERBB2 mRNA | 107 | 16 | 15.0 | 8.8–23.1 |
| **Breast cancer** | HER2 protein | 107 | 13 | 12.1 | 6.6–19.9 |
| **Gastric/GEJ/Esophageal cancer** | ERBB2 mRNA | 28 | 6 | 21.4 | 8.3–41.0 |
| **Gastric/GEJ/Esophageal cancer** | HER2 protein | 28 | 6 | 21.4 | 8.3–41.0 |
| **NSCLC** | ERBB2 mRNA | 142 | 7 | 4.9 | 2.0–9.9 |
| **NSCLC** | HER2 protein | 142 | 5 | 3.5 | 1.2–8.0 |
| **SCLC** | ERBB2 mRNA | 22 | 0 | 0.0 | 0.0–15.4 |
| **SCLC** | HER2 protein | 22 | 1 | 4.5 | 0.1–22.8 |
| **Pancreatic cancer** | ERBB2 mRNA | 50 | 1 | 2.0 | 0.1–10.6 |
| **Pancreatic cancer** | HER2 protein | 50 | 1 | 2.0 | 0.1–10.6 |
| **Ovarian cancer** | ERBB2 mRNA | 62 | 2 | 3.2 | 0.4–11.2 |
| **Ovarian cancer** | HER2 protein | 62 | 1 | 1.6 | 0.0–8.7 |
| **Urothelial cancer** | ERBB2 mRNA | 25 | 3 | 12.0 | 2.5–31.2 |
| **Urothelial cancer** | HER2 protein | 25 | 1 | 4.0 | 0.1–20.4 |
| **Prostate cancer** | ERBB2 mRNA | 66 | 2 | 3.0 | 0.4–10.5 |
| **Prostate cancer** | HER2 protein | 66 | 3 | 4.5 | 0.9–12.7 |
| **Cervical cancer** | ERBB2 mRNA | 31 | 1 | 3.2 | 0.1–16.7 |
| **Cervical cancer** | HER2 protein | 31 | 1 | 3.2 | 0.1–16.7 |
| **Head and neck cancer** | ERBB2 mRNA | 34 | 0 | 0.0 | 0.0–10.3 |
| **Head and neck cancer** | HER2 protein | 34 | 0 | 0.0 | 0.0–10.3 |
| **Mesothelioma** | ERBB2 mRNA | 32 | 0 | 0.0 | 0.0–10.9 |
| **Mesothelioma** | HER2 protein | 32 | 0 | 0.0 | 0.0–10.9 |
| **NEC** | ERBB2 mRNA | 24 | 0 | 0.0 | 0.0–14.2 |
| **NEC** | HER2 protein | 24 | 0 | 0.0 | 0.0–14.2 |
| **Melanoma** | ERBB2 mRNA | 22 | 0 | 0.0 | 0.0–15.4 |
| **Melanoma** | HER2 protein | 22 | 1 | 4.5 | 0.1–22.8 |
| **Sarcoma** | ERBB2 mRNA | 16 | 0 | 0.0 | 0.0–20.6 |
| **Sarcoma** | HER2 protein | 16 | 0 | 0.0 | 0.0–20.6 |

Evaluable sample counts, positive counts, prevalence and 95% confidence intervals are reported for each tumor type and screening method at the prespecified positivity threshold.

### Supplementary Figure 11. Sensitivity of molecular HER2 prevalence to the classification threshold.

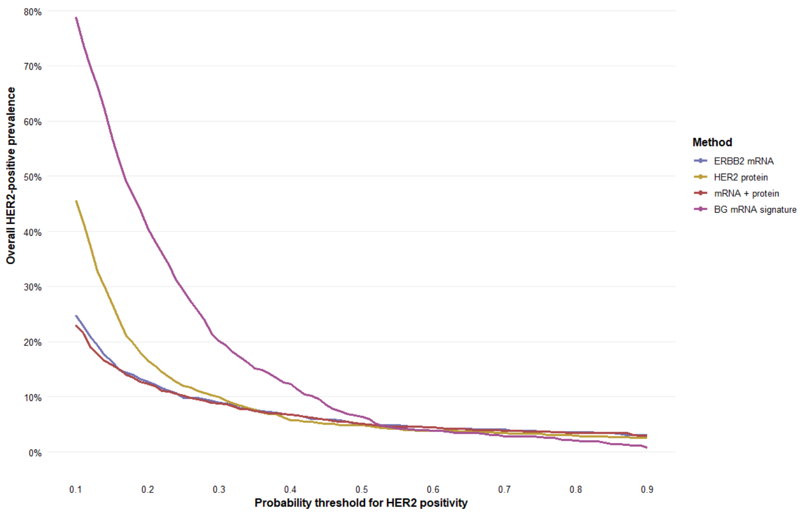

Overall HER2-positive prevalence across a range of probability thresholds for ERBB2 mRNA, HER2 protein, the mRNA + protein model and the BG RNA signature.

### Supplementary Figure 12. Intersections of genomic and molecular HER2 classifications.

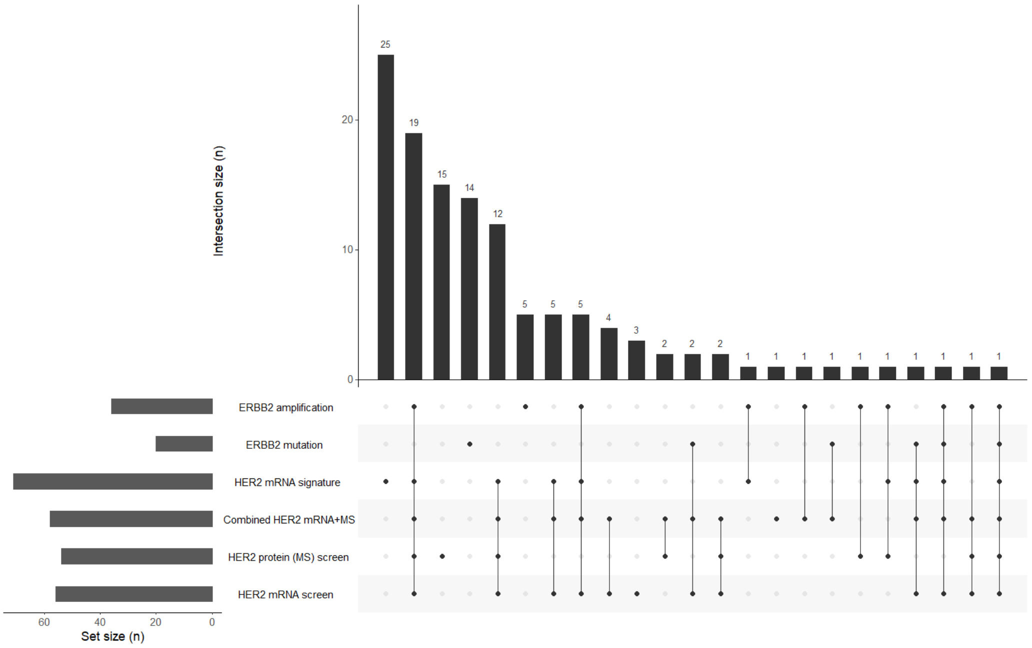

UpSet plot showing overlap among ERBB2 amplification, ERBB2 mutation and the molecular screening methods. Vertical bars show intersection sizes; horizontal bars show the number identified by each method.

### Supplementary Figure 13. Pairwise overlap of HER2 screening methods in the internal cohort.

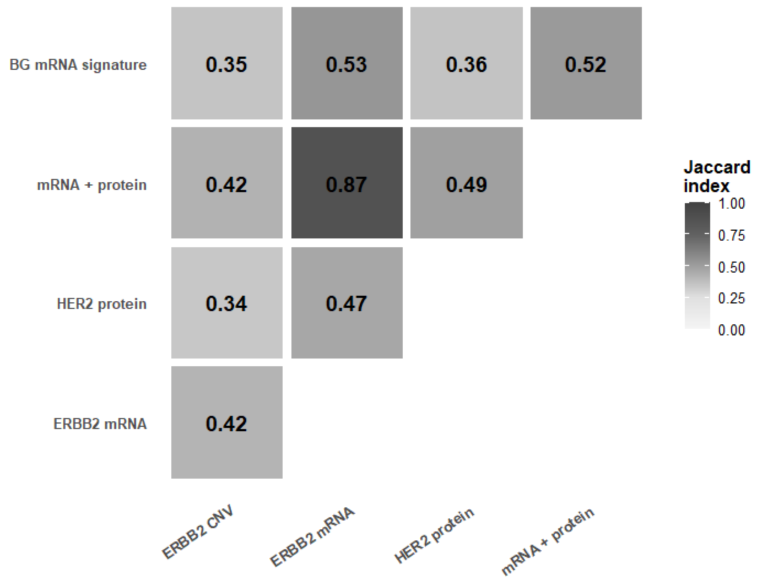

Heatmap of Jaccard indices for samples classified as positive by each pair of methods. The Jaccard index is the intersection divided by the union of the two positive sets.

### Supplementary Table 9. Intersection and union counts for internal HER2 screening methods.

| Method 1 | Method 2 | N evaluable | Intersection | Union (N) | Jaccard index |
|---|---|---|---|---|---|
| **ERBB2 mRNA** | mRNA + protein | 1130 | 53 | 61 | 0.869 |
| **ERBB2 mRNA** | BG RNA signature | 1130 | 44 | 83 | 0.530 |
| **mRNA + protein** | BG RNA signature | 1130 | 44 | 85 | 0.518 |
| **HER2 protein** | mRNA + protein | 1130 | 37 | 75 | 0.493 |
| **ERBB2 mRNA** | HER2 protein | 1130 | 35 | 75 | 0.467 |
| **ERBB2 CNV** | mRNA + protein | 1130 | 28 | 66 | 0.424 |
| **ERBB2 CNV** | ERBB2 mRNA | 1130 | 27 | 65 | 0.415 |
| **HER2 protein** | BG RNA signature | 1130 | 33 | 92 | 0.359 |
| **ERBB2 CNV** | BG RNA signature | 1130 | 28 | 79 | 0.354 |

| | | | | | |
|---|---|---|---|---|---|
| **ERBB2 CNV** | HER2 protein | 1130 | 23 | 67 | 0.343 |

Pairwise positive-set intersection counts, union counts and corresponding Jaccard indices for the internal screening methods

### Supplementary Figure 14. Leading transcripts in the PAN protein-informed RNA model.

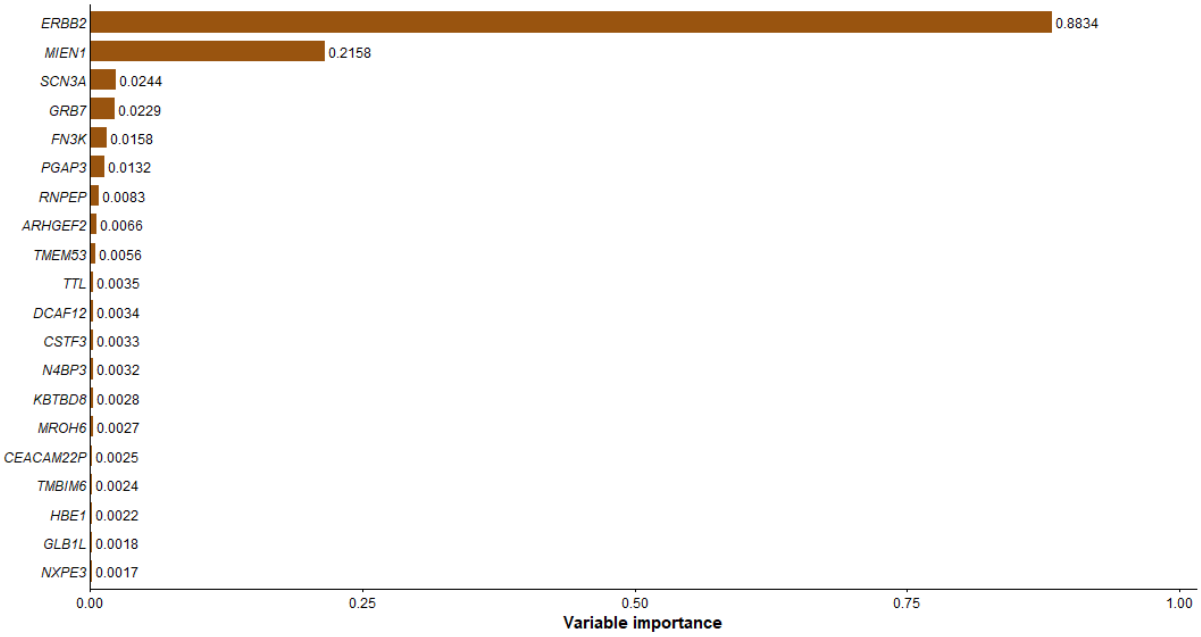

Ranked variable importance of the leading transcripts used to predict HER2 protein abundance from RNA expression.

### Supplementary Table 10. Baseline characteristics of the internal HER2-treated cohort.

| | | *N=51* | % |
|---|---|---|---|
| *Age* | Median (IQR) | 66 (59-70) | |
| *Gender* | Female | 25 | 49.0% |
| | Male | 26 | 51.0% |
| *Performance Status* | 0 | 26 | 51.0% |
| | 1 | 20 | 39.2% |
| | 2 | 5 | 9.8% |
| *HER2 Alteration* | ERBB2 amplification | 36 | 70.6% |
| | ERBB2mut | 15 | 29.4% |
| *Treatment* | Capecitabine and Trastuzumab | 1 | 2.0% |
| | Lapatinib | 1 | 2.0% |
| | Lapatinib and Trastuzumab | 6 | 11.8% |
| | Neratinib | 1 | 2.0% |
| | T-DM1 | 25 | 49.0% |
| | Trastuzumab and Pertuzuzmab | 16 | 31.4% |
| | Trastuzumab Deruxtecan (T-DXd) | 1 | 2.0% |
| *Cancer Type* | Breast cancer | 3 | 5.9% |
| | BTC | 3 | 5.9% |
| | Cervical cancer | 1 | 2.0% |
| | Colorectal cancer | 11 | 21.6% |
| | Endometrial cancer | 1 | 2.0% |
| | Head & neck cancer | 3 | 5.9% |
| | NEC | 2 | 3.9% |
| | NSCLC | 12 | 23.5% |
| | Ovarian cancer | 1 | 2.0% |
| | Pancreatic cancer | 1 | 2.0% |
| | Prostate cancer | 1 | 2.0% |
| | Salivary gland cancer | 1 | 2.0% |

| | | |
|---|---|---|
| SCLC | 1 | 2.0% |
| Small intestine cancer | 3 | 5.9% |
| Squamous Esophageal cancer | 3 | 5.9% |
| Urothelial cancer | 4 | 7.8% |

Clinical characteristics, tumor types, ERBB2 alterations and treatment regimens in the internal treated cohort. Values are counts and percentages or median (interquartile range), as indicated.

### Supplementary Figure 15. HER2 protein abundance and progression-free survival in the internal treated cohort.

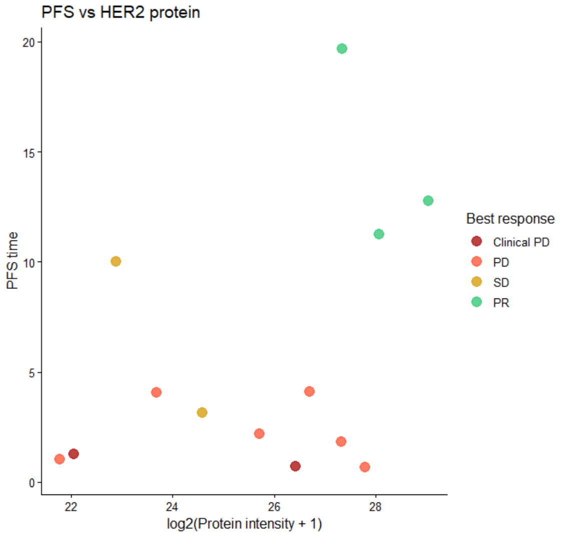

Each point represents an evaluable treated observation with measured HER2 protein abundance and progression-free survival. Colors indicate best response.

### Supplementary Table 11. Clinical outcomes by RNA-based HER2 classification in the internal treated cohort.

| | ERBB2 mRNA | | | Combined RNA Signature | | |
|---|---|---|---|---|---|---|
| | N Negative = 23 | N Positive = 28 | p | N Negative = 31 | N Positive = 20 | p |
| **CBR** | 3 (13.0%) | 11 (39.3%) | 0.058 | 7 (22.6%) | 7 (35.0%) | 0.355 |
| **ORR** | 0 (0.0%) | 6 (21.4%) | 0.027 | 0 (0.0%) | 6 (30.0%) | 0.002 |

| | | | | |
|---|---|---|---|---|
| **PFS** | 0.71 (0.40–1.26) | 0.242 | 0.64 (0.35–1.15) | 0.136 |
| **OS** | 1.41 (0.77–2.57) | 0.268 | 0.95 (0.52–1.73) | 0.858 |

Response and clinical benefit rates are reported by molecular group. Survival associations are summarized as hazard ratios with 95% confidence intervals comparing positive with negative groups for ERBB2 mRNA and the combined RNA signature.

### Supplementary Figure 16. Progression-free survival by RNA-based HER2 classification in the internal treated cohort

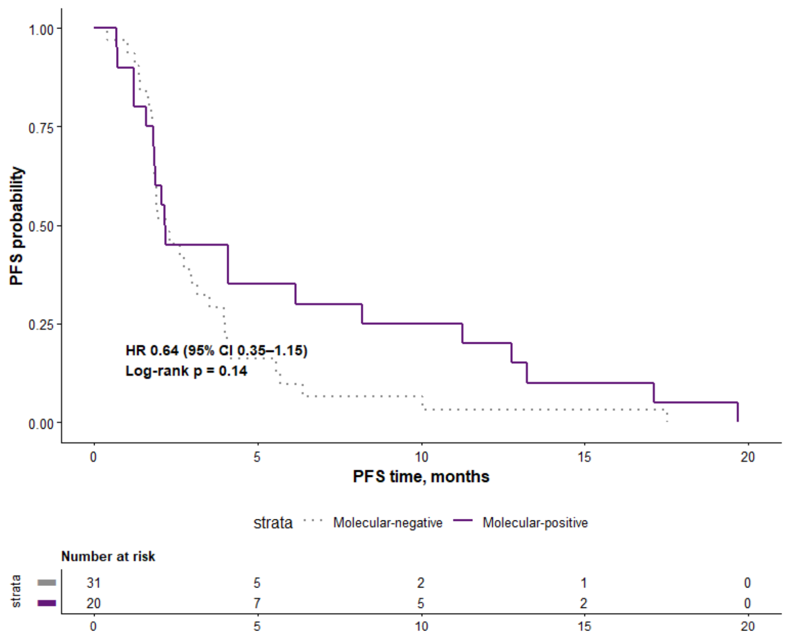

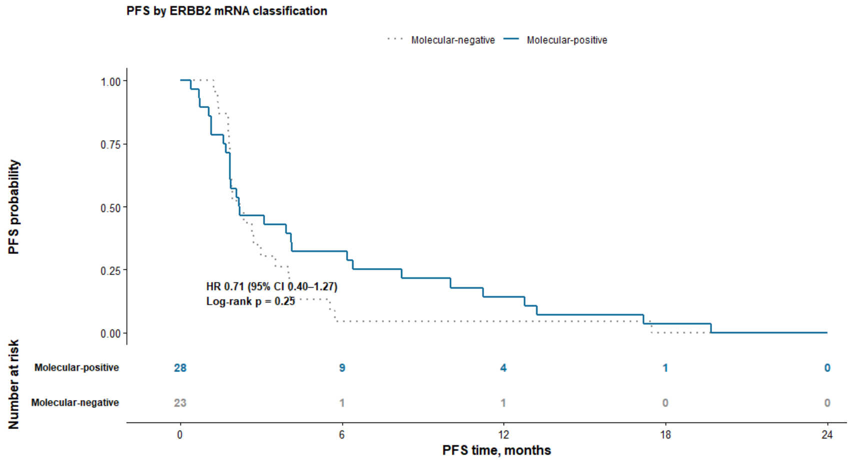

Kaplan–Meier curves stratified by the combined RNA signature and ERBB2 mRNA classification. Numbers at risk, hazard ratios with 95% confidence intervals and log-rank P values are shown.

**Supplementary Figure 17. Overall survival in relation to RNA-based HER2 measurements in the internal treated cohort.**

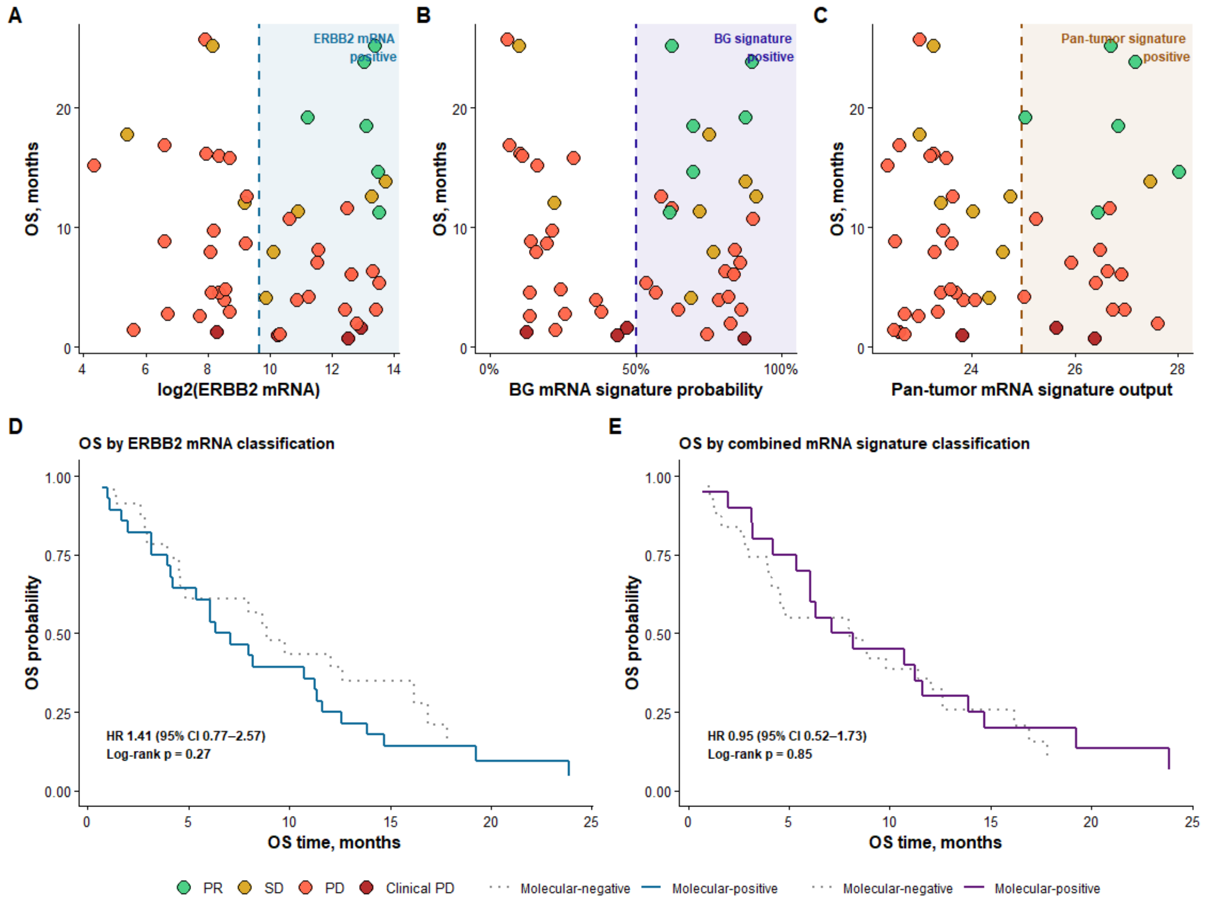

a–c, Overall survival plotted against ERBB2 mRNA, BG RNA signature probability and the PAN protein-informed RNA score, with positivity thresholds indicated. d,e, Kaplan–Meier curves by ERBB2 mRNA and combined RNA signature classification, respectively. Colors in scatterplots indicate response.

**Supplementary Table 12. Genomic and molecular characteristics of the external screening cohort.**

| | |
|---|---|
| **ERBB2 Status call** | |
| AMP Only | 106 (2.9%) |
| MUTATION Only | 64 (1.7%) |
| MUTATION+AMP | 15 (0.4%) |
| None | 3,512 (95.0%) |
| **Number of prior treatment lines** | 1.0 (0.0, 2.0) |
| **Prior treatment lines** | |
| 0 | 1,749 (47.3%) |
| 1 | 709 (19.2%) |
| 2 | 453 (12.3%) |
| ≥3 | 786 (21.3%) |
| **Biopsy type** | |
| Metastasis | 2,146 (58.0%) |
| null | 137 (3.7%) |
| Primary | 547 (14.8%) |
| Unknown | 867 (23.5%) |

**Supplementary Table 13. Primary tumor locations and biopsy sites in the external screening co-hort.**

| Primary tumor location | |
|---|---|
| Adrenal gland | 7 (0.2%) |
| Ampulla of Vater | 3 (0.1%) |
| Anus | 14 (0.4%) |
| Appendix | 4 (0.1%) |
| Biliary tract | 77 (2.1%) |
| Bladder | 109 (2.9%) |
| Bone | 11 (0.3%) |
| Bone marrow/myeloid | 3 (0.1%) |
| Brain/central nervous system | 165 (4.5%) |
| Breast | 658 (17.8%) |
| Cancer of unknown primary (CUP) | 153 (4.1%) |
| Cervix | 37 (1.0%) |
| Colon | 398 (10.8%) |
| Endometrium/uterus | 40 (1.1%) |
| Esophagus/gastroesophageal junction | 146 (3.9%) |
| Eye | 6 (0.2%) |
| Gallbladder | 22 (0.6%) |
| Hypopharynx | 3 (0.1%) |
| Kidney | 96 (2.6%) |
| Larynx | 2 (0.1%) |
| Liver | 43 (1.2%) |
| Lung | 365 (9.9%) |
| Lymphoid | 21 (0.6%) |
| Nasal cavity | 6 (0.2%) |
| Nasopharynx | 2 (0.1%) |
| Oral cavity/tongue | 9 (0.2%) |
| Oropharynx | 6 (0.2%) |
| Other | 6 (0.2%) |
| Ovary | 149 (4.0%) |
| Pancreas | 151 (4.1%) |
| Paranasal sinuses | 3 (0.1%) |

| Biopsy site | |
|---|---|
| Abdomen/abdominal wall | 99 (2.7%) |
| Adrenal gland | 32 (0.9%) |
| Anogenital region | 1 (0.0%) |
| Bladder | 13 (0.4%) |
| Bone | 192 (5.2%) |
| Brain/central nervous system | 127 (3.4%) |
| Breast | 86 (2.3%) |
| Cervix | 3 (0.1%) |
| Colon | 35 (0.9%) |
| Endometrium/uterus | 1 (0.0%) |
| Esophagus/gastroesophageal junction | 21 (0.6%) |
| Gallbladder | 3 (0.1%) |
| Kidney | 30 (0.8%) |
| Liver | 1,247 (33.7%) |
| Lung | 253 (6.8%) |
| Lymph node | 723 (19.6%) |
| Muscle/intra-muscular | 39 (1.1%) |
| Nervous system | 4 (0.1%) |
| Nose/nasopharynx | 10 (0.3%) |
| Not Available | 137 (3.7%) |
| Oral cavity/tongue | 5 (0.1%) |
| Other soft tissue | 43 (1.2%) |
| Ovary | 16 (0.4%) |
| Pancreas | 7 (0.2%) |
| Pelvic | 9 (0.2%) |
| Penis | 1 (0.0%) |
| Pericardium | 1 (0.0%) |
| Peritoneal effusion | 1 (0.0%) |
| Peritoneum/omentum | 172 (4.7%) |
| Pleura | 11 (0.3%) |
| Pleural effusion | 4 (0.1%) |

| | |
|---|---|
| Penis | 6 (0.2%) |
| Peritoneum | 1 (0.0%) |
| Pleura | 16 (0.4%) |
| Prostate | 300 (8.1%) |
| Rectum | 45 (1.2%) |
| Salivary gland | 18 (0.5%) |
| Skin | 237 (6.4%) |
| Small bowel/intestine | 48 (1.3%) |
| Soft tissue | 169 (4.6%) |
| Stomach | 38 (1.0%) |
| Testis | 3 (0.1%) |
| Thymus | 7 (0.2%) |
| Thyroid gland | 23 (0.6%) |
| Unknown | 17 (0.5%) |
| Urachus | 2 (0.1%) |
| Ureter/renal pelvis | 31 (0.8%) |
| Urethra | 8 (0.2%) |
| Vagina | 4 (0.1%) |
| Vulva | 9 (0.2%) |

| | |
|---|---|
| Prostate | 13 (0.4%) |
| Rectum | 6 (0.2%) |
| Retroperitoneal | 12 (0.3%) |
| Salivary gland | 3 (0.1%) |
| Skin (cutaneous) | 13 (0.4%) |
| Skin (subcutaneous) | 218 (5.9%) |
| Small bowel/intestine | 6 (0.2%) |
| Soft tissue of extremity | 7 (0.2%) |
| Soft tissue of head and neck | 5 (0.1%) |
| Spleen | 1 (0.0%) |
| Stomach | 11 (0.3%) |
| Thorax/thorax wall | 21 (0.6%) |
| Thyroid gland | 1 (0.0%) |
| Trachea | 1 (0.0%) |
| Unknown | 36 (1.0%) |
| Vagina | 13 (0.4%) |
| Vulva | 4 (0.1%) |

Counts and percentages of external screening samples by primary tumor locationand biopsy site.

**Supplementary Figure 18. Overlap between ERBB2 amplification and RNA-based HER2 classification in the external cohort.**

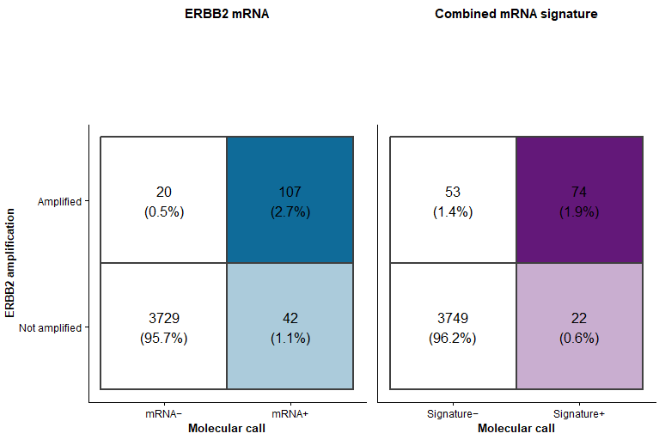

**Supplementary Figure 19. Pairwise overlap of HER2 screening methods in the external cohort.**

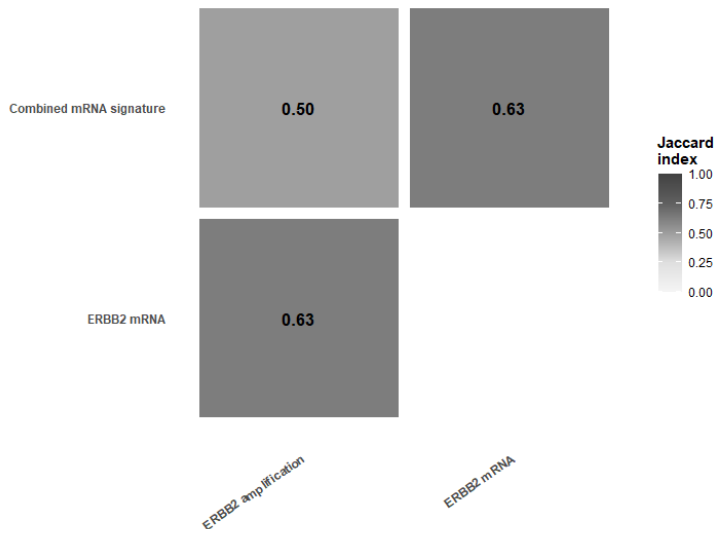

Jaccard indices summarize overlap among samples positive for ERBB2 amplification, ERBB2 mRNA and the combined RNA signature.

### Supplementary Table 14. Baseline characteristics of the external HER2-treated cohort.

**Baseline characteristics of HER2-treated RNA validation cohort**

| Characteristic | Overall, N = 113[1] |
|---|---|
| **Age at HER2 treatment start, years** | 60.0 (52.0, 67.0) |
| **Sex** | |
| female | 87 (77.0%) |
| male | 26 (23.0%) |
| **Primary tumor location** | |
| Biliary tract | 1 (0.9%) |
| Breast | 67 (59.3%) |
| Colon | 6 (5.3%) |
| Esophagus/gastroesophageal junction | 18 (15.9%) |
| Lung | 14 (12.4%) |
| Rectum | 3 (2.7%) |
| Salivary gland | 1 (0.9%) |
| Stomach | 3 (2.7%) |
| **Biopsy site** | |
| Adrenal gland | 1 (0.9%) |
| Bone | 7 (6.2%) |
| Brain/central nervous system | 1 (0.9%) |
| Breast | 9 (8.0%) |
| Liver | 54 (47.8%) |
| Lung | 12 (10.6%) |
| Lymph node | 19 (16.8%) |
| Other soft tissue | 1 (0.9%) |
| Peritoneum/omentum | 2 (1.8%) |
| Skin (subcutaneous) | 7 (6.2%) |
| **Biopsy type** | |

**Baseline characteristics of HER2-treated RNA validation cohort**

| Characteristic | Overall, N = 113[1] |
|---|---|
| Metastasis | 87 (77.0%) |
| Primary | 17 (15.0%) |
| Unknown | 9 (8.0%) |
| **HER2-targeted treatment** | |
| Trastuzumab | 54 (47.8%) |
| Trastuzumab + Ado-trastuzumab emtansine (T-DM1) | 2 (1.8%) |
| Trastuzumab + Pertuzumab | 57 (50.4%) |
| **ERBB2 status** | 67 (59.3%) |
| **ERBB2 Amplification** | 59 (52.2%) |
| **ERBB2 Amplification and mutation** | 8 (7.1%) |
| **ERBB2 Mutation** | 4 (3.5%) |
| **Prior treatment lines, grouped** | |
| 0 | 43 (38.1%) |
| 1 | 22 (19.5%) |
| 2 | 13 (11.5%) |
| ≥3 | 35 (31.0%) |

Clinical characteristics and treatment regimens for 113 matched biopsy–treatment episodes from 102 unique patients. Values are counts and percentages or median (interquartile range), as indicated.

**Supplementary Table 15. Associations between RNA-based HER2 classification and outcomes in the external treated cohort.**

| Endpoint | Method | N | Effect | Estimate (95% CI) | P value |
|---|---|---|---|---|---|
| **ORR** | ERBB2 mRNA | 102 | OR | 2.03 (0.86–4.77) | 0.142 |
| **ORR** | Combined RNA signature | 102 | OR | 1.81 (0.81–4.07) | 0.157 |
| **CBR** | ERBB2 mRNA | 102 | OR | 3.35 (1.30–8.62) | 0.0148 |
| **CBR** | Combined RNA signature | 102 | OR | 2.44 (0.88–6.81) | 0.0991 |
| **ToT** | ERBB2 mRNA | 110 | HR | 0.42 (0.27–0.64) | 0.0000456 |
| **ToT** | Combined RNA signature | 110 | HR | 0.57 (0.38–0.86) | 0.0066 |
| **OS** | ERBB2 mRNA | 112 | HR | 0.41 (0.25–0.65) | 0.000171 |
| **OS** | Combined RNA signature | 112 | HR | 0.59 (0.37–0.94) | 0.0276 |

Odds ratios for objective response and clinical benefit – mixed responses excluded, and hazard ratios for time on treatment and overall survival, compare molecular-positive with molecular-negative groups. Endpoint-specific denominators, event or response counts, 95% confidence intervals and P values are reported. Response odds ratios are cross-product estimates with Wald 95% confidence intervals; response P values are from two-sided Fisher's exact tests.

**Supplementary Figure 20. Time on treatment and overall survival by combined RNA signature status in the external cohort.**

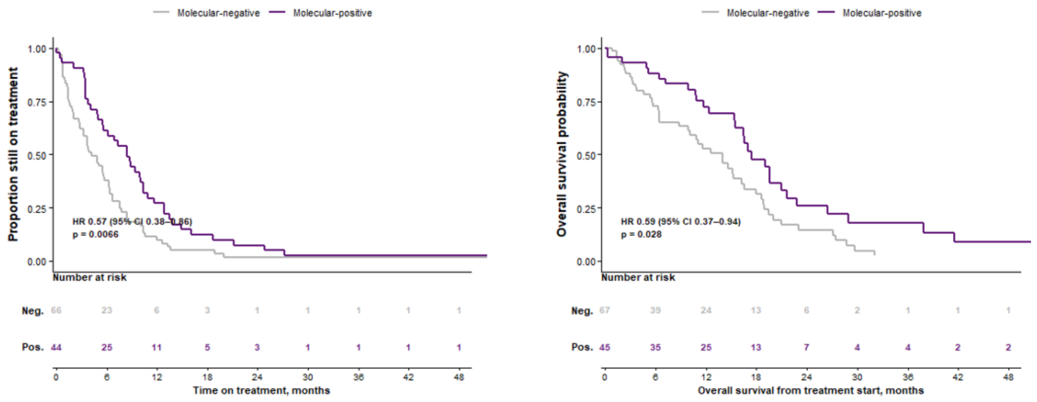

Kaplan–Meier curves for time on treatment and overall survival stratified by combined RNA signature status. Numbers at risk and hazard ratios with 95% confidence intervals are shown.

### Supplementary Table 16. Associations of continuous ERBB2 copy number and mRNA expression with external treatment outcomes.

| Predictor | N | Events | Standard errors | Effect | Estimate (95% CI) | p | AIC | AUC | C-index |
|---|---|---|---|---|---|---|---|---|---|
| **PR/CR** | | | | | | | | | |
| **ERBB2 copy number** | 102 | 41 | Model-based | OR per 1 SD increase | 1.23 (0.82–1.91) | 0.325 | 140.4 | 0.60 | NA |
| **ERBB2 mRNA** | 102 | 41 | Model-based | OR per 1 SD increase | 1.43 (0.95–2.20) | 0.090 | 138.5 | 0.61 | NA |
| **CBR** | | | | | | | | | |
| **ERBB2 copy number** | 102 | 78 | Model-based | OR per 1 SD increase | 1.19 (0.74–2.29) | 0.530 | 114.9 | 0.63 | NA |
| **ERBB2 mRNA** | 102 | 78 | Model-based | OR per 1 SD increase | 1.75 (1.10–2.87) | 0.020 | 109.6 | 0.64 | NA |
| **Time on treatment** | | | | | | | | | |
| **ERBB2 copy number** | 110 | 104 | Model-based | HR per 1 SD increase | 0.93 (0.72–1.19) | 0.553 | 769.7 | NA | 0.61 |
| **ERBB2 mRNA** | 110 | 104 | Model-based | HR per 1 SD increase | 0.65 (0.52–0.81) | <0.001 | 756.2 | NA | 0.61 |
| **Overall survival** | | | | | | | | | |
| **ERBB2 copy number** | 112 | 77 | Model-based | HR per 1 SD increase | 1.01 (0.79–1.29) | 0.951 | 564.8 | NA | 0.40 |
| **ERBB2 mRNA** | 112 | 77 | Model-based | HR per 1 SD increase | 0.65 (0.51–0.84) | <0.001 | 553.6 | NA | 0.60 |

Odds ratios and hazard ratios are reported per standard-deviation increase in each continuous predictor, with endpoint-specific denominators, 95% confidence intervals and P values.

**Supplementary Figure 21. Time on treatment in relation to continuous genomic and RNA-based HER2 measurements.**

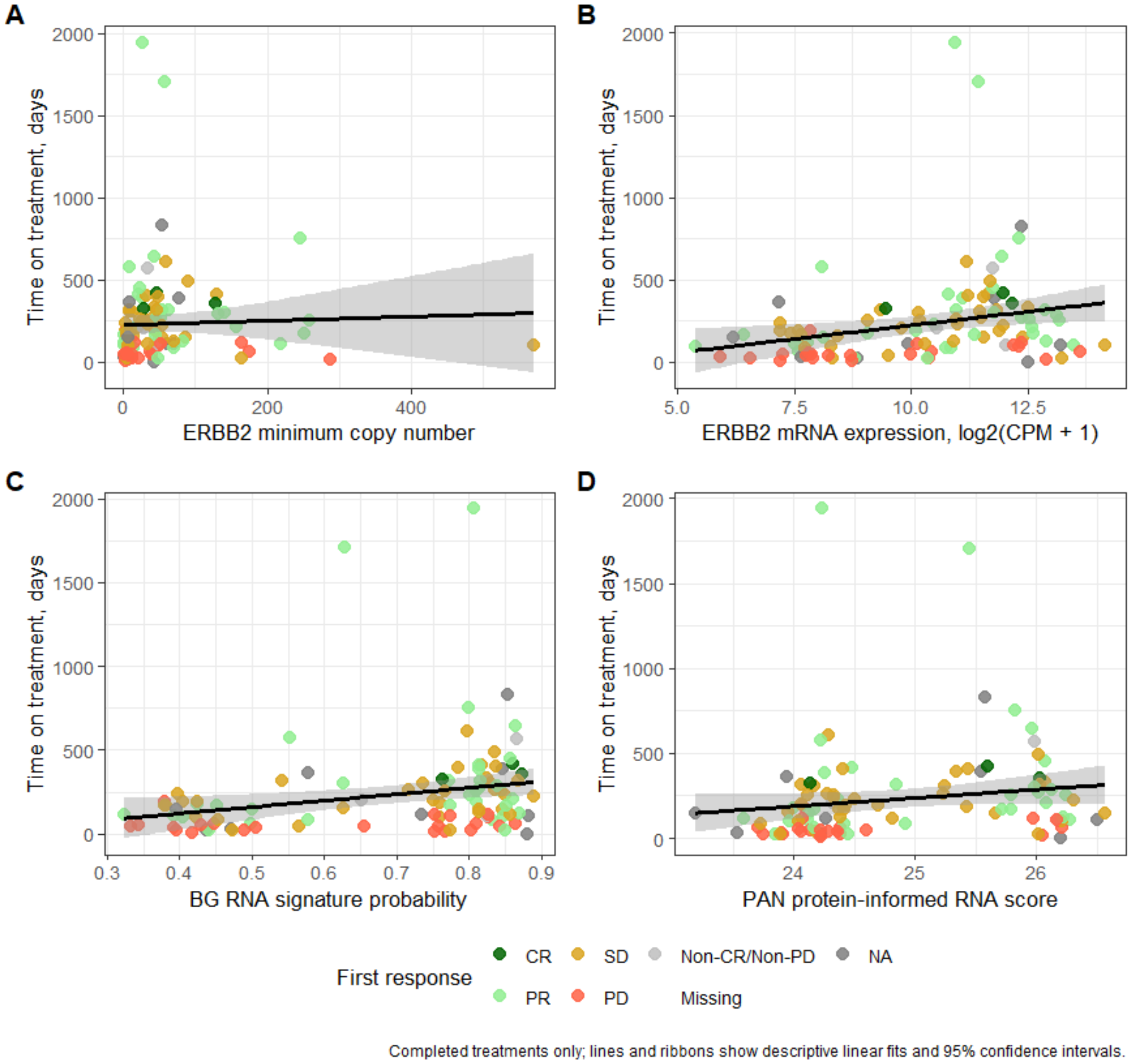

Time on treatment plotted against ERBB2 copy number, ERBB2 mRNA, BG RNA signature probability and the PAN protein-informed RNA score. Points are colored by response; fitted trends and uncertainty bands are displayed.

**Supplementary Figure 22. Subgroup associations between RNA-based HER2 classification and treatment outcomes.**

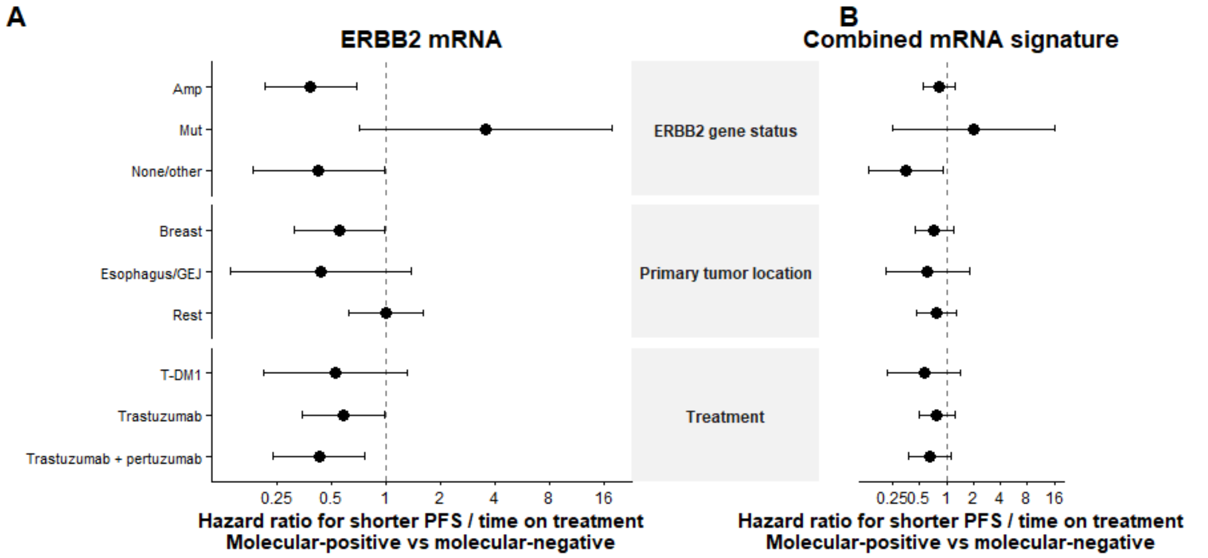

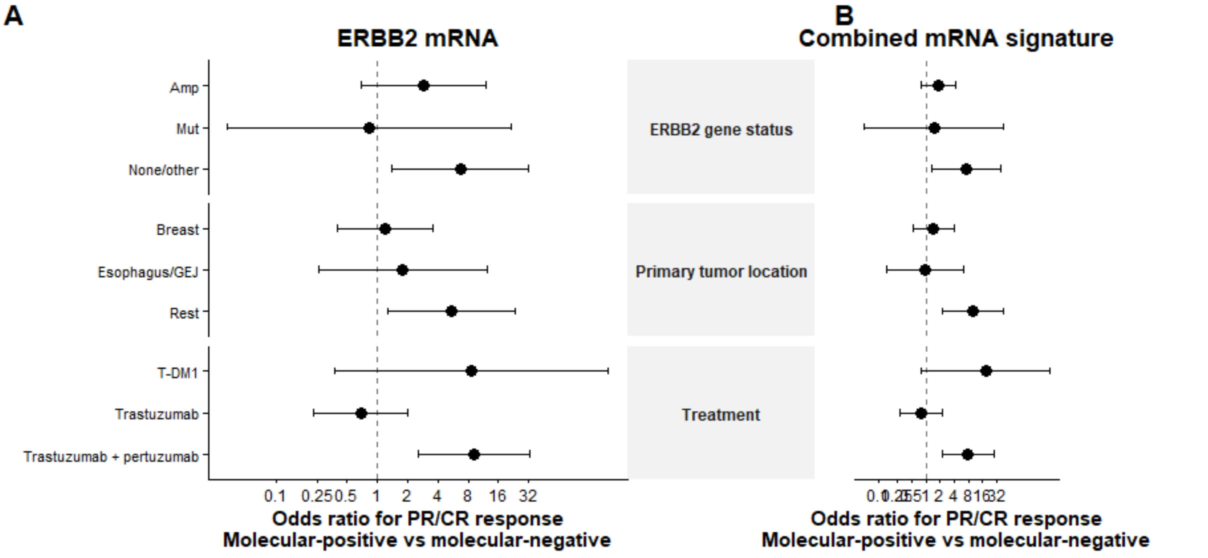

Forest plots show survival hazard ratios and objective-response odds ratios for ERBB2 mRNA and the combined RNA signature across genomic, tumor-type and treatment subgroups. Estimates compare molecular-positive with molecular-negative groups; horizontal bars show 95% confidence intervals.

**Supplementary Table 17. Treatment-outcome associations in the pooled cohort excluding ERBB2-mutated tumors.**

| Endpoint | Method | N | Events / responses | Effect | Estimate (95% CI) | P value |
|---|---|---|---|---|---|---|
| ORR | ERBB2 mRNA | 127 | 43 | OR | 3.12 (1.29–7.55) | 0.0103 |
| ORR | Combined RNA signature | 127 | 43 | OR | 2.25 (1.06–4.76) | 0.0396 |
| CBR | ERBB2 mRNA | 127 | 81 | OR | 3.05 (1.42–6.57) | 0.0060 |
| CBR | Combined RNA signature | 127 | 81 | OR | 1.68 (0.80–3.49) | 0.198 |
| ToT/PFS | ERBB2 mRNA | 134 | 129 | HR | 0.49 (0.33–0.71) | 0.000152 |
| ToT/PFS | Combined RNA signature | 134 | 129 | HR | 0.59 (0.42–0.85) | 0.0039 |
| OS | ERBB2 mRNA | 136 | 107 | HR | 0.47 (0.31–0.70) | 0.000254 |
| OS | Combined RNA signature | 136 | 107 | HR | 0.60 (0.41–0.89) | 0.0103 |

Associations of ERBB2 mRNA and combined RNA signature positivity with response, clinical benefit, progression-free survival/time on treatment and overall survival. Endpoint-specific sample sizes, event or response counts, effect estimates, 95% confidence intervals and P values are reported

### Supplementary Table 18. IHC/FISH distribution of reference samples included in DVP analyses.

| Cancer type | N | IHC0 | Ultra-low | IHC1+ | IHC2+/FISH− | IHC2+/FISH unavailable | IHC2+/FISH+ | IHC3+ |
|---|---|---|---|---|---|---|---|---|
| **Breast** | 46 | 2 (4.3%) | 5 (10.9%) | 11 (23.9%) | 12 (26.1%) | 0 (0.0%) | 6 (13.0%) | 10 (21.7%) |
| **Gastroesophageal** | 33 | 1 (3.0%) | 0 (0.0%) | 14 (42.4%) | 13 (39.4%) | 1 (3.0%) | 3 (9.1%) | 1 (3.0%) |

IHC/FISH categories among 46 breast and 33 gastroesophageal reference samples retained after applying the ≥20-tile criterion. Values are n (%), with percentages calculated within each cancer type.

### Supplementary Figure 23. Tile counts and proteomic depth across DVP samples.

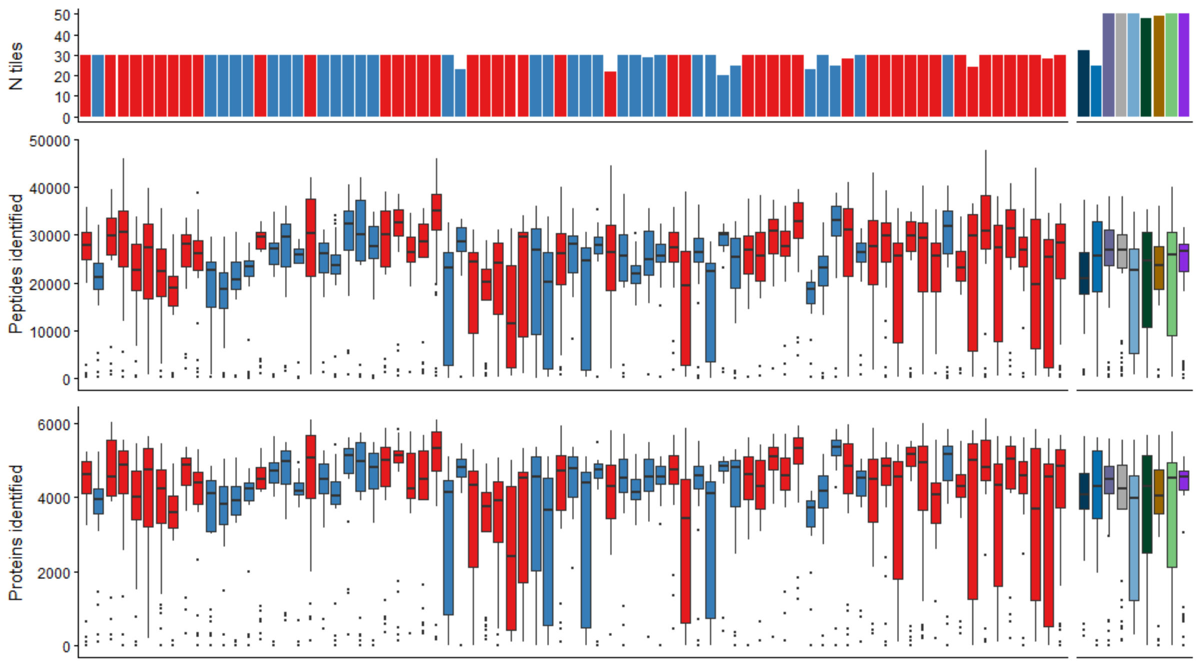

Upper panel shows the number of tiles per sample. Middle and lower panels show the distributions of quantified peptides and proteins across tiles within each sample.

#### Supplementary Table 19. Spatial HER2 detection and intensity across clinical IHC/FISH categories.

| IHC/FISH category | N | HER2-detected tiles, median % (IQR) | Samples with detected HER2, n | HER2 intensity, median $log_2$ (IQR) |
|---|---|---|---|---|
| **IHC0/Ultra-low** | 8 | 10.0% (0.0–23.3%) | 5 | 13.37 (12.88–14.28) |
| **IHC1+** | 25 | 23.3% (10.0–43.3%) | 21 | 14.13 (13.78–14.76) |
| **IHC2+/FISH−** | 25 | 50.0% (23.3–72.7%) | 25 | 14.27 (14.03–14.49) |
| **IHC2+/FISH+** | 9 | 56.7% (53.3–64.0%) | 9 | 15.09 (14.06–15.43) |
| **IHC3+** | 11 | 66.7% (48.3–78.3%) | 11 | 16.22 (15.40–17.79) |

Slide-level HER2 detection percentages and median intensities among detected tiles are summarized as median (interquartile range). The analysis includes 78 reference samples with ≥20 tiles and resolved IHC/FISH status. Slides without detected HER2 contribute only to detection summaries.

**Supplementary Figure 24: Leave-one-slide-out DVP classification across clinical IHC/FISH categories**

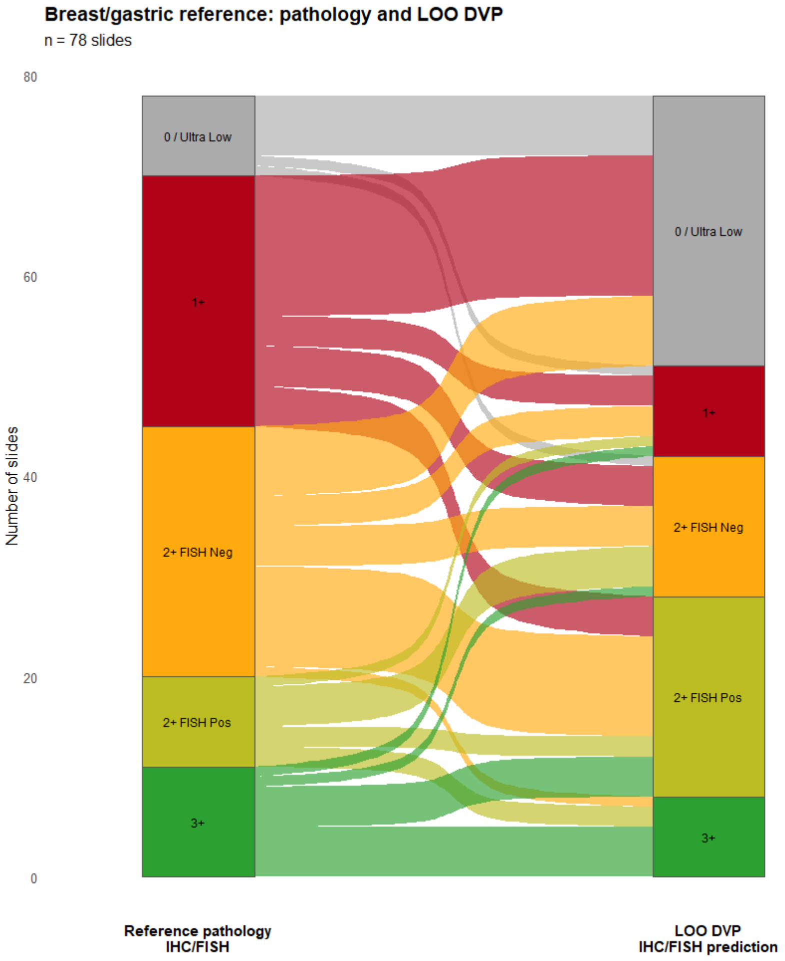

Alluvial plot comparing clinical reference IHC/FISH categories with DVP categories assigned by leave-one-slide-out reference-curve matching in 78 breast and gastroesophageal slides. Each flow represents one slide; flow color indicates its clinical reference category.

#### Supplementary Figure 25: Routine pathology, central pathology and DVP classifications in breast cancer

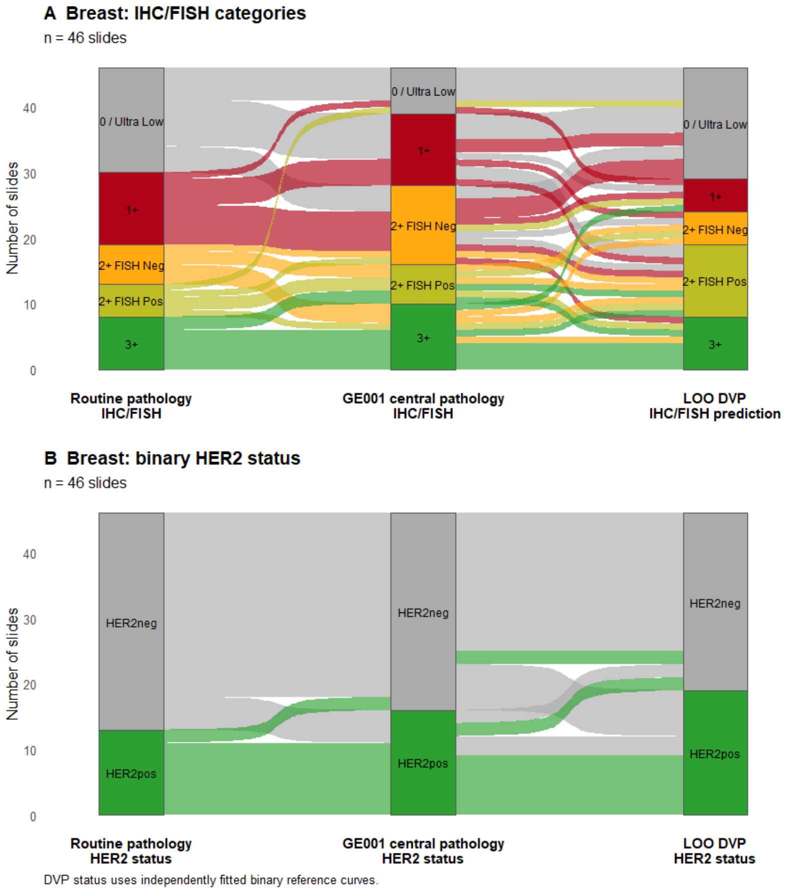

Three-way comparison of routine clinical pathology, central GE001 pathology and leave-one-slide-out DVP assignments for IHC/FISH categories (**A**) and binary HER2 status (**B**). Each flow represents one breast slide. Binary DVP status was assigned using independently fitted binary reference curves.

### Supplementary table 20: Agreement among routine pathology, central pathology and DVP classification in breast cancer

| Analysis method | Comparison | N | Agreement (95% CI) |
|---|---|---|---|
| **IHC/FISH category** | Routine pathology vs GE001 central pathology | 46 | 39.1% (25.1-54.6%) |
| **IHC/FISH category** | Routine pathology vs LOO DVP prediction | 46 | 45.7% (30.9-61.0%) |
| **IHC/FISH category** | GE001 central pathology vs LOO DVP prediction | 46 | 32.6% (19.5-48.0%) |
| **Binary HER2 status** | Routine pathology vs GE001 central pathology | 46 | 84.8% (71.1-93.7%) |
| **Binary HER2 status** | Routine pathology vs LOO DVP prediction | 46 | 69.6% (54.2-82.3%) |
| **Binary HER2 status** | GE001 central pathology vs LOO DVP prediction | 46 | 76.1% (61.2-87.4%) |

Pairwise slide-level agreement for the three assessments, shown separately for IHC/FISH category and binary HER2 status. Parentheses give exact binomial 95% confidence intervals.

### Supplementary Figure 26: DVP classification of tumors discordant between clinical pathology and bulk HER2 assays

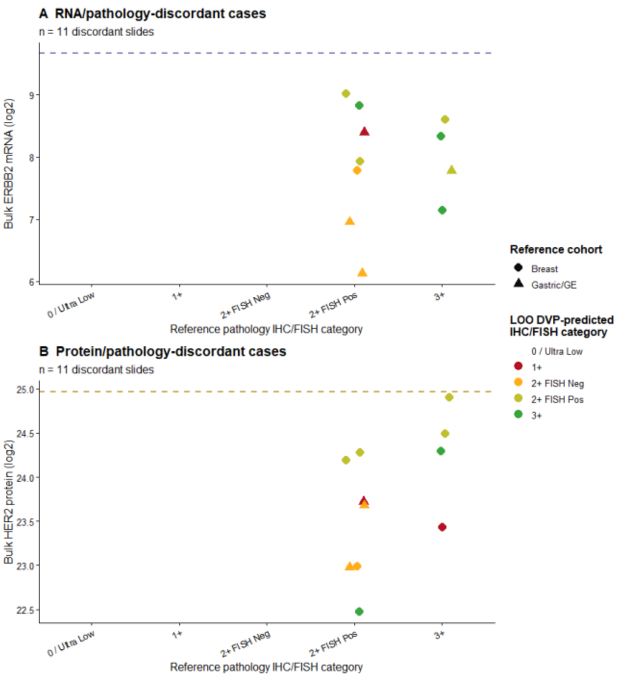

Clinical reference IHC/FISH categories and leave-one-slide-out DVP assignments for slides discordant between clinical HER2 status and bulk ERBB2 mRNA (**A**) or bulk HER2 protein (**B**). Each panel includes slides with an available measurement for that assay. Dashed lines mark the respective bulk positivity thresholds; point shape indicates reference cohort and color indicates the DVP-assigned category.

**Supplementary Figure 27. Agreement between routine and central HER2 assessment in breast cancer.**

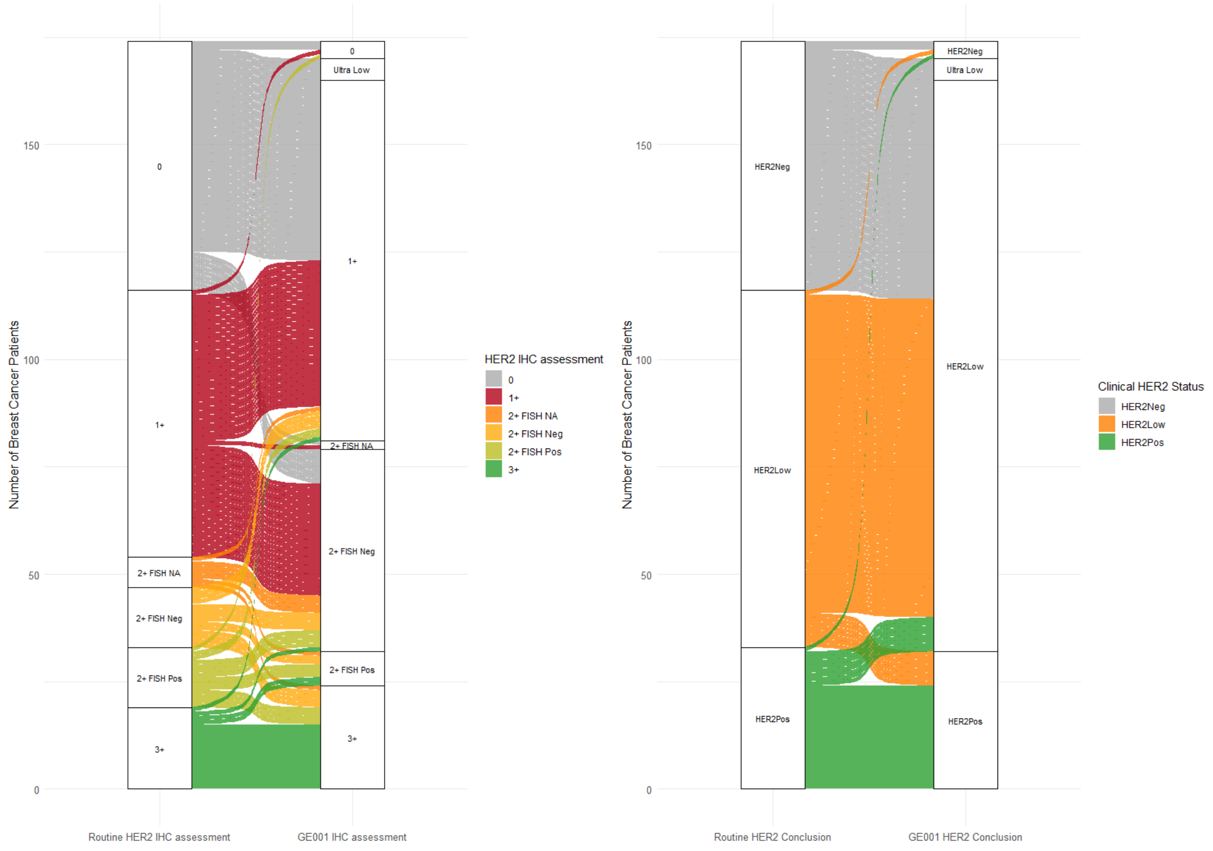

Alluvial plots show transitions from routine diagnostic assessment to central HercepTest mAb pharmDx assessment. Left, individual IHC/FISH categories; right, grouped HER2 classifications. Flow widths represent sample counts.

**Supplementary Figure 28. Expression of candidate reference transcripts across tumor types.**

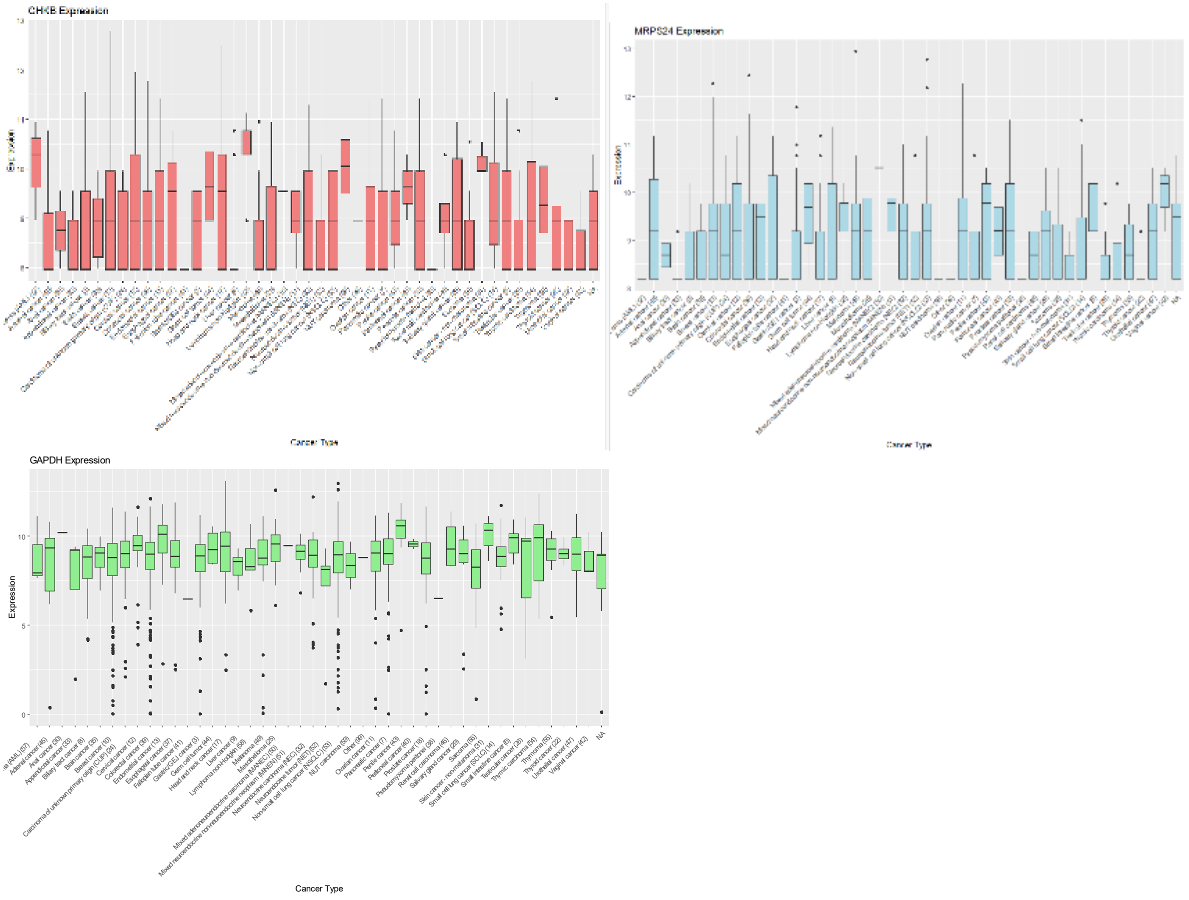

.

Distributions of GAPDH, CHKB and MRPS24 expression across tumor types, evaluated as candidate denominators for ERBB2 RNA ratio normalization.

### Supplementary Figure 29. Biopsy-site composition across tumor types.

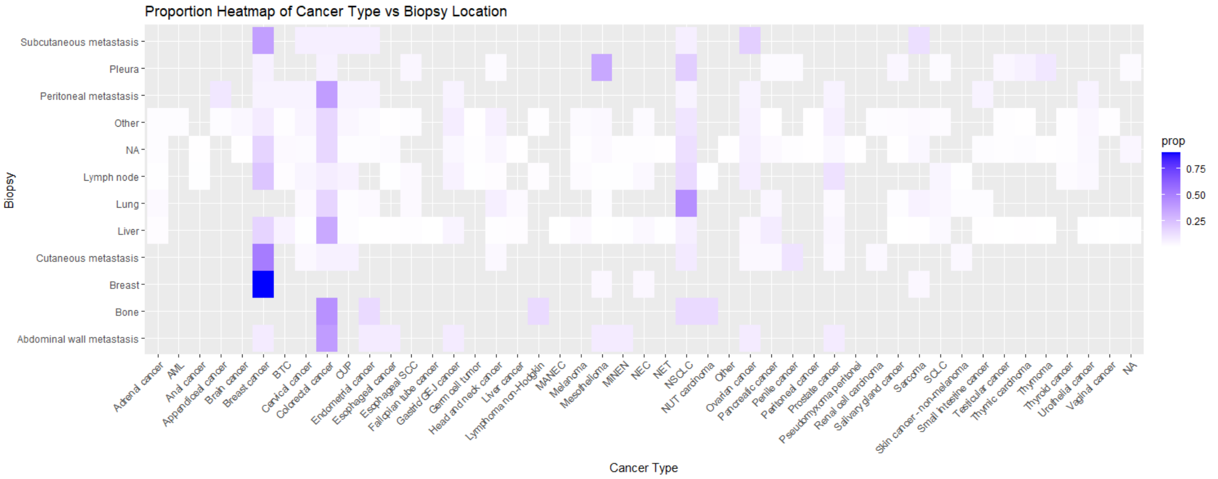

Heatmap showing biopsy-site proportions within tumor types. Only samples with available tumor-type and biopsy-site annotation are included.

### Supplementary Figure 30. Estimated biopsy-site effects on ERBB2 mRNA and HER2 protein abundance.

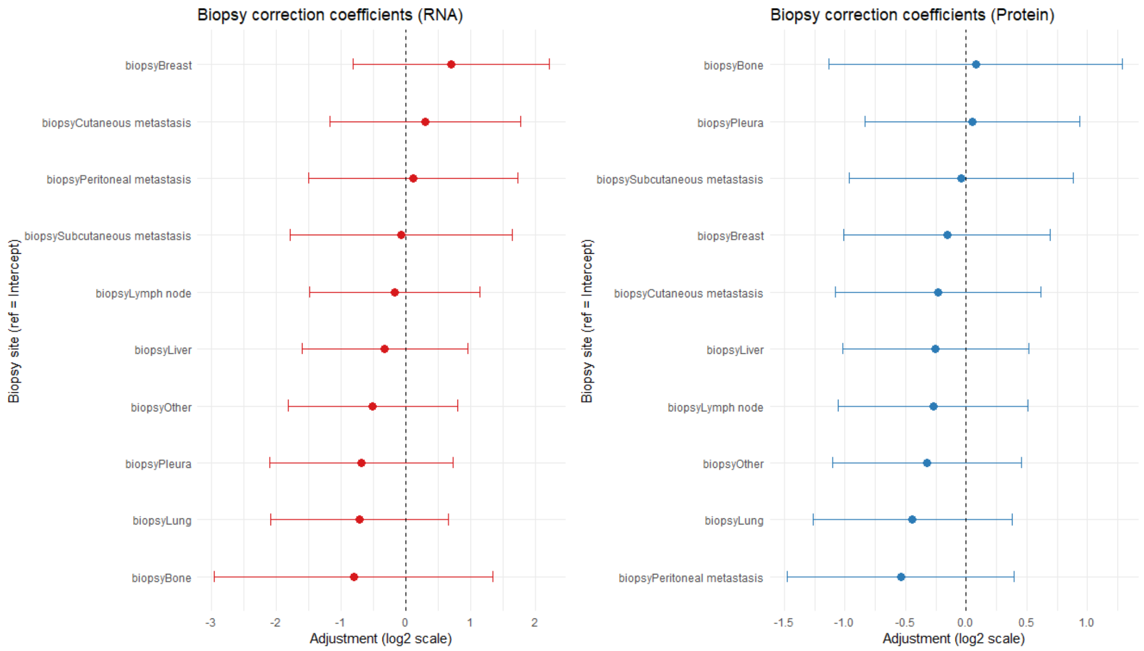

Points and horizontal bars show regression coefficients and 95% confidence intervals for biopsy site relative to the reference category, on the $\log_2$ scale. RNA and protein estimates are shown separately.

### Supplementary Figure 31. RNA-derived stromal scores across biopsy sites.

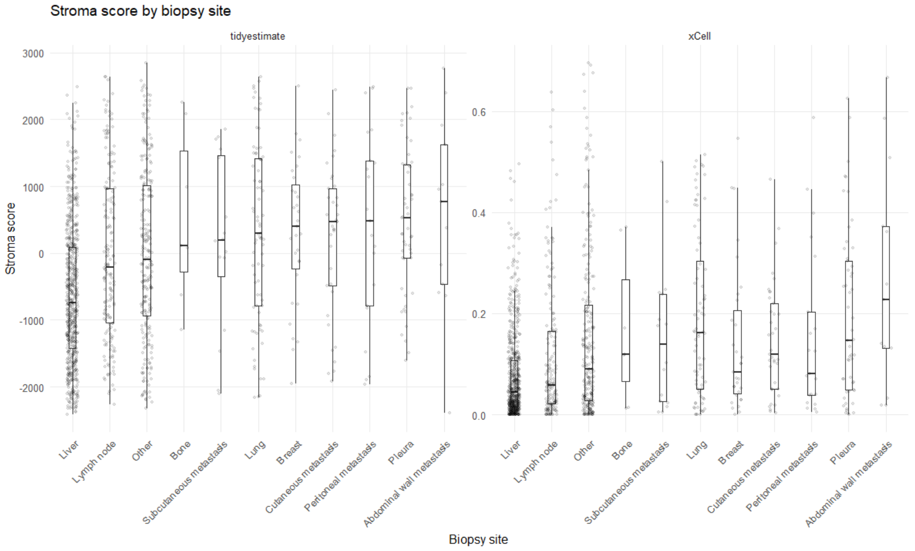

Distributions of ESTIMATE and xCell stromal scores by biopsy site. Boxes show medians and interquartile ranges, with individual sample values overlaid.

### Supplementary Figure 32. Biopsy-site effects on ERBB2 mRNA with corresponding stromal scores

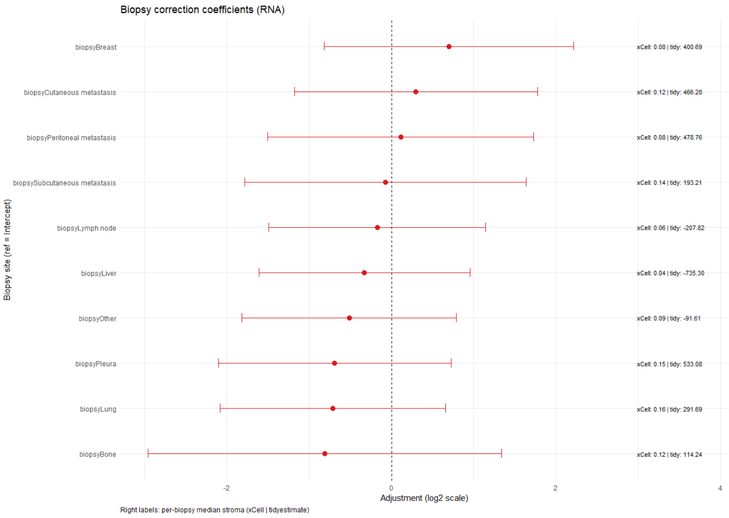

Regression coefficients and 95% confidence intervals for biopsy-site effects on ERBB2 mRNA. Adjacent annotations report site-specific median xCell and ESTIMATE stromal scores.

**Supplementary Table 21. Sensitivity analysis of RNA model development.**

A) Internal discrimination and screening prevalence

| Cohort | Outcome | N | Primary models | Sensitivity-analysis models |
|---|---|---|---|---|
| **Breast/gastroesophageal reference** | BG AUC (95% CI) | 171 | 0.862 (0.793–0.931) | 0.841 (0.764–0.917) |
| **Internal pan-tumor scree-ning** | BG positive n; % (95% CI) | 1,130 | 71; 6.28% (4.94–7.86%) | 60; 5.31% (4.08–6.78%) |
| **Internal pan-tumor scree-ning** | Combined positive n; % (95% CI) | 1,130 | 33; 2.92% (2.02–4.08%) | 34; 3.01% (2.09–4.18%) |
| **External pan-tumor screen-ing (HMF)** | Combined positive n; % (95% CI) | 3,898 | 96; 2.46% (2.00–3.00%) | 97; 2.49% (2.02–3.03%) |

B) Agreement in combined signature classification

| Cohort | N | Concordant n (%) | Negative → positive, n | Positive → negative, n |
|---|---|---|---|---|
| **Internal pan-tumor screening** | 1,130 | 1,129 (99.91%) | 1 | 0 |
| **Internal treated** | 51 | 50 (98.04%) | 1 | 0 |
| **External pan-tumor screening (HMF)** | 3,898 | 3,897 (99.97%) | 1 | 0 |
| **External treated (HMF)** | 113 | 113 (100.00%) | 0 | 0 |

C) Associations with treatment outcomes

| Cohort and endpo-int | Type | Primary estimate (95% CI) | P value | Sensitivity analysis estimate (95% CI) | P value |
|---|---|---|---|---|---|
| **Internal treated ORR** | OR | ∞ (2.19–∞) | 0.0022 | ∞ (1.99–∞) | 0.0030 |
| **Internal treated PFS** | HR | 0.64 (0.35–1.15) | 0.136 | 0.68 (0.38–1.23) | 0.205 |
| **HMF treated ORR** | OR | 1.80 (0.75–4.40) | 0.157 | 1.80 (0.75–4.40) | 0.157 |
| **HMF treated CBR** | OR | 2.42 (0.81–8.28) | 0.099 | 2.42 (0.81–8.28) | 0.099 |
| **HMF treated ToT** | HR | 0.57 (0.38–0.86) | 0.0066 | 0.57 (0.38–0.86) | 0.0066 |
| **HMF treated OS** | HR | 0.59 (0.37–0.94) | 0.028 | 0.59 (0.37–0.94) | 0.028 |

a, Internal discrimination and screening prevalence. b, Agreement in combined RNA signature classification. c, Treatment-outcome associations under the primary and sensitivity-analysis models. The sensitivity analysis evaluates feature selection within cross-validation folds; clinical effect estimates compare positive with negative groups within each pipeline. Response odds ratios are conditional maximum-likelihood estimates with exact 95% confidence intervals and two-sided P values from Fisher's exact test. Small differences from Supplementary Table 15 reflect the estimation method
